# A fast and robust strategy to remove variant level artifacts in Alzheimer’s Disease Sequencing Project data

**DOI:** 10.1101/2021.10.28.21265577

**Authors:** Michael E. Belloy, Yann Le Guen, Sarah J. Eger, Valerio Napolioni, Michael D. Greicius, Zihuai He

## Abstract

Whole-exome sequencing (WES) and whole-genome sequencing (WGS) are expected to be critical to further elucidate the missing genetic heritability of Alzheimer’s disease (AD) risk by identifying rare coding and/or noncoding variants that contribute to AD pathogenesis. In the United States, the Alzheimer’s Disease Sequencing Project (ADSP) has taken a leading role in sequencing AD-related samples at scale, with the resultant data being made publicly available to researchers to generate new insights into the genetic etiology of AD. In order to achieve sufficient power, the ADSP has adapted a study design where subsets of larger AD cohorts are collected and sequenced across multiple centers, using a variety of sequencing kits. This approach may lead to variable variant quality across sequencing centers and/or kits. Here, we performed exome-wide and genome-wide association analyses on AD risk using the latest ADSP WES and WGS data releases. We observed that many variants displayed large variation in allele frequencies across sequencing centers/kits and contributed to spurious association signals with AD risk. We also observed that sequencing kit/center adjustment in association models could not fully account for these spurious signals. To address this issue, we designed and implemented novel filters that aim to capture and remove these center/kit-specific artifactual variants. We conclude by deriving a novel, fast, and robust approach to filter variants that represent sequencing center- or kit-related artifacts underlying spurious associations with AD risk in ADSP WES and WGS data. This approach will be important to support future robust genetic association studies on ADSP data, as well as other studies with similar designs.

**Author Summary:** Next generation sequencing data represents a highly valuable resource to uncover rare coding and/or noncoding genetic variants that contribute to Alzheimer’s disease risk. In order to achieve large sample sizes that are required for such data, the Alzheimer’s Disease Sequencing Project (ADSP) has taken the leading role in sequencing Alzheimer’s disease related samples at scale in the United States. The ADSP’s study design however leads to variable variant quality across the involved sequencing centers, necessitating a quality control approach that ensures robust genetic association analyses. Here, we present and validate a rigorous quality control pipeline, where we specifically developed a new strategy to handle inter-center variant quality issues in the ADSP. In doing so, we provide a first glance into exome- and genome-wide associations with Alzheimer’s disease risk using the latest releases of ADSP data (respectively 20.5k and 16.9k individuals). In sum, our pipeline is important to support future robust genetic association studies on ADSP data, as well as other studies with similar design. This in turn will contribute to accelerating Alzheimer’s disease gene discovery and gene-driven therapy development.

## Introduction

Late-onset Alzheimer’s disease (AD) is marked by a strong genetic component, with heritability estimates ranging from 59% to 79%^1,2^. Largely supported by single nucleotide polymorphism (SNP) genotyping arrays and variant imputation, large-scale meta-analyses of genome-wide association studies have so far implicated over 50 loci relevant to AD in subjects of European ancestry^2–6^. Despite these important advances, most risk variants identified so far have common allele frequencies and it’s estimated that only about half of AD’s genetic heritability has been captured, such that much of AD’s genetic component remains to be identified^2^. In response to this observation, there has been a shift to start using whole-exome sequencing (WES) or whole-genome sequencing (WGS) to help capture rare and/or coding variants that contribute to AD risk, which has led to several recent initial successes^7–15^.

In the United States, the Alzheimer’s Disease Sequencing Project (ADSP) has taken a leading role in sequencing of AD-related samples at scale, with resultant data being made publicly available to researchers to generate new insights into the genetic etiology of AD. In order to achieve sufficient power to support analyses of sequencing data and rare variants, the ADSP has adapted a study design where subsets of larger AD cohorts are collected and sequenced across multiple centers, using a variety of sequencing kits^16–18^. This in turn can lead to “center” or “kit” effects that traditionally are accounted for by using center/kit covariate adjustment. However, a prior study using a prior version of the ADSP WES discovery phase observed that center/kit covariate adjustment could not account for variable variant qualities across centers and kits, which in turn may lead to spurious associations or impact the identification of AD-associated risk variants^19^.

Since then, the ADSP has further expanded its efforts and as of 2021 provides WES and WGS data on respectively 20.5k and 16.9k individuals across diverse ancestries^18^. In our exploratory analyses of these data, we observed many variants that displayed large variation in allele frequencies across centers/kits and contributed to spurious association signals with AD risk. Similar to the prior study^19^, we also observed that kit/center adjustment could not fully account for these signals. Thus, in the current study, we design and implement novel filters that aim to capture and remove these center/kit-specific artifactual variants. We additionally test filters containing putatively artifactual variants identified in the gnomAD reference database^20^. The filters are designed such that they can be implemented post-hoc to association analyses, leaving flexibility to researchers to either run full sample analyses with robust variant quality control, or, to identify variants that require targeted analyses.

## Methods

### Ascertainment of Genotype and Phenotype Data

Genotype data for subjects with AD-related clinical outcome measures were available from the Alzheimer’s Disease Sequencing project (ADSP) whole exome sequencing (WES) and whole genome sequencing (WGS) data. Notably, the ADSP performed targeted sequencing of samples in case-control (majority), family-based, population-based, and longitudinal cohorts, performing sequencing across multiple sequencing centers and using various sequencing kits (**Table S1-2**). Ascertainment of genotype/phenotype data for these samples is described in detail elsewhere^18^. In addition to the ADSP samples, we also had access to several publicly available SNP microarray and WGS datasets (**Table S1**), largely comprising data from the Alzheimer’s Disease Genetics Consortium (ADGC). The latter have a large degree of sample overlap with ADSP. In order to ensure the most up-to-date and parsimonious phenotypes, we performed a cross-sample genotype/phenotype harmonization, which is summarized in **Supplementary Methods**.

Participants or their caregivers provided written informed consents in the original studies. The current study protocol was granted an exemption by the Stanford Institutional Review Board because the analyses were carried out on “de-identified, off-the-shelf” data.

### Genetic Data Quality Control and Processing

The ADSP WES and WGS data (NG00067.v5) were joint called by the ADSP following the SNP/Indel Variant Calling Pipeline and data management tool used for analysis of whole genome and exome sequencing (WGS/WES) for the Alzheimer’s Disease Sequencing Project (VCPA)^21^. The WES data was currently only released for bi-allelic variants, which the ADSP has quality controlled. The WGS data was released for bi-allelic and multi-allelic variants separately, which the ADSP had not yet quality controlled. The current analyses of ADSP WGS were restricted to bi-allelic variants, to which we applied the Variant Quality Score Recalibration (VSQR) quality control filter (“PASS” variants; GATK v4.1)^22^. The WES/WGS data were available in genome build hg38, which we annotated using dbSNP153 variant identifiers.

Genetic data underwent standard quality control (QC). Detailed descriptions of all processing procedures and sequential sample filtering steps are in **Supplementary Methods** and **Table S3-4**. For the purpose of the presented genetic association analyses, only non-Hispanic subjects of European ancestry were considered to focus on the largest ancestry population (SNPweights v2.1; **Figure S1**)^23^. Principal component analysis of genotyped SNPs provided principal components (PCs) capturing population substructure (PC-AiR, **Figure S2**)^24^. In both the WES and WGS data respectively, variants with a genotyping rate less than 95%, deviating from Hardy Weinberg Equilibrium (HWE) in the full sample or in controls (p<10^−6^), and a minor allele count less than 10, were excluded. After this standard quality control, the total number of remaining variants was 224,270 for ADSP WES and 14,772,936 for ADSP WGS.

### Primary filters to remove sequencing center/kit-related variant level artifacts

We designed filters to assess whether there were significant deviations in genotype distributions for any given variant across sequencing centers and kits respectively. To avoid bias from frequency differences across cases and controls, we only assessed genotypes in control individuals.

The primary filters made use of the fast Fisher exact test as implemented by Plink (v.1.9; command -- fisher)^25^. However, this test can currently only be implemented by comparing two groups at a time (e.g. two genotyping centers) while we observed variant issues across multiple groups. We therefore compared every individual sequencing center/kit to all others and combined the P-values from the multiple tests through the Cauchy combination test^25^. Variants with a combined P-value lower than the heuristic threshold of 10^−5^ were flagged to be filtered.

We additionally tested two other types of sequencing center/kit-based variant filters. On one hand, we performed chi square tests (R v.3.6.0) that respectively considered all sequencing centers or kits at once. Variants with a P-value lower than the heuristic threshold of 10^−5^ were flagged to be filtered. On the other hand, we performed Fisher tests with Monte Carlo (MC) simulation of P-values (R v.3.6.0) that respectively considered all sequencing centers or kits. The MC approach was chosen to allow feasible run times. Variants with a P-value lower than the heuristic threshold of 10^−3^ were flagged to be filtered (this threshold reflects that the P-values from MC simulation are less small than those obtained for the other tests).

The three filters were compared in terms of speed by calculating the time needed to derive the respective variant filters on a 1MB genetic region of chromosome 1 in ADSP WGS. Computing time was evaluated on a single CPU from an 80-core Xeon Gold 6138T processor @ 2.00GHz.

### Filters from the Genome Aggregation Database (gnomAD)

In addition to the filters proposed above, we used the gnomAD data base (v3.1.1) reference to identify potential variant artifacts^20^. Specifically, we created filters for variants that have: (1) a “non-PASS” flag in gnomAD, corresponding to those that did not pass gnomAD sample quality control filters and may thus be more prone to sequencing issues; (2) a “LCR” flag in gnomAD, corresponding to those located in a Low Complexity Region and may thus be more prone to low coverage, read misalignment, and subsequent genotype issues; (3) a differential frequency of more than 10% between our current samples and non-Finish European (nfe) participants in gnomAD, which may indicate an issue with those variants in our samples. The three gnomAD filters were evaluated with the goal of supporting the primary ADSP WES/WGS center/kit-based variant filters.

### Filters for discordant variants across duplicate samples

A final set of filters was designed to flag variants that are discordant across duplicate samples. Notably, the ADSP WES and WGS data both respectively contain a few hundred duplicate samples, generally covering multiple sequencing centers and/or kits. Discordant variants across such duplicates therefore provide a reference of artifactual variants that should be removed and are largely reflecting center/kit-related genotyping issues. We evaluated these filters with the primary goal of comparing them with the primary ADSP WES/WGS center/kit-based variant filters as well as the gnomAD-based variant filters. In a secondary goal, we also assessed to what extent these duplicate discordant variant filters themselves could handle center/kit-related variant issues that drove observations of spurious association signals.

### Statistical analyses, Variant Annotation, and Visualization

Exome-wide and genome-wide association studies on AD case-control status were conducted respectively on ADSP WES and WGS, using LMM-BOLT (v.2.3.5). LMM-BOLT employs a Bayesian mixture model that allows the inclusion of related individuals by adjusting for the genetic relationship matrix (GRM)^26^, thereby maximizing sample size and power. Given the current minor allele count thresholds, the approximate fifty-fifty ratio of cases to controls, and sample sizes exceeding 5,000 participants for both ADSP WES and WGS, the resultant test statistics are expected to be well-calibrated^26^. After analyses, association statistics were transformed back to a logistic scale taking into account the case fraction^26^. Per convention, variants were considered at suggestive (P<=10^−5^) or genome-wide (P<=5×10^−8^) significance.

Case-control association analyses considered two models. Model-1 included covariates for sex, *APOE*\*4 dosage, *APOE*\*2 dosage, and the first 5 genetic PCs. We did not adjust for age as we previously showed that this can lead to significant power loss when the age of cases is younger than for controls^15^, which is true for ADSP given their initial design to prioritize old controls and young cases (**Table 1 & Table S5-6**). Model-2 was the same as Model-1 but additionally included covariates for sequencing center and kit.

**Table 1.**
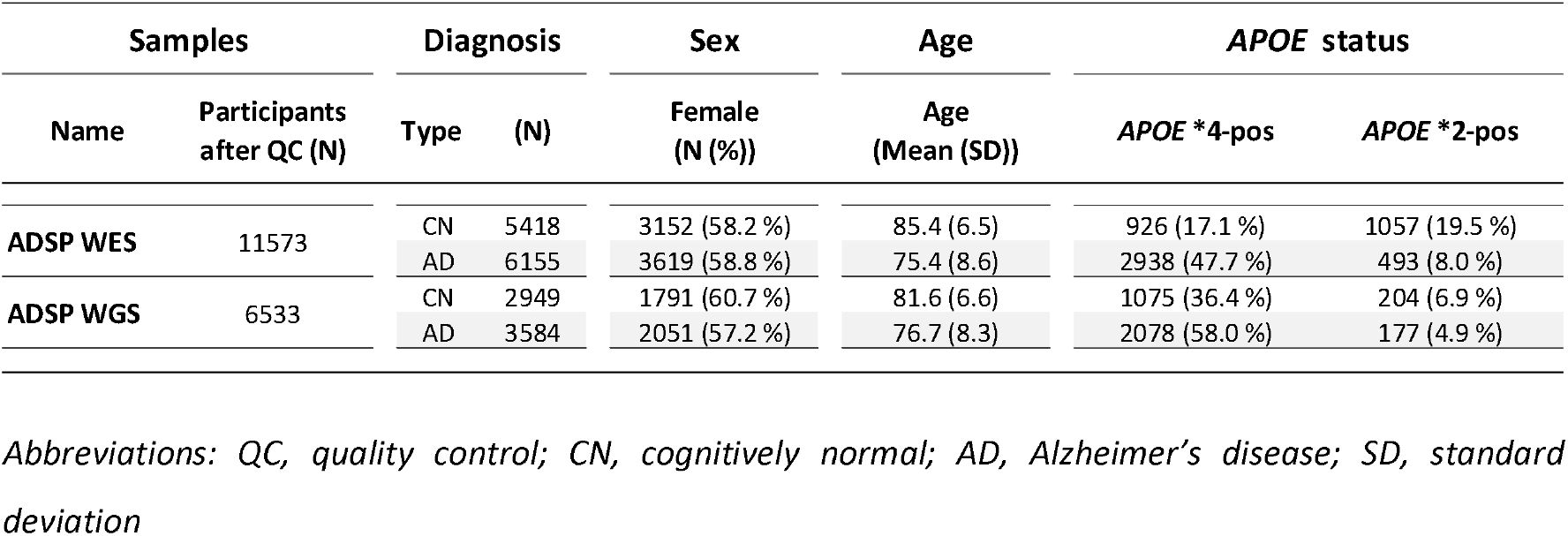
Sample demographics. Samples were restricted to those passing genetic/phenotypic quality control, being non-Hispanic, and being of European ancestry.

| Samples |  | Diagnosis |  | Sex | Age | APOE status |  |
| --- | --- | --- | --- | --- | --- | --- | --- |
| Name | Participants after QC (N) | Type | (N) | Female (N (%)) | Age (Mean (SD)) | APOE *4-pos | APOE *2-pos |
| ADSP WES | 11573 | CN | 5418 | 3152 (58.2 %) | 85.4 (6.5) | 926 (17.1 %) | 1057 (19.5 %) |
|  |  | AD | 6155 | 3619 (58.8 %) | 75.4 (8.6) | 2938 (47.7 %) | 493 (8.0 %) |
| ADSP WGS | 6533 | CN | 2949 | 1791 (60.7 %) | 81.6 (6.6) | 1075 (36.4 %) | 204 (6.9 %) |
|  |  | AD | 3584 | 2051 (57.2 %) | 76.7 (8.3) | 2078 (58.0 %) | 177 (4.9 %) |

*Abbreviations: QC, quality control; CN, cognitively normal; AD, Alzheimer's disease; SD, standard deviation*

The *APOE* locus (1Mb region centered on *APOE*) was removed from all summary statistics. Independent loci were determined by sliding window when no variants with P<=10^−5^ were observed within 200Kb from one another. Manhattan plots provide RefSeq curated gene annotations for the gene closest (<500Kb) to the top significant variant per locus. Only variants with P<=10^−6^ were annotated to improve visualization. Suggestive significance levels were indicated by gray dotted lines and green dots for variants. Genome-wide significance levels were indicated by black solid lines and red dots for variants. Variant densities were indicated at the bottom of Manhattan plots (dark green = low density, yellow=medium density, red = high density). Plots were generated using the R package CMplot^27^.

## Results

Sample demographics are provided in **Table 1**, with per center/kit demographics in **Table S5-6**. In initial exome and genome-wide analyses using model-1, we observed many spurious associations (P<=1e-5). We identified that variants underlying these spurious signals displayed increased variation in allele frequency across sequencing centers/kits for the full frequency range (**Figure 1A-B**). We also observed that such variants could not consistently be accounted for by adjustment for sequencing center/kit in model-2; A specific example of such a variant is provided in **Figure 1C**.

**Figure 1.**
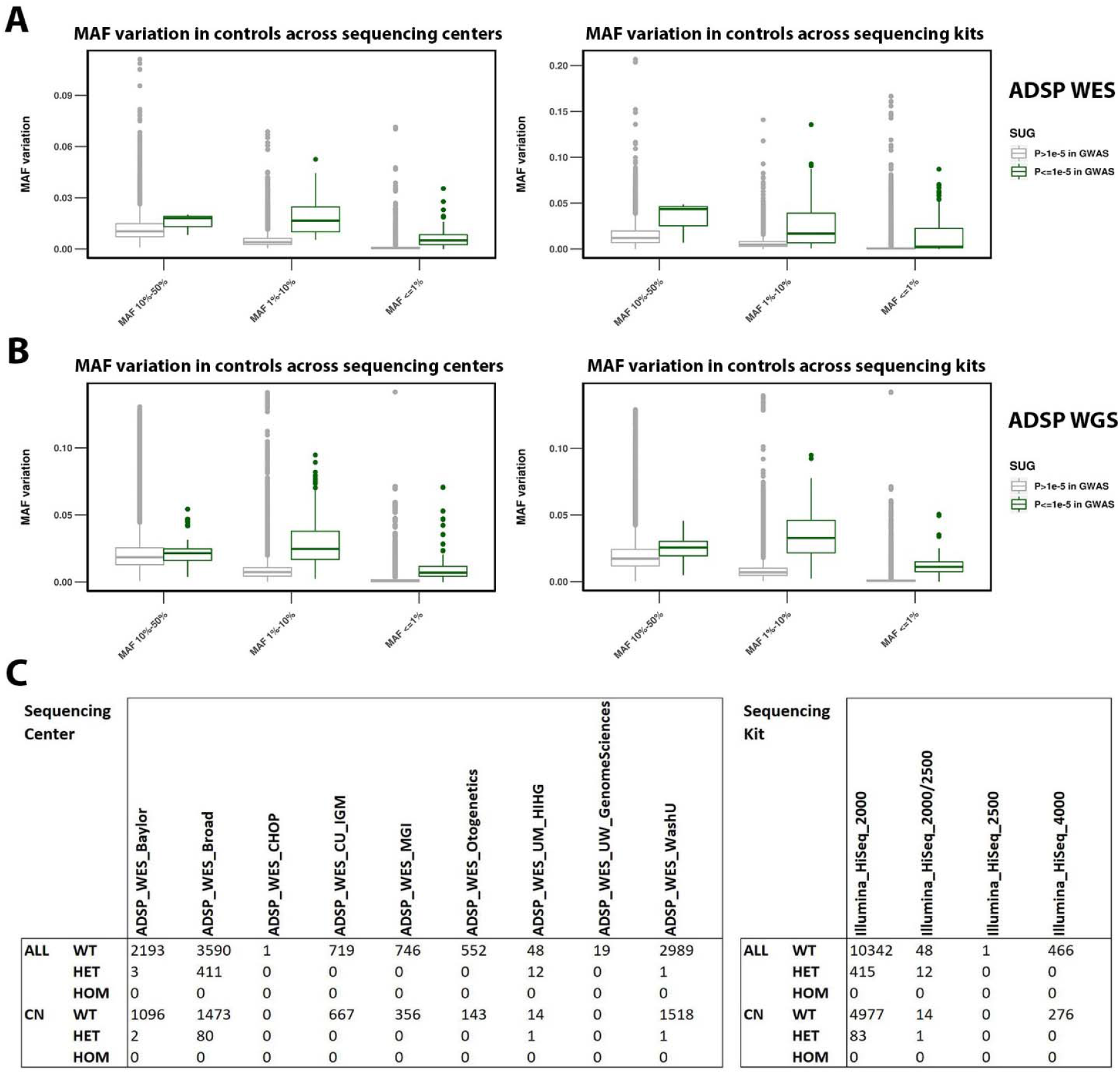
Variant artifacts across different sequencing centers/kits drive spurious associations in ADSP WES and WGS data. In initial exome-wide and genome-wide association studies of ADSP WES and WGS, we observed many spurious associations (P<=1e-5) using model-1 (i.e. not adjusting for sequencing center/kit; **cf. Figure 2A & 3A**). Upon inspection of these signals, it was notable that these variants displayed large variation in genotype counts across sequencing centers/kits. The MAF variation in control subjects for all analyzed variants is visualized in **A**) for ADSP WES, and in **B)** for ADSP WGS. **C**) A specific example of a variant showing spurious association is provided. This variant, rs199707443, has a MAF of 0.003% in non-Finnish Europeans in gnomAD v3.1.1, contrasting the 411 heterozygote counts in the Broad sequencing center. Notably, this particular variant still showed genome-wide significant association with AD risk even after sequencing center/kit adjustment (**cf. Figure 2B**). *Abbreviations: MAF, minor allele frequency; CN, cognitively normal; WT; Wild type; HET, heterozygote; HOM, homozygote*.

Based on these observations, three versions of filters were designed and evaluated for their capacity to capture putative center/kit-related variant artifacts. In assessing computing time, the filter using the Fisher exact test implemented in Plink followed by Cauchy combination of P-values implemented in R proved to be the fastest, taking 32 seconds to be constructed using a single CPU for a 1Mb region in ADSP WGS (5,402 variants). Comparatively, constructing the chi square test filter implemented in R took 93 seconds, while the Fisher test with MC filter implemented in R took 128 seconds. Given the faster speed, as well as the expected higher robustness provided by an exact test, we present the filter using the Fisher exact test implemented in Plink as the primary filter, while the other two represent supporting analyses. Throughout the remainder of the manuscript we will use the term “filtered” to describe variants that were removed by filters and the term “non-filtered” to describe variants that were not removed by filters.

The Fisher exact center/kit-based variant filters showed they strongly reduced the number of spurious associations observed with model-1 in ADSP WES (**Figure 2A & 2C**) and WGS (**Figure 3A & 3C**). When further adjusting for sequencing center/kit in model-2, spurious associations appeared essentially absent in ADSP WES (**Figure 2D**) and WGS (**Figure 3D**). Notably, the spurious associations did not appear to be driven by inflation, as for instance the genomic control factor (λ) was consistent prior to and after applying variant filters in ADSP WGS for the respective models (**Figure 3**). The slightly larger λ for ADSP WES in model-1 prior to applying the variant filters (**Figure 2A**) indicated that the large number of spurious variants with regard to the relatively small total set of variants was likely driving some modest inflation. Consistent observations were made for the other two center/kit-based variant filters (**Figure S3-6**). When intersecting variants identified across these three sets of filters, the filter derived from the fisher exact test implemented in Plink overlapped strongly (>96%) with the other two filters that in turn showed less overlap (**Figure S7)**. This was consistent with the Fisher exact test being the most conservative and robust.

**Figure 2.**
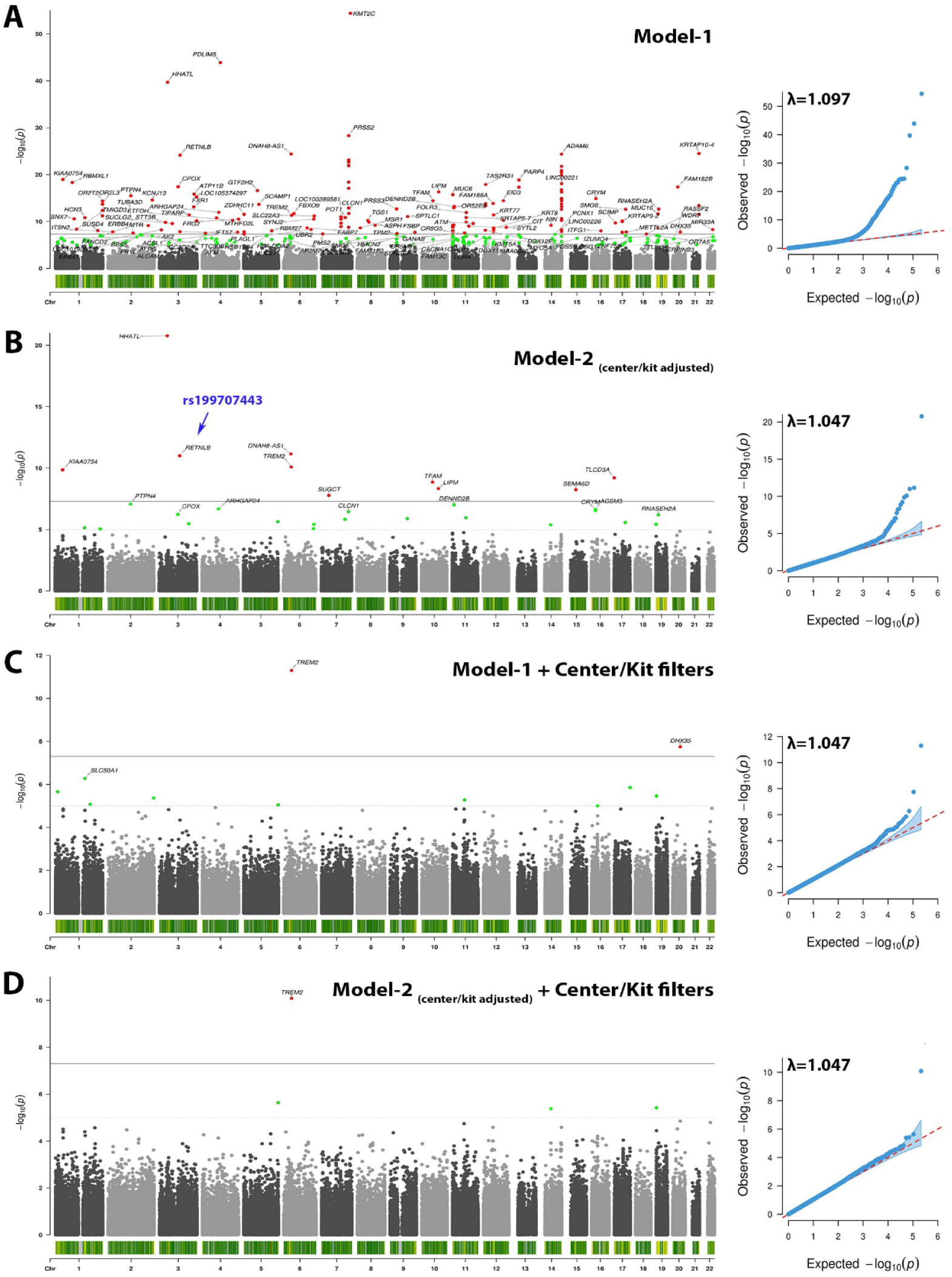
The proposed center/kit-based variant filters remove spurious associations in ADSP WES. Figure shows Manhattan (left) and quantile-quantile (right) plots. **A)** Model-1 indicates many spurious hits. **B)** Model-2 shows that adjustment for center/kit can reduce many, but not all, spurious hits. The variant described in **Figure 1C** is highlighted by the blue arrow. **C)** Filters remove most spurious hits. **D)** Further adjustment for center/kit removes few additional spurious hits.

**Figure 3.**
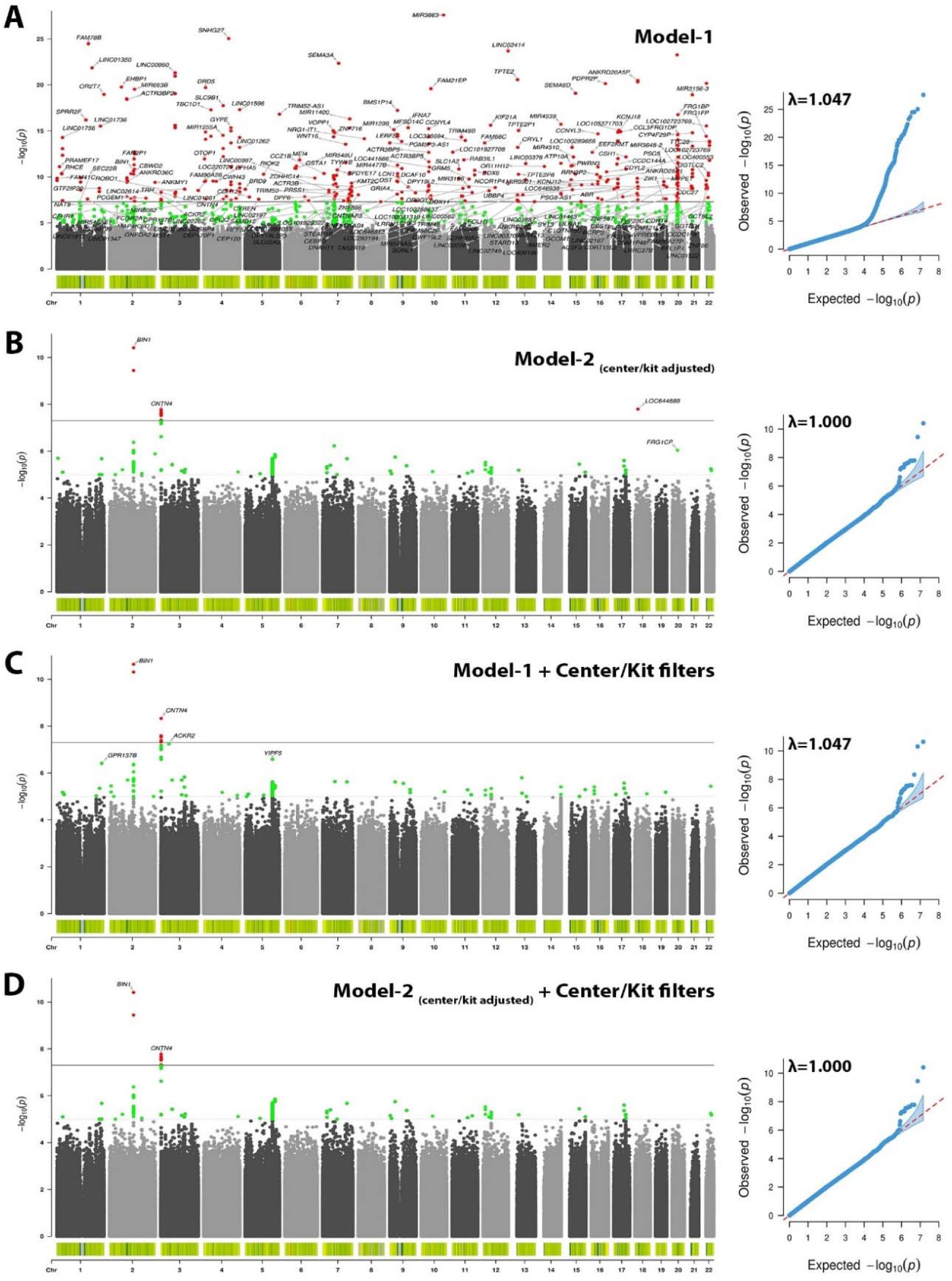
The proposed center/kit-based variant filters remove spurious associations in ADSP WGS. Figure shows Manhattan (left) and quantile-quantile (right) plots. **A)** Model-1 indicates many spurious hits. **B)** Model-2 shows that adjustment for center/kit can reduce many, but not all, spurious hits. **C)** Filters remove most spurious hits. **D)** Further adjustment for center/kit removes few additional spurious hits.

Closer inspection of the center/kit-based variant filters showed that non-filtered variants displayed fairly concordant P-values across model-1 and model-2, whereas filtered variants showed many discrepancies (**Figure 4A-B & 4D-E**). This was consistent with the filtered variants driving spurious associations. Additionally, it was apparent that filters removed variants across the full frequency range (**Figure 4C & 4F**) consistent with the increased MAF variation across all frequency ranges for variants underlying spurious association signals (**Figure 1A-B**).

**Figure 4.**
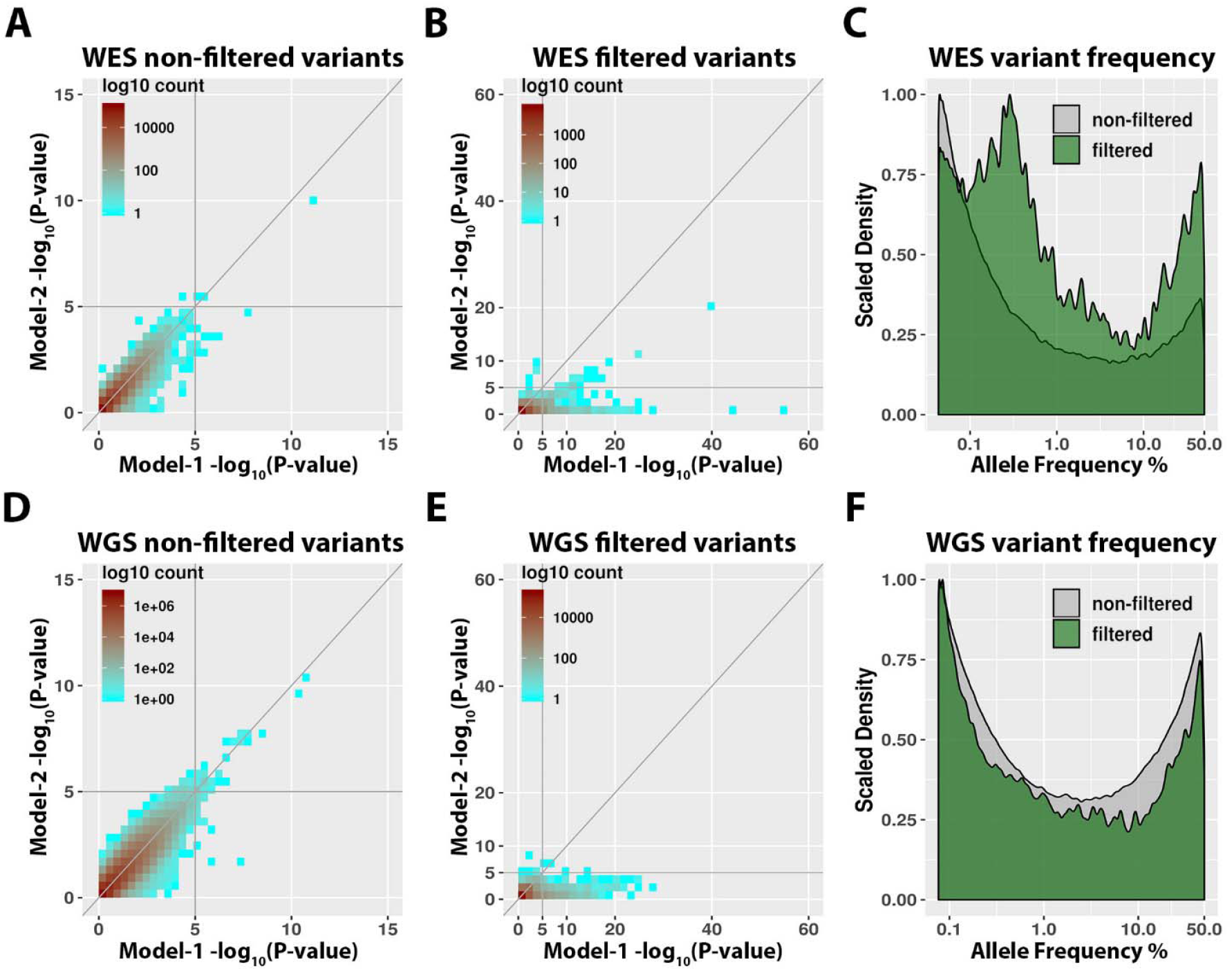
Metrics of variants removed by the proposed center/kit-based variant filters. **A-C)** ADSP WES. **D-E)** ADSP WGS. **A & D)** Variants that passed filters showed largely consistent P-values across model-1 and model-2 case-control association analyses, with only few variants remaining that reach suggestive significance in model-1 but lose suggestive significance upon center/kit adjustment in model-2 (lower right quadrant). **B & E)** Variants that were removed by filters showed many inconsistent P-values across model-1 and model-2, consistent with center/kit related variant artifacts that could not fully be accounted for by model-2. **C & F)** Frequency density plots, comparing variants that were filtered/removed to those that were not filtered. Note that variants were consistently filtered across the full frequency range, with increased density at frequencies <1% or >10% in ADSP WES.

We then assessed to what extent the gnomAD-based filters could remove the observed spurious associations. Visual assessment of Manhattan plots showed that the gnomAD-based filters could only account for a part of the spurious associations (**Figure S8-9**). Similarly, closer inspection of the gnomAD-based filters showed that they mainly removed variants with frequencies <1% (**Figure S10**). P-values across model-1 and model-2 further showed many discrepancies both for non-filtered and filtered variants (although fewer for non-filtered variants). In sum, the gnomAD-based filters could remove some spurious signals, but were less effective than the center/kit-based variant filters.

We further assessed to what extent the duplicate discordant variants filters could remove the observed spurious associations. Manhattan plots showed that the duplicate discordant variant filters could account for many of the spurious associations, but several remained when using model-1, while when using model-2 the Manhattan plots looked similar to those using the center-kit-based variant filters (**Figure S11-12**). Closer inspection of the duplicate discordant variant filters similarly showed they mainly removed variants with frequencies >10% and did not remove a set of variants that lose suggestive significance when going from model-1 to model-2 (**Figure S13**). An illustrative example of such a variant is provided in **Table S7**, confirming these variants represent genotyping issues that more ideally should be removed from the data. In sum, the duplicate discordant filters could remove many spurious signals, but were less effective than the center/kit-based variant filters, yet more effective than the gnomAD-based variant filters.

We also sought to understand the overlap between the different proposed filters. The three gnomAD-based variant filters appeared to show little overlap with one another (**Figure S14**) and overlapped with less than 20% of the variants in the center/kit-based variant filters (**Figure S15**). Further, in ADSP WES and WGS, respectively 32% and 14% of duplicate discordant variants overlapped center/kit-based variant filters, while vice versa 31% and 15% of center/kit-based filtered variants overlapped duplicate discordant variants (**Figure S16**). In the same comparison, respectively 28% and 49% of duplicate discordant variants overlapped gnomAD-based variant filters, while vice versa 53% and 17% of gnomAD-based filtered variants overlapped duplicate discordant variants (**Figure S17**). In sum, this confirmed that all three types of filters captured overlapping as well as unique variants. Notably, the center/kit- and gnomAD-based variant filters could capture a subset of reference artifactual variants present in the duplicate discordant variant filters, but identified many additional signals that represented likely artifactual variants and that contributed to spurious association signals.

Then, we sought to assess whether the use of these different types of variant filters could omit the need for adjusting for sequencing center/kit as implemented in model-2, which may be desirable for certain studies or research questions. We thus inspected all variants that passed suggestive significance in either model-1 or model-2 in ADSP WES (**Table 2**) and WGS (**Table S8**) after applying the center/kit-based filters (which we showed removed the most spurious signals). We observed that many variants that lose suggestive significance after center/kit adjustment in model-2 have fairly small (above threshold) P-values in the center/kit-based Fisher exact tests and/or are covered in the gnomAD-based and duplicate discordant variant filters. Similarly, assessing Manhattan plots and variant metrics suggested that the gnomAD-based and/or duplicate discordant variant filters removed few additional variants underlying spurious signals (**Figure S18-23**). This suggests there may be added value in using model-2 and/or applying the gnomAD-based filters to reduce spurious signals. Obviously, adding the duplicate discordant variant filters will inherently remove artifactual signals and help reduce spurious signals.

**Table 2.**
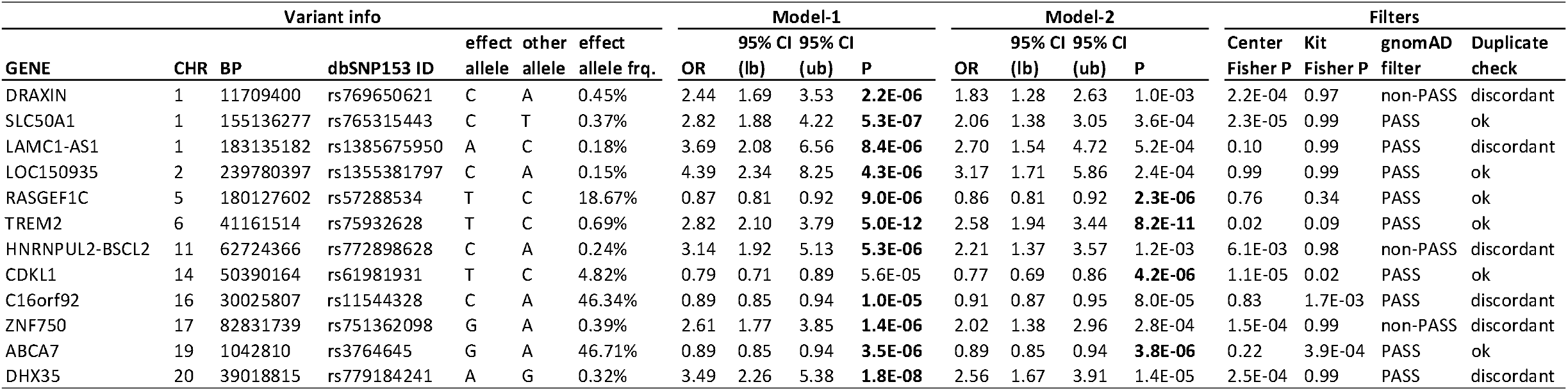
ADSP WES variants passing suggestive significance after applying centers/kit-based filters. Variants shown passed suggestive significance in either model-1 or model-2. Note that many variants that lose suggestive significance after center/kit adjustment in model-2 have fairly small P-values (but above threshold) in the center/kit Fisher tests and/or have a non-PASS flag in gnomAD or are flagged by the duplicate discordant variant filter. This suggests there is added value in using model 2 and/or applying the gnomAD and duplicate discordant variant filters to reduce spurious signals, or, that model 1 without gnomAD filters can be used contingent on post-hoc assessment of the association signal’s robustness.

Lastly, as a robustness check, we compared association statistics from the current ADSP WES analyses to variants that we identified in a prior study using a prior version of the ADSP WES data and observed highly concordant findings (**Table S9**)^15^.

## Discussion

We present a novel, fast, and robust approach to filter variants that represent sequencing center- or kit-related artifacts underlying spurious associations with AD risk in ADSP WES and WGS data, that cannot fully be accounted for by center/kit covariate adjustment. In addition, we show that filters comprising variants that may be prone to artifacts as identified by gnomAD were less efficient in removing spurious signals, but may still have added value on top of the center/kit-based filters. Similarly, filters containing variants that were discordant across duplicate samples could remove many, but not all, spurious signals, and added onto the center/kit-based filters. In sum, the presented filters are important to support future robust studies on ADSP data. In addition, these filters allow flexibility given that they can be applied in post-hoc quality control. Researchers may thus inspect filtered variants in targeted analyses in subsets of the ADSP data where no artifactual genotype enrichment is observed (e.g. excluding a single sequencing center/kit that showed an artifactual increase in genotype counts compared to the others).

Certain study designs or research questions may benefit from not adjusting by sequencing center/kit (i.e. cohort adjustment). For example, a study that considers specific strata and/or low frequency variants may observe some co-linearity between variant genotype observations and sequencing centers/kits. However, this does not necessarily indicate artifactual variants and may be driven by chance or variable cohort study designs across samples sequenced by different centers. We observed that the presented center/kit-based variant filters could handle nearly all spurious associations when not adjusting for sequencing center/kit in model-1. Inspecting the remaining signals passing suggestive significance, it was apparent that the gnomAD-based and duplicate discordant variant filters could remove a few additional spurious signals. Similarly, the P-values from the Fisher exact tests across sequencing centers/kits was fairly small for several variants that passed suggestive significance in their association with AD risk in model-1 but lost suggestive significance upon center/kit adjustment in model-2. In sum, we suggest that model-2 with application of center/kit-based, gnomAD-based, and duplicate discordant variant filters is the most conservative approach, but model-1 using only center/kit-based and duplicate discordant variant filters may reasonably be implemented, contingent on post-hoc assessment of the association signals’ robustness.

The center/kit-based filtering approach will further be valuable beyond the currently presented exome- and genome-wide univariate AD risk association analyses in European ancestry samples. Notably, removal of artifactual variants may lead to improved association statistics in gene-based testing, which is particularly relevant for WES/WGS data^7^. The filter approach can also be applied to non-European samples available in ADSP WES/WGS. Lastly, the approach to check for variant artifacts by comparing genotype distributions across sequencing centers/kits may also be used in other studies with a similar design as the current ADSP data. Notably, pre-processing of UK Biobank SNP array data has already implemented a similar type of filter as the one we described here in order to remove variants that may represent batch or array effects^28^. In turn, the approach described here and applied to WES/WGS data could also be applied to the large amount of SNP array data sets used in large-scale genetic studies of AD^3^.

The current study is the first to report exome- and genome-wide AD risk association findings for the newly released ADSP 20.5k WES and 16.9k WGS data. After quality control and filter implementation, we observed few signals passing the genome-wide significance threshold. In the ADSP WES data, *TREM2* and *ABCA7*—well-established AD risk genes^2,6^—were observed with variants respectively at genome-wide and suggestive significance, consistent with observations for similar models in prior studies on the prior ADSP WES discovery phase data^7,15^. Despite only observing 4 variants in ADSP WES that passed suggestive significance in model-2, our findings were overall highly consistent with prior work^15^. We also observed that certain variants identified previously were not present in our current summary statistics (**Table S9**), reflecting differences in joint calling, quality control, and the fact that currently only bi-allelic data were available for the new ADSP WES data. Notably, the common protective variant on *ABCA7* identified here has not been previously reported (and we confirm it appears to not have been successfully joint called in the prior ADSP WES data; dbGaP accession ID: phs000572). In the ADSP WGS data, in addition to several suggestive hits, *BIN1*—a well-established AD risk gene^2,6^—and *CNTN4* were identified with variants at genome-wide significance. The common protective variant on *CNTN4* appears novel and may be of relevance to AD pathogenesis given that Contactin 4 (CNTN4) is a binding partner of Amyloid Precursor Protein (APP) and CNTN4/APP interaction may play a role in promoting target-specific axon arborization^29,30^. Overall, these initial findings appear promising but suggest that the current ADSP WES/WGS data may still suffer from power limitations limiting discovery of novel risk variants. As such, gene-based testing, analyses on available non-European ancestry samples, and novel methodological approaches to gain additional power^12,15^, will all be crucial to support future advances into disentangling the missing heritability of AD using ADSP samples and other complimentary large-scale sequencing data.

## Conclusion

We present a novel, fast, and robust approach to filter variants that represent sequencing center- or kit-related artifacts underlying spurious associations with AD risk in ADSP WES and WGS data. This approach will be important to support future robust studies on ADSP data, as well as other studies with similar designs.

## Data Availability

All data used in the discovery analyses are available upon application to:
- dbGaP (https://www.ncbi.nlm.nih.gov/gap/)
- NIAGADS (https://www.niagads.org/)
- LONI (https://ida.loni.usc.edu/)
- Synapse (https://www.synapse.org/)
- Rush (https://www.radc.rush.edu/)
- NACC (https://naccdata.org/)

https://www.niagads.org/

https://ida.loni.usc.edu/

https://www.synapse.org/

https://www.radc.rush.edu/

https://naccdata.org/

## Contributions

M.E.B. performed data processing, performed data analyses, designed analyses, designed study, wrote paper, and obtained funding. Y.L.G. performed data processing and designed analyses. S.J.E. performed data processing. V.N. performed data processing and supervised work. M.D.G supervised analyses, supervised work, and obtained funding. Z.H designed study, designed analyses, supervised analyses, supervised work, wrote paper, and obtained funding.

## Data sharing statement

All data used in the analyses are available upon application to:

‐ dbGaP (https://www.ncbi.nlm.nih.gov/gap/)
‐ NIAGADS (https://www.niagads.org/)
‐ LONI (https://ida.loni.usc.edu/)
‐ Synapse (https://www.synapse.org/)
‐ Rush (https://www.radc.rush.edu/)
‐ NACC (https://naccdata.org/)

The specific data repository and identifier for each cohort is indicated in **Table S1-2** of the supplement.

Summary statistics from the current study will be available at https://www.niagads.org/home/

## Acknowledgements

Funding for this study was provided by the The Iqbal Farrukh & Asad Jamal Fund, the NIH (AG060747 and AG047366, granted to M.D.G, AG066206 and AG066515 granted to Z.H), and the Alzheimer’s Association (AARF-20-683984, granted to M.E.B), the European Union’s Horizon 2020 research and innovation program under the Marie Skłodowska-Curie (grant agreement No. 890650, granted to Y.L.G).

Biological samples used in this study were stored at study investigators’ institutions and at the National Cell Repository for Alzheimer’s Disease (NCRAD) at Indiana University, which receives government support under a cooperative agreement grant (U24 AG21886) awarded by the National Institute on Aging (NIA). We thank contributors who collected samples used in this study, as well as patients and their families, whose help and participation made this work possible. Phenotypic data were provided by principal investigators, the NIA funded Alzheimer’s Disease Centers (ADCs), the National Alzheimer’s Coordinating Center (NACC, U01AG016976), and the National Institute on Aging Genetics of Alzheimer’s Disease Data Storage Site (NIAGADS, U24AG041689) at the University of Pennsylvania, funded by NIA. Contributors to the Genetic Analysis Data included Study Investigators on projects that were individually funded by NIA, and other NIH institutes, and by private U.S. organizations, or foreign governmental or nongovernmental organizations.

Data for this study were prepared, archived, and distributed by the National Institute on Aging Alzheimer’s Disease Data Storage Site (NIAGADS) at the University of Pennsylvania (U24-AG041689-01); Alzheimer’s Disease Genetics Consortium (ADGC), U01 AG032984, RC2 AG036528; NACC, U01 AG016976; NIA-LOAD (Columbia University), U24 AG026395, U24 AG026390, R01AG041797; Banner Sun Health Research Institute P30 AG019610; Boston University, P30 AG013846, U01 AG10483, R01 CA129769, R01 MH080295, R01 AG017173, R01 AG025259, R01 AG048927, R01AG33193, R01 AG009029; Columbia University, P50 AG008702, R37 AG015473, R01 AG037212, R01 AG028786; Duke University, P30 AG028377, AG05128; Emory University, AG025688; Group Health Research Institute, UO1 AG006781, UO1 HG004610, UO1 HG006375, U01 HG008657; Indiana University, P30 AG10133, R01 AG009956, RC2 AG036650; Johns Hopkins University, P50 AG005146, R01 AG020688; Massachusetts General Hospital, P50 AG005134; Mayo Clinic, P50 AG016574, R01 AG032990, KL2 RR024151; Mount Sinai School of Medicine, P50 AG005138, P01 AG002219; New York University, P30 AG08051, UL1 RR029893, 5R01AG012101, 5R01AG022374, 5R01AG013616, 1RC2AG036502, 1R01AG035137; North Carolina A&T University, P20 MD000546, R01 AG28786-01A1; Northwestern University, P30 AG013854; Oregon Health & Science University, P30 AG008017, R01 AG026916; Rush University, P30 AG010161, R01 AG019085, R01 AG15819, R01 AG17917, R01 AG030146, R01 AG01101, RC2 AG036650, R01 AG22018; TGEN, R01 NS059873; University of Alabama at Birmingham, P50 AG016582, UL1RR02777; University of Arizona, R01 AG031581; University of California, Davis, P30 AG010129; University of California, Irvine, P50 AG016573, P50 AG016575, P50 AG016576, P50 AG016577; University of California, Los Angeles, P50 AG016570; University of California, San Diego, P50 AG005131; University of California, San Francisco, P50 AG023501, P01 AG019724; University of Kentucky, P30 AG028383, AG05144; University of Michigan, P30 AG053760 and AG063760; University of Pennsylvania, P30 AG010124; University of Pittsburgh, P50 AG005133, AG030653, AG041718, AG07562, AG02365; University of Southern California, P50 AG005142; University of Texas Southwestern, P30 AG012300; University of Miami, R01 AG027944, AG010491, AG027944, AG021547, AG019757; University of Washington, P50 AG005136, R01 AG042437; University of Wisconsin, P50 AG033514; Vanderbilt University, R01 AG019085; and Washington University, P50 AG005681, P01 AG03991, P01 AG026276. The Kathleen Price Bryan Brain Bank at Duke University Medical Center is funded by NINDS grant # NS39764, NIMH MH60451 and by Glaxo Smith Kline. Genotyping of the TGEN2 cohort was supported by Kronos Science. The TGen series was also funded by NIA grant AG041232, The Banner Alzheimer’s Foundation, The Johnnie B. Byrd Sr. Alzheimer’s Institute, the Medical Research Council, and the state of Arizona and also includes samples from the following sites: Newcastle Brain Tissue Resource (funding via the Medical Research Council, local NHS trusts and Newcastle University), MRC London Brain Bank for Neurodegenerative Diseases (funding via the Medical Research Council), South West Dementia Brain Bank (funding via numerous sources including the Higher Education Funding Council for England (HEFCE), Alzheimer’s Research Trust (ART), BRACE as well as North Bristol NHS Trust Research and Innovation 58 Department and DeNDRoN), The Netherlands Brain Bank (funding via numerous sources including Stichting MS Research, Brain Net Europe, Hersenstichting Nederland Breinbrekend Werk, International Parkinson Fonds, Internationale Stiching Alzheimer Onderzoek), Institut de Neuropatologia, Servei Anatomia Patologica, Universitat de Barcelona.

The NACC database is funded by NIA/NIH Grant U01 AG016976. NACC data are contributed by the NIA-funded ADCs: P30 AG019610 (PI Eric Reiman, MD), P30 AG013846 (PI Neil Kowall, MD), P30 AG062428-01 (PI James Leverenz, MD) P50 AG008702 (PI Scott Small, MD), P50 AG025688 (PI Allan Levey, MD, PhD), P50 AG047266 (PI Todd Golde, MD, PhD), P30 AG010133 (PI Andrew Saykin, PsyD), P50 AG005146 (PI Marilyn Albert, PhD), P30 AG062421-01 (PI Bradley Hyman, MD, PhD), P30 AG062422-01 (PI Ronald Petersen, MD, PhD), P50 AG005138 (PI Mary Sano, PhD), P30 AG008051 (PI Thomas Wisniewski, MD), P30 AG013854 (PI Robert Vassar, PhD), P30 AG008017 (PI Jeffrey Kaye, MD), P30 AG010161 (PI David Bennett, MD), P50 AG047366 (PI Victor Henderson, MD, MS), P30 AG010129 (PI Charles DeCarli, MD), P50 AG016573 (PI Frank LaFerla, PhD), P30 AG062429-01(PI James Brewer, MD, PhD), P50 AG023501 (PI Bruce Miller, MD), P30 AG035982 (PI Russell Swerdlow, MD), P30 AG028383 (PI Linda Van Eldik, PhD), P30 AG053760 (PI Henry Paulson, MD, PhD), P30 AG010124 (PI John Trojanowski, MD, PhD), P50 AG005133 (PI Oscar Lopez, MD), P50 AG005142 (PI Helena Chui, MD), P30 AG012300 (PI Roger Rosenberg, MD), P30 AG049638 (PI Suzanne Craft, PhD), P50 AG005136 (PI Thomas Grabowski, MD), P30 AG062715-01 (PI Sanjay Asthana, MD, FRCP), P50 AG005681 (PI John Morris, MD), P50 AG047270 (PI Stephen Strittmatter, MD, PhD).

The genotypic and associated phenotypic data used in the study “Multi-Site Collaborative Study for Genotype-Phenotype Associations in Alzheimer’s Disease (GenADA)” were provided by the GlaxoSmithKline, R&D Limited.

ROSMAP study data were provided by the Rush Alzheimer’s Disease Center, Rush University Medical Center, Chicago. Data collection was supported through funding by NIA grants P30AG10161, R01AG15819, R01AG17917, R01AG30146, R01AG36836, U01AG32984, U01AG46152, the Illinois Department of Public Health, and the Translational Genomics Research Institute.

The AddNeuroMed data are from a public-private partnership supported by EFPIA companies and SMEs as part of InnoMed (Innovative Medicines in Europe), an Integrated Project funded by the European Union of the Sixth Framework program priority FP6-2004-LIFESCIHEALTH-5. Clinical leads responsible for data collection are Iwona Kłoszewska (Lodz), Simon Lovestone (London), Patrizia Mecocci (Perugia), Hilkka Soininen (Kuopio), Magda Tsolaki (Thessaloniki), and Bruno Vellas (Toulouse), imaging leads are Andy Simmons (London), Lars-Olad Wahlund (Stockholm) and Christian Spenger (Zurich) and bioinformatics leads are Richard Dobson (London) and Stephen Newhouse (London).

Data collection and sharing for this project was funded by the Alzheimer’s Disease Neuroimaging Initiative (ADNI) (National Institutes of Health Grant U01 AG024904) and DOD ADNI (Department of Defense award number W81XWH-12-2-0012). ADNI is funded by the National Institute on Aging, the National Institute of Biomedical Imaging and Bioengineering and through generous contributions from the following: AbbVie. Alzheimer’s Association; Alzheimer’s Drug Discovery Foundation; Araclon Biotech; BioClinica. Inc.; Biogen; Bristol-Myers Squibb Company; CereSpir. Inc.; Cogstate; Eisai Inc.; Elan Pharmaceuticals. Inc.; Eli Lilly and Company; EuroImmun; F. Hoffmann-La Roche Ltd and its affiliated company Genentech. Inc.; Fujirebio; GE HealtControlsare; IXICO Ltd.; Janssen Alzheimer Immunotherapy Research & Development. LLC.; Johnson & Johnson Pharmaceutical Research & Development LLC.; Lumosity; Lundbeck; Merck & Co. Inc.; Meso Scale Diagnostics. LLC.; NeuroRx Research; Neurotrack Technologies; Novartis Pharmaceuticals Corporation; Pfizer Inc.; Piramal Imaging; Servier; Takeda Pharmaceutical Company; and Transition Therapeutics. The Canadian Institutes of Health Research is providing funds to support ADNI clinical sites in Canada. Private sector contributions are facilitated by the Foundation for the National Institutes of Health. The grantee organization is the Northern California Institute for Research and Education, and the study is coordinated by the Alzheimer’s Therapeutic Research Institute at the University of Southern California. ADNI data are disseminated by the Laboratory for Neuro Imaging at the University of Southern California.

The Alzheimer’s Disease Sequencing Project (ADSP) is comprised of two Alzheimer’s Disease (AD) genetics consortia and three National Human Genome Research Institute (NHGRI) funded Large Scale Sequencing and Analysis Centers (LSAC). The two AD genetics consortia are the Alzheimer’s Disease Genetics Consortium (ADGC) funded by NIA (U01 AG032984), and the Cohorts for Heart and Aging Research in Genomic Epidemiology (CHARGE) funded by NIA (R01 AG033193), the National Heart, Lung, and Blood Institute (NHLBI), other National Institute of Health (NIH) institutes and other foreign governmental and non-governmental organizations. The Discovery Phase analysis of sequence data is supported through UF1AG047133 (to Drs. Schellenberg, Farrer, Pericak-Vance, Mayeux, and Haines); U01AG049505 to Dr. Seshadri; U01AG049506 to Dr. Boerwinkle; U01AG049507 to Dr. Wijsman; and U01AG049508 to Dr. Goate and the Discovery Extension Phase analysis is supported through U01AG052411 to Dr. Goate, U01AG052410 to Dr. Pericak-Vance and U01 AG052409 to Drs. Seshadri and Fornage.

The ADGC cohorts included in ADSP include: Adult Changes in Thought (ACT) (UO1 AG006781, UO1 HG004610, UO1 HG006375, U01 HG008657), the Alzheimer’s Disease Centers (ADC) (P30 AG019610, P30 AG013846, P50 AG008702, P50 AG025688, P50 AG047266, P30 AG010133, P50 AG005146, P50 AG005134, P50 AG016574, P50 AG005138, P30 AG008051, P30 AG013854, P30 AG008017, P30 AG010161, P50 AG047366, P30 AG010129, P50 AG016573, P50 AG016570, P50 AG005131, P50 AG023501, P30 AG035982, P30 AG028383, P30 AG010124, P50 AG005133, P50 AG005142, P30 AG012300, P50 AG005136, P50 AG033514, P50 AG005681, and P50 AG047270), the Chicago Health and Aging Project (CHAP) (R01 AG11101, RC4 AG039085, K23 AG030944), Indianapolis Ibadan (R01 AG009956, P30 AG010133), the Memory and Aging Project (MAP) (R01 AG17917), Mayo Clinic (MAYO) (R01 AG032990, U01 AG046139, R01 NS080820, RF1 AG051504, P50 AG016574), Mayo Parkinson’s Disease controls (NS039764, NS071674, 5RC2HG005605), University of Miami (R01 AG027944, R01 AG028786, R01 AG019085, IIRG09133827, A2011048), the Multi-Institutional Research in Alzheimer’s Genetic Epidemiology Study (MIRAGE) (R01 AG09029, R01 AG025259), the National Cell Repository for Alzheimer’s Disease (NCRAD) (U24 AG21886), the National Institute on Aging Late Onset Alzheimer’s Disease Family Study (NIA-LOAD) (R01 AG041797), the Religious Orders Study (ROS) (P30 AG10161, R01 AG15819), the Texas Alzheimer’s Research and Care Consortium (TARCC) (funded by the Darrell K Royal Texas Alzheimer’s Initiative), Vanderbilt University/Case Western Reserve University (VAN/CWRU) (R01 AG019757, R01 AG021547, R01 AG027944, R01 AG028786, P01 NS026630, and Alzheimer’s Association), the Washington Heights-Inwood Columbia Aging Project (WHICAP) (RF1 AG054023), the University of Washington Families (VA Research Merit Grant, NIA: P50AG005136, R01AG041797, NINDS: R01NS069719), the Columbia University Hispanic Estudio Familiar de Influencia Genetica de Alzheimer (EFIGA) (RF1 AG015473), the University of Toronto (UT) (funded by Wellcome Trust, Medical Research Council, Canadian Institutes of Health Research), and Genetic Differences (GD) (R01 AG007584). The CHARGE cohorts are supported in part by National Heart, Lung, and Blood Institute (NHLBI) infrastructure grant HL105756 (Psaty), RC2HL102419 (Boerwinkle) and the neurology working group is supported by the National Institute on Aging (NIA) R01 grant AG033193.

The CHARGE cohorts participating in the ADSP include the following: Austrian Stroke Prevention Study (ASPS), ASPS-Family study, and the Prospective Dementia Registry-Austria (ASPS/PRODEM-Aus), the Atherosclerosis Risk in Communities (ARIC) Study, the Cardiovascular Health Study (CHS), the Erasmus Rucphen Family Study (ERF), the Framingham Heart Study (FHS), and the Rotterdam Study (RS). ASPS is funded by the Austrian Science Fond (FWF) grant number P20545-P05 and P13180 and the Medical University of Graz. The ASPS-Fam is funded by the Austrian Science Fund (FWF) project I904), the EU Joint Programme - Neurodegenerative Disease Research (JPND) in frame of the BRIDGET project (Austria, Ministry of Science) and the Medical University of Graz and the Steiermärkische Krankenanstalten Gesellschaft. PRODEM-Austria is supported by the Austrian Research Promotion agency (FFG) (Project No. 827462) and by the Austrian National Bank (Anniversary Fund, project 15435. ARIC research is carried out as a collaborative study supported by NHLBI contracts (HHSN268201100005C, HHSN268201100006C, HHSN268201100007C, HHSN268201100008C, HHSN268201100009C, HHSN268201100010C, HHSN268201100011C, and HHSN268201100012C). Neurocognitive data in ARIC is collected by U01 2U01HL096812, 2U01HL096814, 2U01HL096899, 2U01HL096902, 2U01HL096917 from the NIH (NHLBI, NINDS, NIA and NIDCD), and with previous brain MRI examinations funded by R01-HL70825 from the NHLBI. CHS research was supported by contracts HHSN268201200036C, HHSN268200800007C, N01HC55222, N01HC85079, N01HC85080, N01HC85081, N01HC85082, N01HC85083, N01HC85086, and grants U01HL080295 and U01HL130114 from the NHLBI with additional contribution from the National Institute of Neurological Disorders and Stroke (NINDS). Additional support was provided by R01AG023629, R01AG15928, and R01AG20098 from the NIA. FHS research is supported by NHLBI contracts N01-HC-25195 and HHSN268201500001I. This study was also supported by additional grants from the NIA (R01s AG054076, AG049607 and AG033040 and NINDS (R01 NS017950). The ERF study as a part of EUROSPAN (European Special Populations Research Network) was supported by European Commission FP6 STRP grant number 018947 (LSHG-CT-2006-01947) and also received funding from the European Community’s Seventh Framework Programme (FP7/2007-2013)/grant agreement HEALTH-F4- 2007-201413 by the European Commission under the programme “Quality of Life and Management of the Living Resources” of 5th Framework Programme (no. QLG2-CT-2002- 01254). High-throughput analysis of the ERF data was supported by a joint grant from the Netherlands Organization for Scientific Research and the Russian Foundation for Basic Research (NWO-RFBR 047.017.043). The Rotterdam Study is funded by Erasmus Medical Center and Erasmus University, Rotterdam, the Netherlands Organization for Health Research and Development (ZonMw), the Research Institute for Diseases in the Elderly (RIDE), the Ministry of Education, Culture and Science, the Ministry for Health, Welfare and Sports, the European Commission (DG XII), and the municipality of Rotterdam. Genetic data sets are also supported by the Netherlands Organization of Scientific Research NWO Investments (175.010.2005.011, 911-03-012), the Genetic Laboratory of the Department of Internal Medicine, Erasmus MC, the Research Institute for Diseases in the Elderly (014-93-015; RIDE2), and the Netherlands Genomics Initiative (NGI)/Netherlands Organization for Scientific Research (NWO) Netherlands Consortium for Healthy Aging (NCHA), project 050-060-810. All studies are grateful to their participants, faculty and staff. The content of these manuscripts is solely the responsibility of the authors and does not necessarily represent the official views of the National Institutes of Health or the U.S. Department of Health and Human Services.

The FUS cohorts include: the Alzheimer’s Disease Centers (ADC) (P30 AG019610, P30 AG013846, P50 AG008702, P50 AG025688, P50 AG047266, P30 AG010133, P50 AG005146, P50 AG005134, P50 AG016574, P50 AG005138, P30 AG008051, P30 AG013854, P30 AG008017, P30 AG010161, P50 AG047366, P30 AG010129, P50 AG016573, P50 AG016570, P50 AG005131, P50 AG023501, P30 AG035982, P30 AG028383, P30 AG010124, P50 AG005133, P50 AG005142, P30 AG012300, P50 AG005136, P50 AG033514, P50 AG005681, and P50 AG047270), Alzheimer’s Disease Neuroimaging Initiative (ADNI) (U19AG024904), Amish Protective Variant Study (RF1AG058066), Cache County Study (R01AG11380, R01AG031272, R01AG21136, RF1AG054052), Case Western Reserve University Brain Bank (CWRUBB) (P50AG008012), Case Western Reserve University Rapid Decline (CWRURD) (RF1AG058267, NU38CK000480), CubanAmerican Alzheimer’s Disease Initiative (CuAADI) (3U01AG052410), Estudio Familiar de Influencia Genetica en Alzheimer (EFIGA) (5R37AG015473, RF1AG015473, R56AG051876), Genetic and Environmental Risk Factors for Alzheimer Disease Among African Americans Study (GenerAAtions) (2R01AG09029, R01AG025259, 2R01AG048927), Gwangju Alzheimer and Related Dementias Study (GARD) (U01AG062602), Hussman Institute for Human Genomics Brain Bank (HIHGBB) (R01AG027944, Alzheimer’s Association “Identification of Rare Variants in Alzheimer Disease”), Ibadan Study of Aging (IBADAN) (5R01AG009956), Mexican Health and Aging Study (MHAS) (R01AG018016), Multi-Institutional Research in Alzheimer’s Genetic Epidemiology (MIRAGE) (2R01AG09029, R01AG025259, 2R01AG048927), Northern Manhattan Study (NOMAS) (R01NS29993), Peru Alzheimer’s Disease Initiative (PeADI) (RF1AG054074), Puerto Rican 1066 (PR1066) (Wellcome Trust (GR066133/GR080002), European Research Council (340755)), Puerto Rican Alzheimer Disease Initiative (PRADI) (RF1AG054074), Reasons for Geographic and Racial Differences in Stroke (REGARDS) (U01NS041588), Research in African American Alzheimer Disease Initiative (REAAADI) (U01AG052410), Rush Alzheimer’s Disease Center (ROSMAP) (P30AG10161, R01AG15819, R01AG17919), University of Miami Brain Endowment Bank (MBB), and University of Miami/Case Western/North Carolina A&T African American (UM/CASE/NCAT) (U01AG052410, R01AG028786). The four LSACs are: the Human Genome Sequencing Center at the Baylor College of Medicine (U54 HG003273), the Broad Institute Genome Center (U54HG003067), The American Genome Center at the Uniformed Services University of the Health Sciences (U01AG057659), and the Washington University Genome Institute (U54HG003079).

