## Supplementary File for "A fast and robust strategy to remove variant level artifacts in Alzheimer’s Disease Sequencing Project data"

### Supplementary Material

### Supplemental Methods

In the current study, we used data from a variety of cohorts and sequencing projects related to AD[1-23]. While we only present analyses on ADSP data, all available genetic/phenotic data were jointly harmonized with the purpose of performing phenotype/covariate harmonization. Details are provided below.

#### Phenotype Ascertainment

*Cohorts and Phenotype Ascertainment*

Details on phenotype ascertainment are described elsewhere[1–6]. Briefly, all individuals with a diagnosis of AD met National Institute of Neurological and Communicative Disorders and Stroke/Alzheimer's Disease and Related Disorders Association (NINCDS-ADRDA) criteria for probable or possible late onset AD[7], or met Diagnosis and Statistical Manual of Mental Disorders IV-V (DSMIV-V) criteria[8–10], or had a clinical dementia rating (CDR® Dementia Staging Instrument[11]) > 0.5. Some cohorts verified AD diagnoses by means of neuropathology, using Braak staging[12], CERAD scoring[13], or National Institute on Aging Reagan (NIA-Reagan) 1997 criteria[14]. Cognitively normal subjects (controls) did not have AD according to the above clinical criteria for AD, did not have a diagnosis of MCI, and had a CDR of 0 and/or Mini-Mental State Examination (MMSE[15]) > 25. In the MIRAGE cohort, control status was evaluated through a Modified Telephone Interview of Cognitive Status score ≥ 86 (a telephone version of the MMSE)[16].

Further, the National Alzheimer's Coordinating Center (NACC), Rush University Religious Orders Study/Memory and Aging Project (ROSMAP), and Alzheimer's Disease Neuroimaging Initiative (ADNI), are longitudinal cohorts that provide detailed information regarding clinical status (control, MCI, demented) and presumed disease etiology at repeated examinations. Additionally, deceased subjects are assessed for neuropathology. Where possible, in NACC, a final diagnoses of MCI or possible/probable/definite AD was obtained using NIA Alzheimer's Association (NIA-AA) 2011 criteria[17,18]. In all three cohorts, AD diagnoses were verified by neuropathology as middle or high AD likelihood following NIA-Reagan 1997 criteria (moderate to frequent neuritic plaques and Braak stage III-VI)[14]. In concordance with the category "possible AD dementia with evidence of the AD pathophysiological process" from the NIA-AA 2011 criteria[17], we attributed possible AD diagnoses to subjects who met clinical criteria for non-AD dementia but also met AD neuropathological criteria. In concordance with the NIA-AA 2011/2012 framework[18,19], we also evaluated neuropathology in MCI subjects to verify presumed AD etiology (cf. page 5). Controls were not re-evaluated based on neuropathology data. Subjects that reverted from dementia to control status during longitudinal follow-up were excluded. Additional cohort-specific details are listed below.

*NACC*

Genotyping waves 1 through 7 from the Alzheimer's Disease Centers (ADC1-7) and a subset of the ADSP projects include subjects ascertained and evaluated by the clinical and neuropathological cores of 32 NIA-funded ADCs. NACC coordinates the collection of these phenotypes, implements diagnoses (cognitively normal, cognitively impaired but not MCI, MCI, demented; and presumed disease etiology) and then provides all data to researchers under the form of the Minimum Data Set (MDS), Uniform Data Set (UDS)[20–22], and Neuropathology data set (NP)[23]. The MDS represents an older subset of the NACC data and only contains cross-sectional data, while the more recent UDS provides longitudinal phenotypes and covariates. Since 2015, the UDS was updated to incorporate the NIA-AA 2011 criteria for MCI and AD[18,24]. In the current study, we used the UDS and NP for which data was collected between September 2005 and March 2021, to determine phenotypes for subjects in ADC1-7, ADSP WES/WGS, and ADGC Exome arrays.

Subjects that had a diagnosis of Down syndrome, central nervous system neoplasm, bipolar disorder, schizophrenia, alcohol-induced dementia, or substance-abuse-induced dementia, were excluded. Subjects carrying mutations of dominantly inherited AD or frontotemporal lobar degeneration (FTLD) were also excluded. Subjects with a final diagnosis of MCI or dementia, for which the etiology was unknown, not due to AD, or only secondary due to AD (and without AD neuropathological information), were excluded. Subjects with a final diagnosis of "cognitively impaired but not MCI", but having no other neurological disorder, were kept as controls, considering that this more consistently matched control criteria in many of the other cohorts considered in this study.

*ROSMAP*

In ROSMAP, subjects were diagnosed at each visit: as possible/probable AD according to NINCDS-ADRDA criteria[7]; as MCI when judged to have cognitive impairment but not meeting dementia criteria according to the clinician; or as control when there was no cognitive impairment or the subject did not meet dementia criteria[25,26]. At time of death, a final clinical diagnosis was made by an expert neurologist, followed by case conference consensus review (blinded to postmortem data)[27].

*ADNI*

In ADNI, subjects were diagnosed at regular visits: as possible/probable AD according to NINCDS-ADRDA criteria[7]; as MCI according to Petersen/Winblad criteria; or as control when not demented, not MCI, CDR = 0, and MMSE > 28. Neuropathology assessments followed the NACC NP framework.

*Phenotype Harmonization*

The available sample contained many subjects that were genotyped multiple times across different studies. This largely reflected efforts from the ADGC, ADSP, and AMP-AD, to perform next generation sequencing (NGS) on existing cohort samples for the purpose of rare variant discovery and AD gene prioritization. In other instances, participants were recruited in different studies at different times. Therefore, to handle potential duplicate discordance and phenotype heterogeneity, we implemented a cross-sample phenotype harmonization procedure aiming to standardize pathology-verified diagnoses where possible, share unique missing information across all duplicate entries of a given subject, resolve longitudinal changes in diagnosis, and flag subjects with unresolvable duplicate discordance for exclusion.

Duplicate samples were identified by determining genetic cryptic relatedness (cf. page 6-7 below), but for the purpose of sample cross-referencing did not include known identical twins in LOAD and ROSMAP samples. First, duplicate samples were flagged as discordant if their age-at-death information differed by more than 2 years or if pathology measures (Braak or neuritic plaque density) differed. Across all cohorts, where possible, AD diagnoses were verified by neuropathology as middle or high AD likelihood following NIA-Reagan 1997 criteria (moderate to frequent neuritic plaques and Braak stage III-VI)[14]. Additionally, when only either neuritic plaque or Braak information was available and in line with NIA-Reagan 1997 middle or high AD likelihood criteria, and/or the cohort/project demographics provided a diagnosis of definite AD, the subject was considered to have pathology-verified AD status. Cognitively normal (CN) subjects with evidence of AD pathology were kept as CN. Further, if at least one entry across duplicate samples indicated a diagnosis of Down syndrome, central nervous system neoplasm, bipolar disorder, schizophrenia, alcohol-induced dementia, substance-abuse-induced dementia, neurological (not including Parkinson's disease) or systemic disease despite being cognitively normal, or carrying mutations of dominantly inherited AD or frontotemporal lobar degeneration (FTLD), then all duplicate samples were marked as such and flagged for exclusion. Extending on the above, all genetic samples were checked for the presence of known pathogenic mutations on *APP, PSEN1, PSEN2* and *MAPT*, whereby carriers and their duplicate samples were flagged for exclusion.

Then, duplicate samples with differing age entries (i.e. longitudinal changes) were evaluated. Reversions from AD or dementia to MCI status, or from MCI to cognitively normal (CN) status, were permitted, but reversions from AD or non-AD dementia to CN status were flagged for exclusion. "Reversions" from AD to non-AD dementia status were permitted, unless pathology (cf. above) indicated the presence of AD pathology, thereby marking the subject as AD. Vice versa, "conversions" from non-AD dementia to AD status were permitted, unless pathology (cf. above) indicated no presence of AD pathology, thereby marking the subject as non-AD dementia. All other types of conversions were directly permitted. Then, duplicate samples for which the diagnoses at the oldest shared age entries differed, or for which diagnoses differed but age was consistent (i.e. apparent cross-sectional discordances), were evaluated. Discordances between AD and non-AD dementia status were resolved on the basis of pathology (cf. above) or flagged as discordant if no pathology data was available. Discordances between CN and AD status, or CN and non-AD dementia status, were resolved as respectively AD or non-AD dementia when those dementia diagnoses corresponded to a unique age-at-onset (of symptoms) without other available age information (i.e. indicating that a conversion likely occurred after the subject was lost to follow-up in the cohort that last observed a CN status), or, were flagged as discordant if duplicate entries shared the same age-at-examination and age-at-last-exam. Discordances between CN and MCI status, or MCI and AD status, or MCI and non-AD dementia status, were resolved as respectively MCI, AD, or non-AD dementia (i.e. keeping the most severe diagnosis).

Finally, once all clinical diagnostic and pathological data were unified across duplicate entries, pathological criteria were applied once more to obtain the final diagnoses. Where possible, AD diagnoses were verified by neuropathology as middle or high AD likelihood following NIA-Reagan 1997 criteria (moderate to frequent neuritic plaques and Braak stage III-VI)[14]. In concordance with the category "possible AD dementia with evidence of the AD pathophysiological process" from the NIA-AA 2011 criteria[17], we attributed possible AD diagnoses to subjects who met clinical criteria for non-AD dementia but also met AD neuropathological criteria. In concordance with the NIA-AA 2011/2012 framework[18,19], we also evaluated neuropathology in MCI subjects to verify presumed AD etiology and considered subjects as cases if AD pathology, following NIA-Reagan 1997 criteria (cf. above), was present (i.e. marking high likelihood of AD etiology). Controls were not re-evaluated based on neuropathology data.

Beyond cross-referencing clinical diagnostic and pathological data across subjects, other covariates were considered for cross-referencing or sharing in case of missingness across duplicate entries. These included age-at-onset of cognitive symptoms, age-at-examination providing clinical diagnosis, at-at-last exam, age-at-death, sex, race, ethnicity, *APOE* genotype provided from demographics, *APOE* genotype provided from whole-genome sequencing, and *APOE* genotype provided from whole-exome sequencing. Duplicate entries with discordant sex or race information were flagged for exclusion.

### Genetic Data Quality Control and Processing

#### *Genetic Data Harmonization and Standard Quality Control*

Genotypes were available from commercial high-density single-nucleotide polymorphism (SNP) genotyping microarrays (Illumina or Affymetrix), Whole-exome sequencing (WES), or Whole-genome sequencing (WGS) (**Table S1**). Genotype samples had their genetic variants lifted to hg19 using liftOver if not released in hg19[28]. Autosomal variants were extracted from the SNP array data and further processed in several stages. First, SNP array data were processed by the Genotype Harmonizer with CEU and TSI HapMap populations as the reference panel, to perform automatic strand alignment[29]. Then, multi-allelic SNPs, SNPs located on common copy number or segmental duplication regions, and duplicated or monomorphic SNPs, were removed. The list of multi-allelic SNPs or SNPs located on common copy number and segmental duplication regions was created using Tri-Typer[30]. The list of CNV and segmental duplication regions was curated from the Eichler lab (eichlerlab.gs.washington.edu/database.html)[31] and the gnomAD website (gnomad.broadinstitute.org/downloads)[32]. All respective genotype data sets were then iteratively merged with each other, applying strand flipping and variant ID updating as applicable, to ultimately obtain parsimonious data sets that could be merged for cross-sample relationship determination and principal component analyses (cf. below).

Genetic data were then further processed using Plink v1.9. The numbers of remaining samples after each quality control (QC) or processing step are listed in **Table S3-4**. For each sample platform, subjects with autosome missingness (≥ 5%) and sex problems (discordance between genetic sex and demographic sex, or deviation of expected X-chromosome homozygosity/heterozygosity) were flagged for exclusion.

#### *Ancestry Determination*

Individual ancestries were determined using SNPweights v.2.1 with populations from the 1000 Genomes Consortium as a reference[33,34]. By applying an ancestry percentage cut-off ≥ 75%, the samples were stratified into the five super populations, South-Asians (SAS), East-Asians (EAS), Americans (AMR), Africans (AFR) and Europeans (EUR) (**Figure S1**). Subjects with a genetic ancestry that differed from their race, as provided in cohort demographics, were flagged for exclusion. Subjects belonging to the European ancestry super population were further determined according to three major ancestries, that is, Northwestern European (NWE), Southeastern European (SEE), and Ashkenazi Jewish (AJE), using reference populations available from SNPweights v.2.1.2[33]. European subjects were stratified into the above-mentioned ethnicities by applying an ancestry percentage cut-off ≥ 50% (**Figure S1**).

*Genetic Duplicate Determination using Plink*

Across all cohorts the relatedness of subjects (after QC indicated above) was evaluated through identity-by-descent (IBD) analysis (using directly genotyped non-palindromic SNPs that shared across all genetic datasets with a call rate > 95%, minor allele frequency (MAF) > 1%). This outcome was used for duplicate (IBD>0.95) tracking across samples, which in turn was used to enable phenotype harmonization.

*Relationship Determination and Principal Component Analysis using GENESIS*

For ADSP WGS and WES data respectively, the relatedness of subjects and principal components capturing population substructure were determined using IBD and principal component analyses (PCA) as implemented through the R package GENESIS (R v3.6.0)[35]. Specifically, this approach first uses an R-implementation of KING-robust to determine kinship coefficients that take into account ancestry divergence. The derived pairwise kinship coefficients are then used to perform a PCA in related samples (PC-AiR) providing accurate ancestry inference not confounded by family structure. The latter output is then used to estimate kinship coefficients using PC-Relate, which accounts for population structure (ancestry) among sample individuals through the use of ancestry representative principal components (PCs) to provide accurate relatedness estimates due only to recent family (pedigree) structure. For each respective data set, these analyses were performed on pruned SNPs ($R^2 < 0.5$, call rate > 99.9%, MAF > 1%, and excluding palindromic SNPs) in non-Hispanic white European ancestry individuals (**Figure S2**).

*APOE genotype assessment in ADSP WES/WGS*

In ADSP WGS, the rs429358 and rs7412 variants showed low genotype missingness across subjects, reflecting good variant quality metrics. In ADSP WES, there was a high genotype missingness at rs7412 (32.5%). This resulted from a low read depth and genotype quality in some of the different WES capture kits that were used in the ADSP WES[2]. We therefore sought to re-call both variants in order to fill out missing *APOE* information where possible. We first inferred the variants' genotype using data called by the ADSP, which required a read depth read depth (DP) >=10 and genotype quality (GQ) >= 20. We then further inferred the variants' genotype if DP and GQ were respectively greater than or equal to 6 and 20, observing at least 20% alternate allele reads to call a heterozygote (e.g. *APOE**3/4).

After this first round of *APOE* genotype ascertainment, some individuals still had either the rs7412 or rs429358 genotype missing (i.e., only one of the two variants could be called using the above criteria), making it impossible to infer their *APOE* genotype from the ADSP NGS data alone. Many of these remaining individuals however had a reported *APOE* genotype in their demographics that could be used to complete the missing information in a second additional round of *APOE* genotype ascertainment. This approach was preferred over relying solely on the *APOE* genotype in the demographics, since the genotype calls on the ADSP NGS data are expected to provide higher accuracy compared to other commonly used *APOE* direct genotyping methods[36]. To illustrate, consider the example where one of these remaining individuals in the sequencing data was homozygous for the reference allele at rs429358, which would suggest the subject is *APOE**3/3, but had a missing genotype at rs7412. In this case, from the ADSP NGS data, we know that this individual is not carrying an *APOE**4 allele, but we cannot determine the presence or absence of an *APOE**2 allele. We then turned to the information from the *APOE* genotype provided in the demographics to infer the most likely *APOE* genotype. For the current example, if the individual has a provided *APOE* genotype that was 2/2, 2/3, or 3/3, then the information in the ADSP NGS data is deemed concordant with the provided *APOE* genotype (that is, rs429358 is always the reference allele for those provided *APOE* genotypes) and we used the provided *APOE* genotype. However, if the provided *APOE* genotype was 4/4 or 3/4, then we would correct it to *APOE**3/3, because the ADSP NGS information clearly indicated there was no *APOE**4 genotype call (similarly a provided *APOE**2/4 genotype would be corrected to *APOE**2/3). This can be generalized as: for remaining individuals with DP>=6 and GQ>=20 at rs429358, the ADSP NGS data at rs429358 was used to change, when discordant, the provided *APOE**3 genotype to *APOE**4, or vice-versa. One additional extension to this step was implemented for the few scenarios where the ADSP NGS data called two rs429358 alleles (i.e. *APOE**4/4) but the allelic distribution indicated that the reference allele was still observed (e.g. 1 REF allele and 7 ALT alleles). In these situations, if the provided *APOE* genotype indicated the presence of *APOE**3, then the genotype was corrected to *APOE**3/4 (reasoning there is sufficient evidence to support the presence of an *APOE**3 genotype). The extra checks described in this paragraph were also applied to subjects in the first QC round (prior paragraph), who had 6<=DP<10 and GQ>=20 for both rs429358 and rs7412.

As a quality check, using these thresholds, we did not observe any discordance in the inferred *APOE* genotype across 3,499 duplicates between the ADSP WGS and ADSP WES.

*ADSP WES/WGS quality control prior to genetic association testing*

After the genetic and phenotypic quality control/harmonization described in the above, the ADSP WES and WGS samples were respectively restricted to all non-Hispanic European ancestry individuals that pass filtering criteria (**Table S3-4**). The remaining samples underwent genetic quality control as detailed in the manuscript's method section, followed by association testing and construction of the genotype filters.

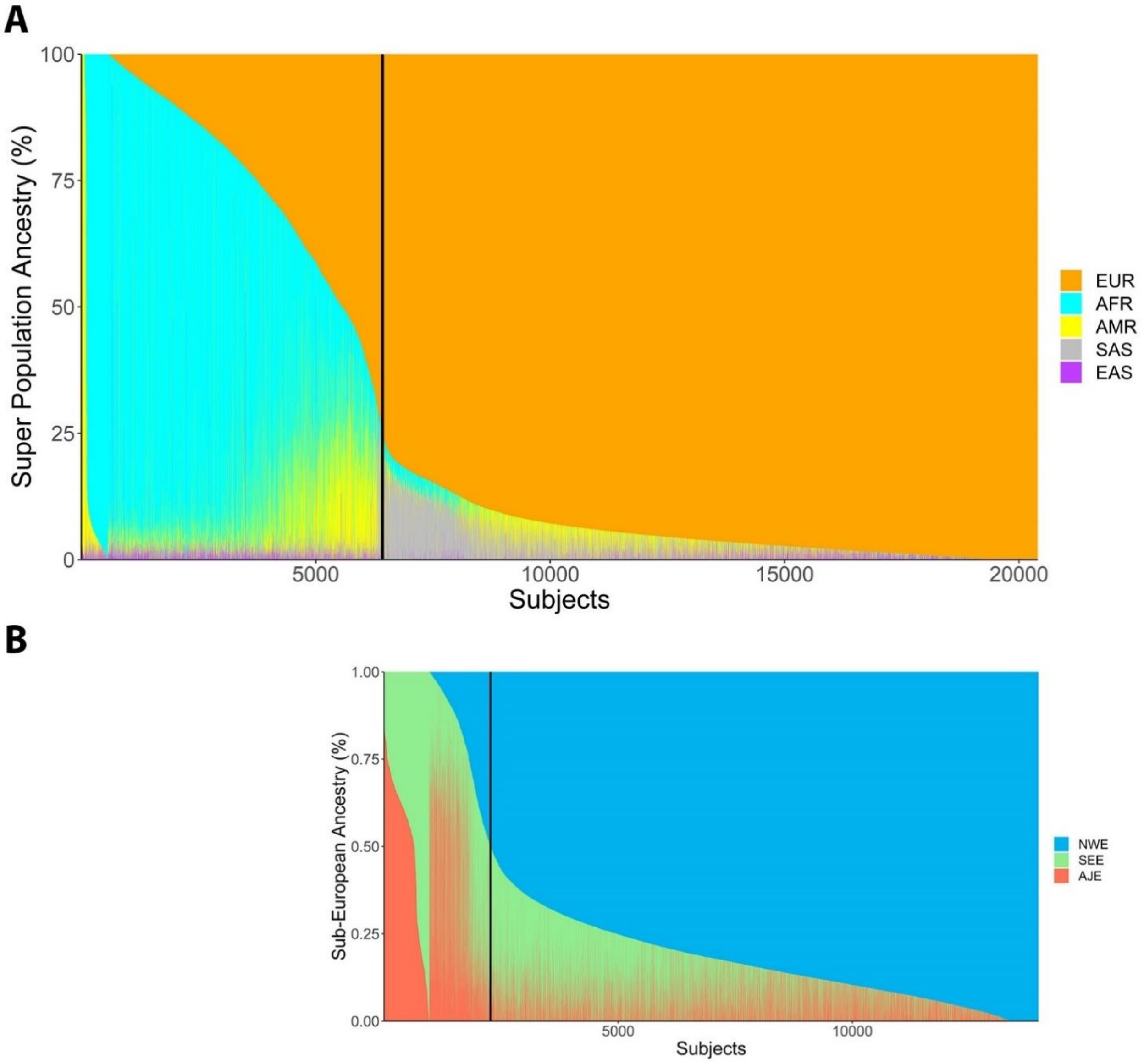

**Figure S1 (part 1). Admixture plot for the five major super populations across ADSP samples. (A-B) ADSP WES. A)** Black vertical line marks the cut-off for EUR ancestry [>=75%]. **B)** Black vertical line marks the cut-off for NWE ancestry [>=50%].

*Abbreviations: EUR, European; AFR, African; AMR; American; SAS, South Asian; EAS; East Asian; NWE, Northwestern European; SEE, Southeastern European; AJE, Ashkenazi Jewish.*

**C**

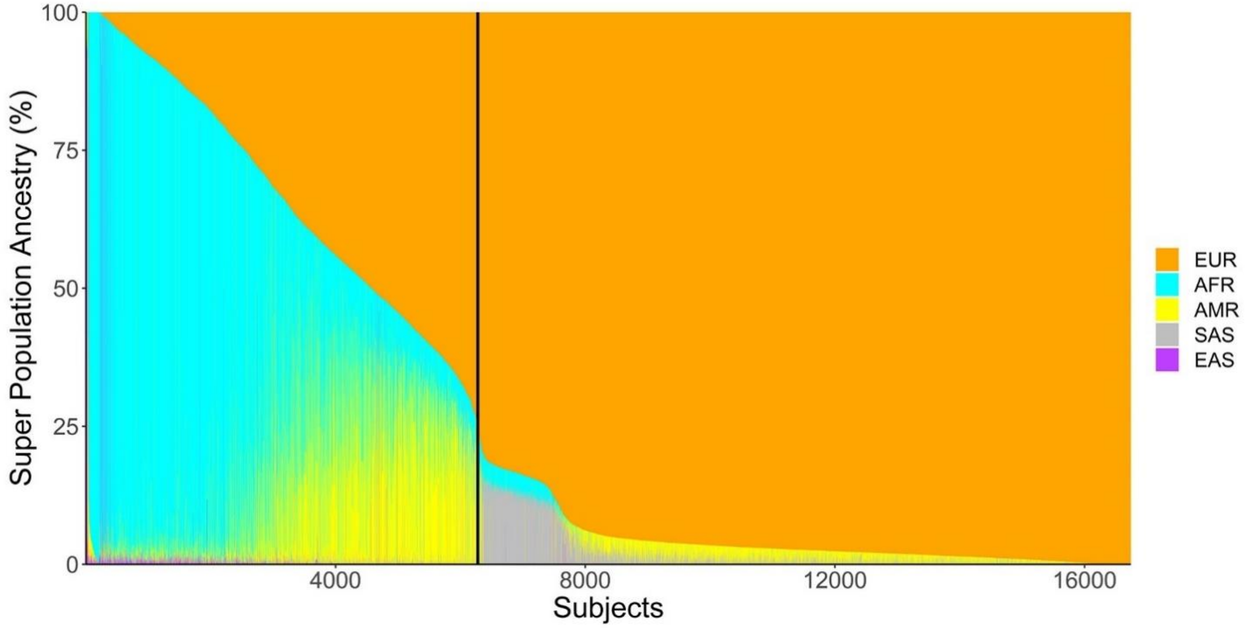

**D**

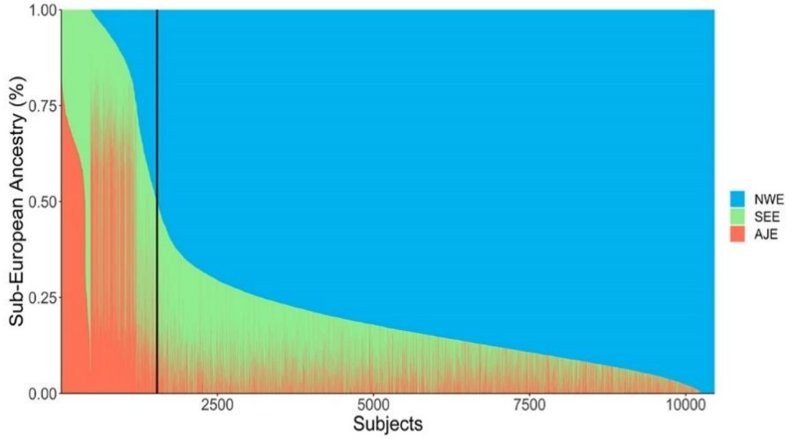

**Figure S1 (part 2). Admixture plot for the five major super populations across ADSP samples. (C-D) ADSP WGS. C)** Black vertical line marks the cut-off for EUR ancestry [>=75%]. **D)** Black vertical line marks the cut-off for NWE ancestry [>=50%].

*Abbreviations: EUR, European; AFR, African; AMR; American; SAS, Southern Asian; EAS; Eastern Asian; NWE, Northwestern European; SEE, Southeastern European; AJE, Ashkenazi Jewish.*

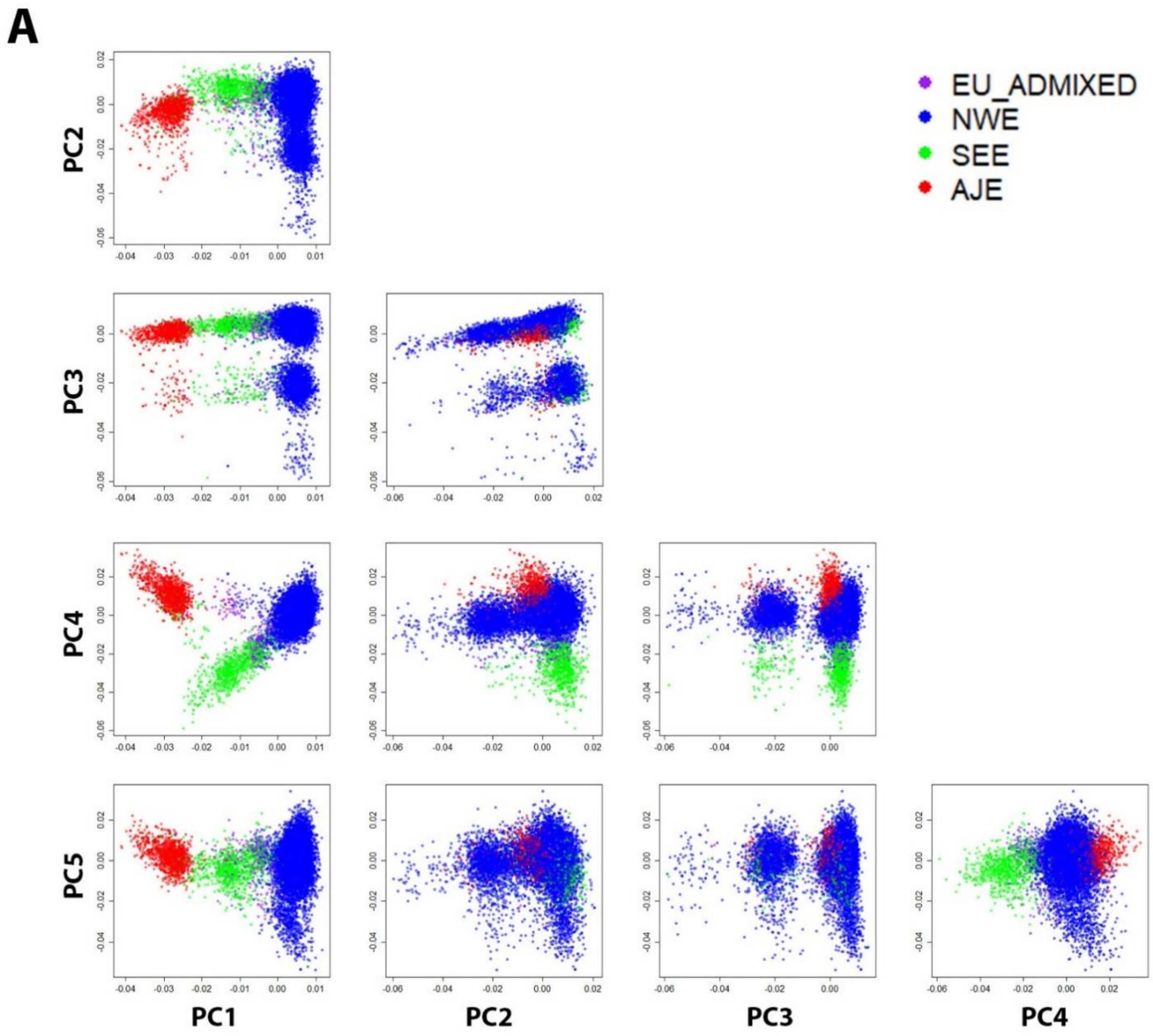

**Figure S2 (part 1). First five principal components of the genetic population structure in European subjects. (A) ADSP WES**. PCs are labelled by sub-European ancestries and confirm that sub-population stratification is well captured.

*Abbreviations: PC, principal component; EU, European; NWE, Northwestern European; SEE, Southeastern European; AJE, Ashkenazi Jewish.*

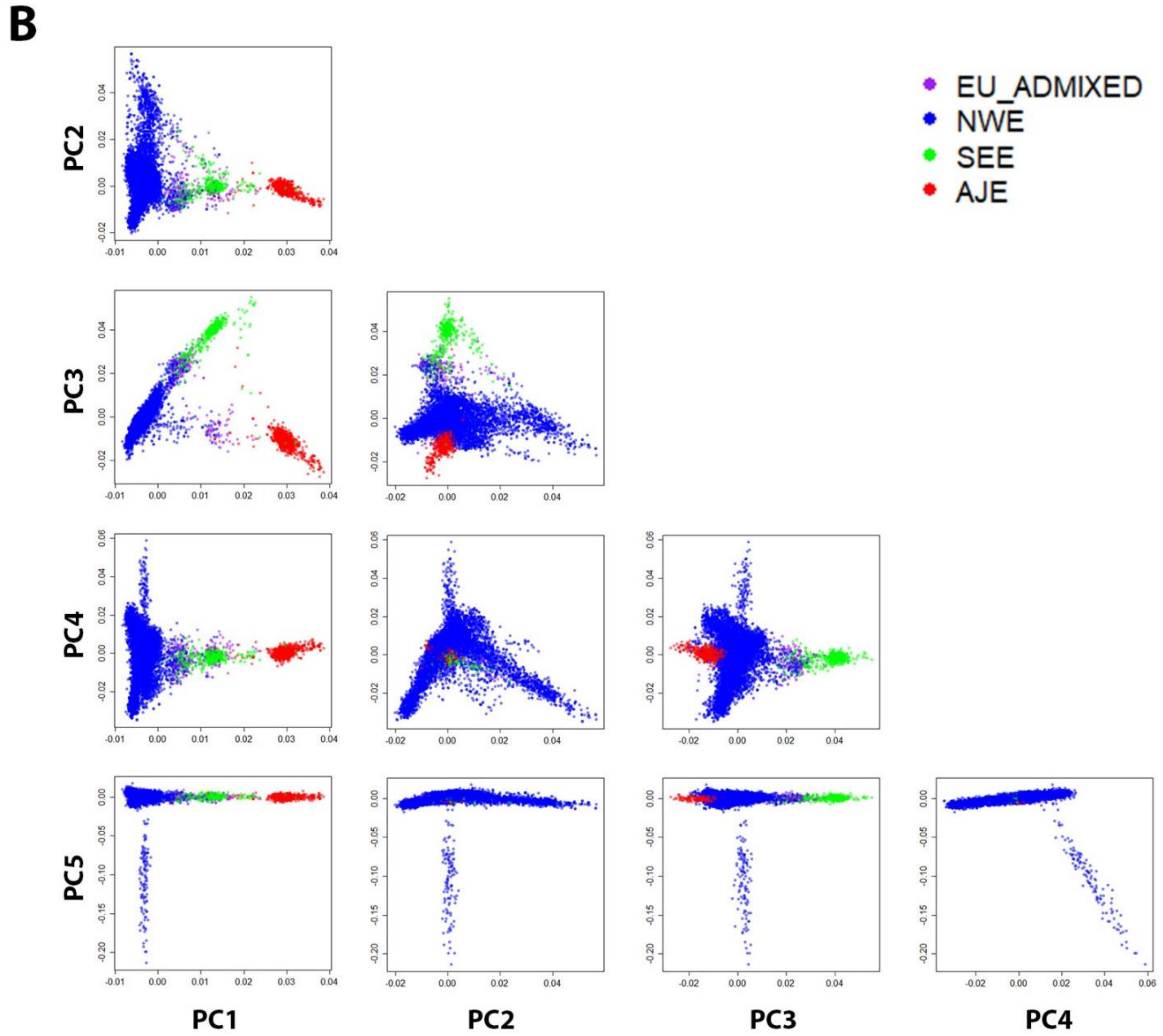

**Figure S2 (part 2). First five principal components of the genetic population structure in European subjects. (B) ADSP WGS**. PCs are labelled by sub-European ancestries and confirm that sub-population stratification is well captured. The NWE outliers in ADSP WGS (B, blue) correspond to samples from the Erasmus (Rotterdam) study, indicating they presented a distinct genetic background.

*Abbreviations: PC, principal component; EU, European; NWE, Northwestern European; SEE, Southeastern European; AJE, Ashkenazi Jewish.*

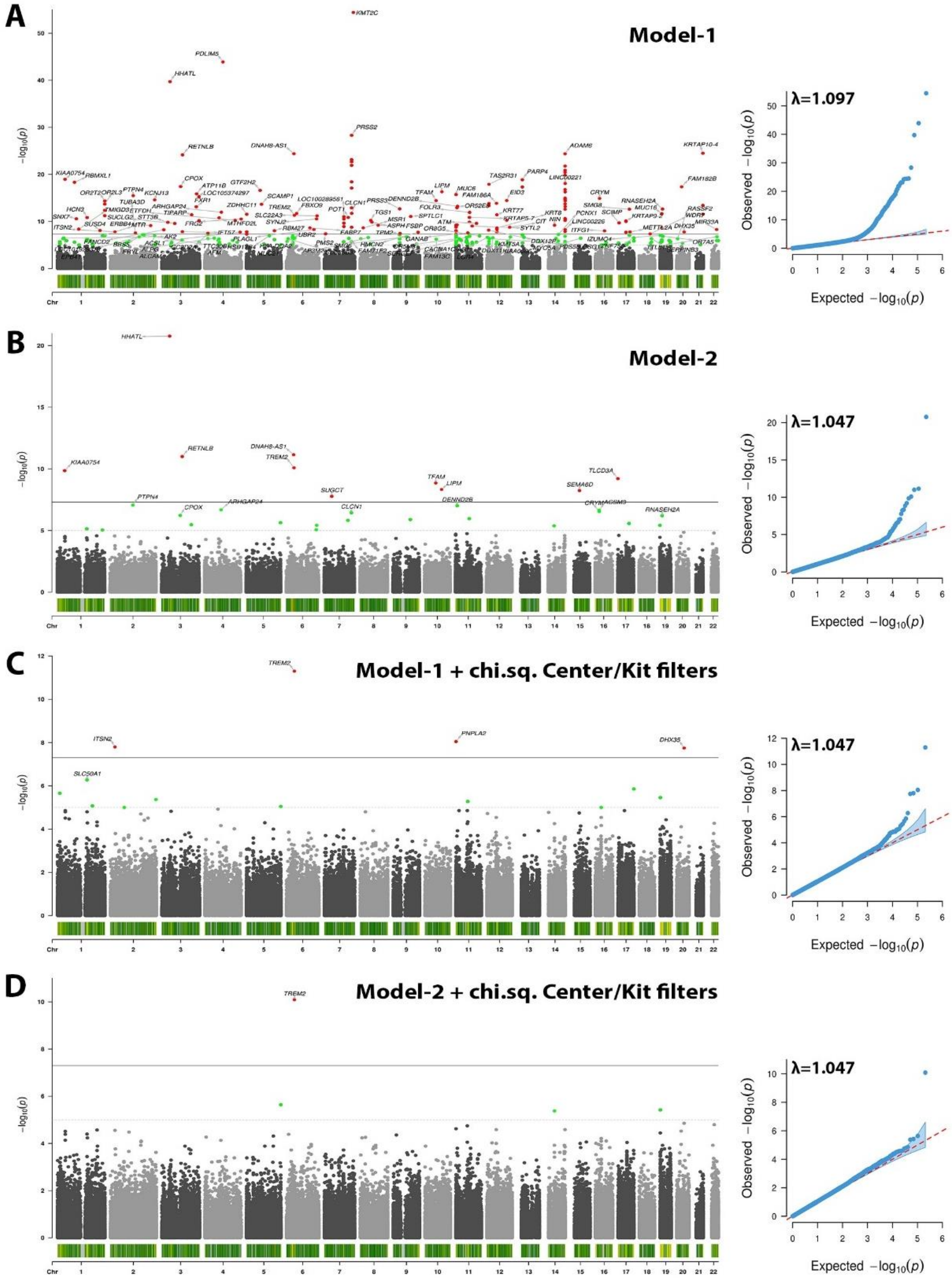

**Figure S3. The proposed center/kit-based variant filters, using chi square tests in R, remove spurious associations in ADSP WES.** Figure shows Manhattan (left) and quantile-quantile (right) plots. **A)** Model-1 indicates many spurious hits. **B)** Model-2 shows that adjustment for center/kit can reduce many, but not all, spurious hits. **C)** Filters remove most spurious hits. **D)** Further adjustment for center/kit removes few additional spurious hits.

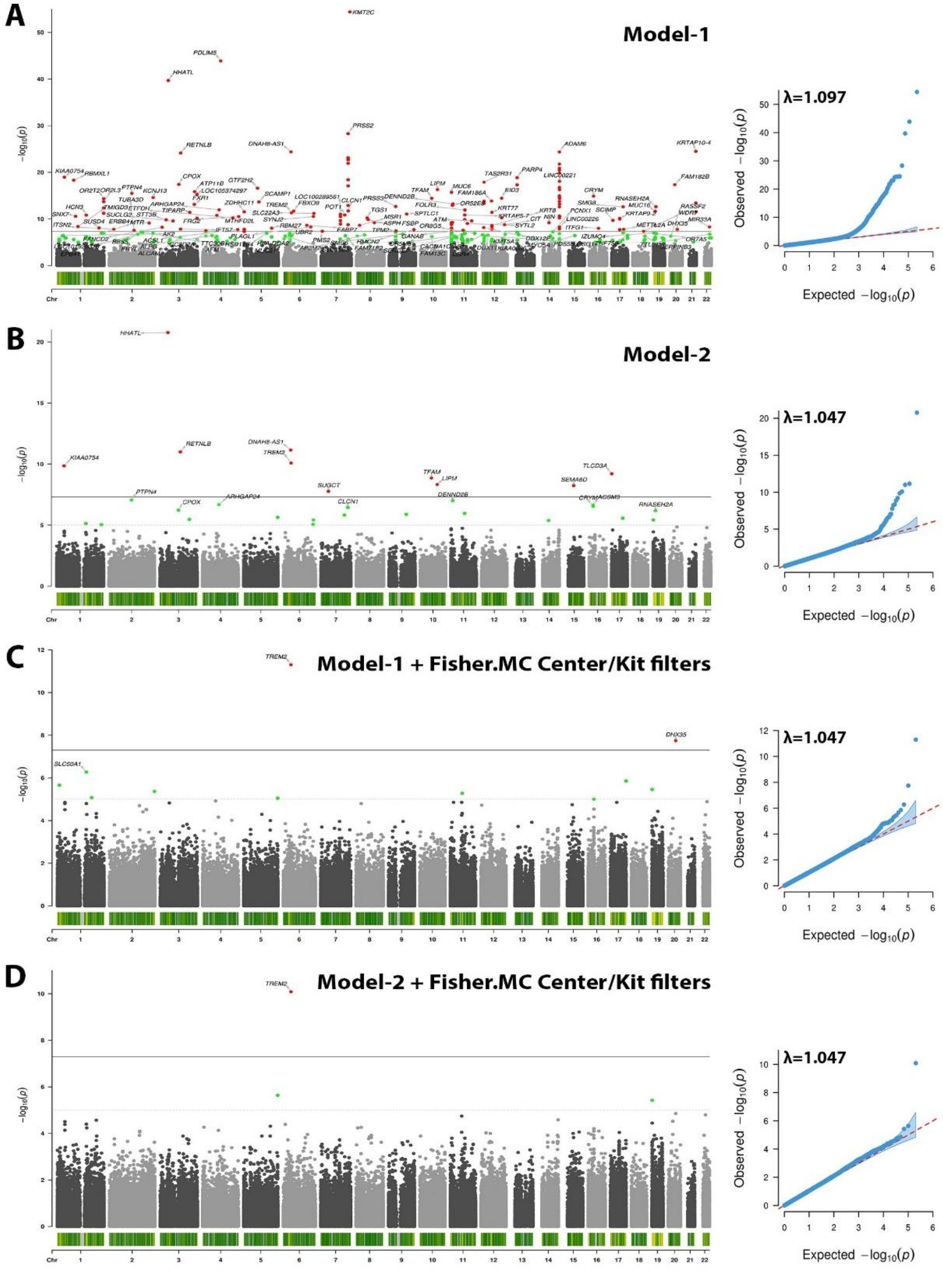

**Figure S4. The proposed center/kit-based variant filters, using Fisher exact Monte Carolo simulation tests in R, remove spurious associations in ADSP WES.** Figure shows Manhattan (left) and quantile-quantile (right) plots. **A)** Model-1 indicates many spurious hits. **B)** Model-2 shows that adjustment for center/kit can reduce many, but not all, spurious hits. **C)** Filters remove most spurious hits. **D)** Further adjustment for center/kit removes few additional spurious hits.

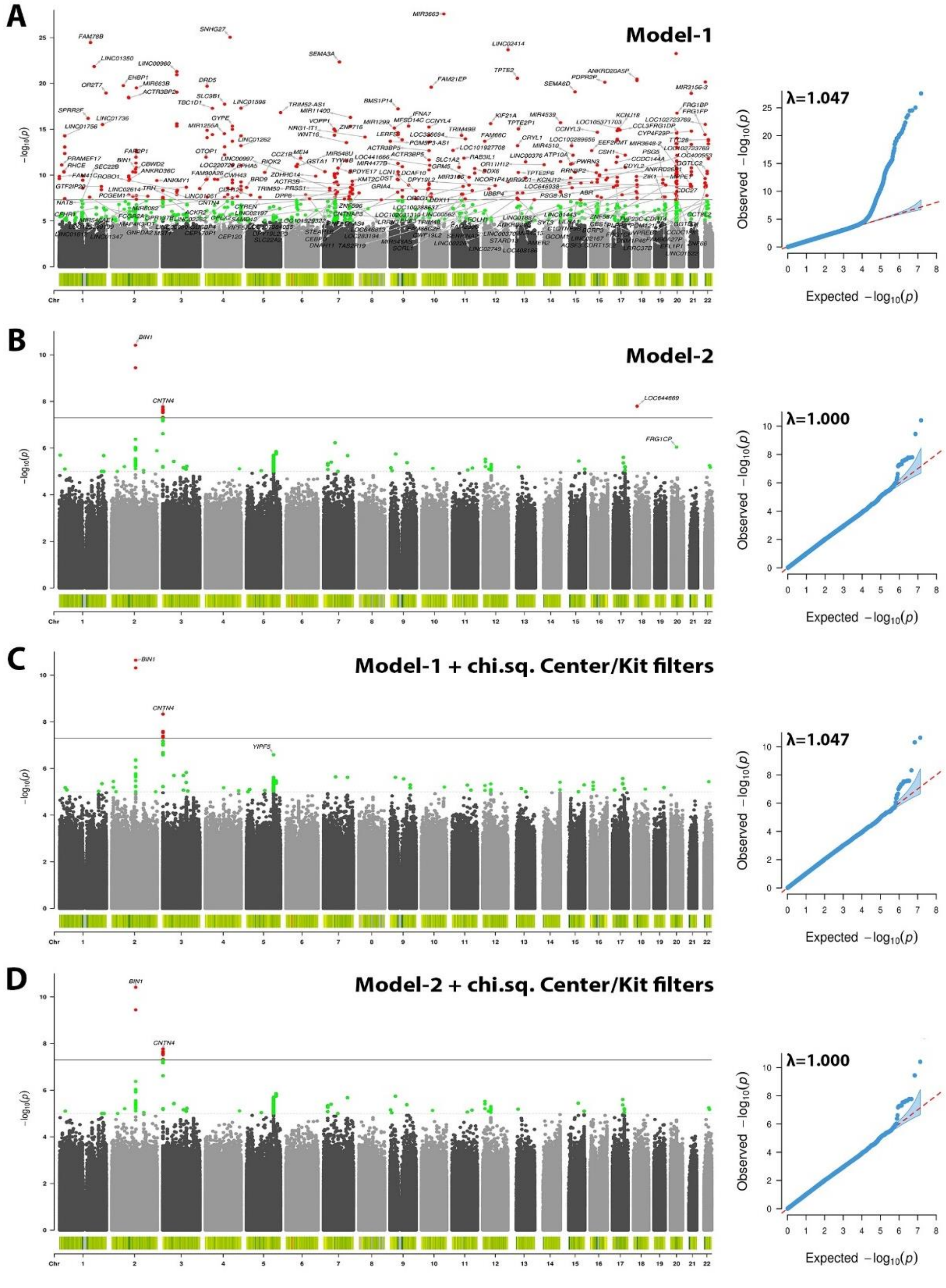

**Figure S5. The proposed center/kit-based variant filters, using chi square tests in R, remove spurious associations in ADSP WGS.** Figure shows Manhattan (left) and quantile-quantile (right) plots. **A)** Model-1 indicates many spurious hits. **B)** Model-2 shows that adjustment for center/kit can reduce many, but not all, spurious hits. **C)** Filters remove most spurious hits. **D)** Further adjustment for center/kit removes few additional spurious hits.

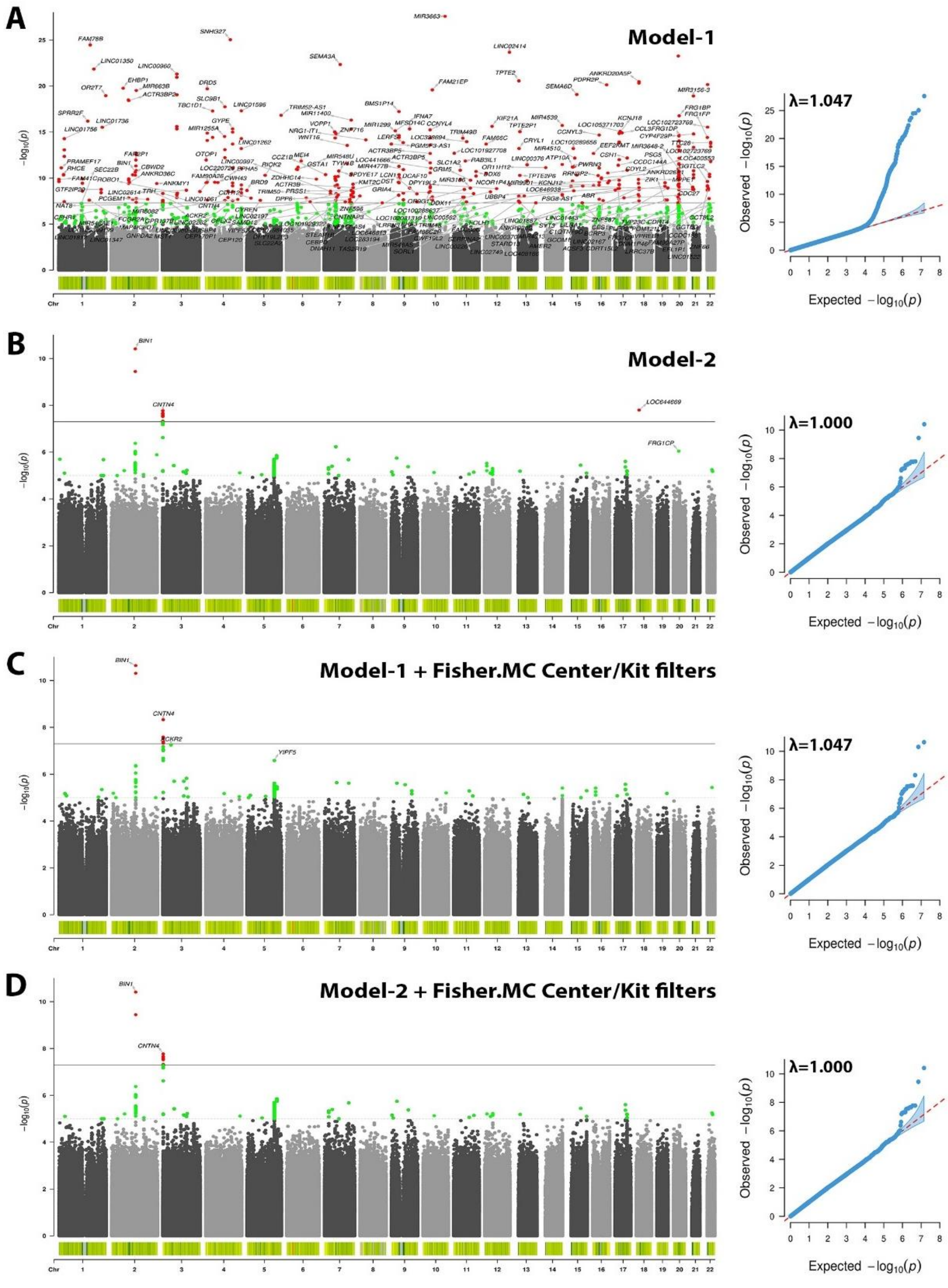

**Figure S6. The proposed center/kit-based variant filters, using Fisher exact Monte Carolo simulation tests in R, remove spurious associations in ADSP WGS.** Figure shows Manhattan (left) and quantile-quantile (right) plots. **A)** Model-1 indicates many spurious hits. **B)** Model-2 shows that adjustment for center/kit can reduce many, but not all, spurious hits. **C)** Filters remove most spurious hits. **D)** Further adjustment for center/kit removes few additional spurious hits.

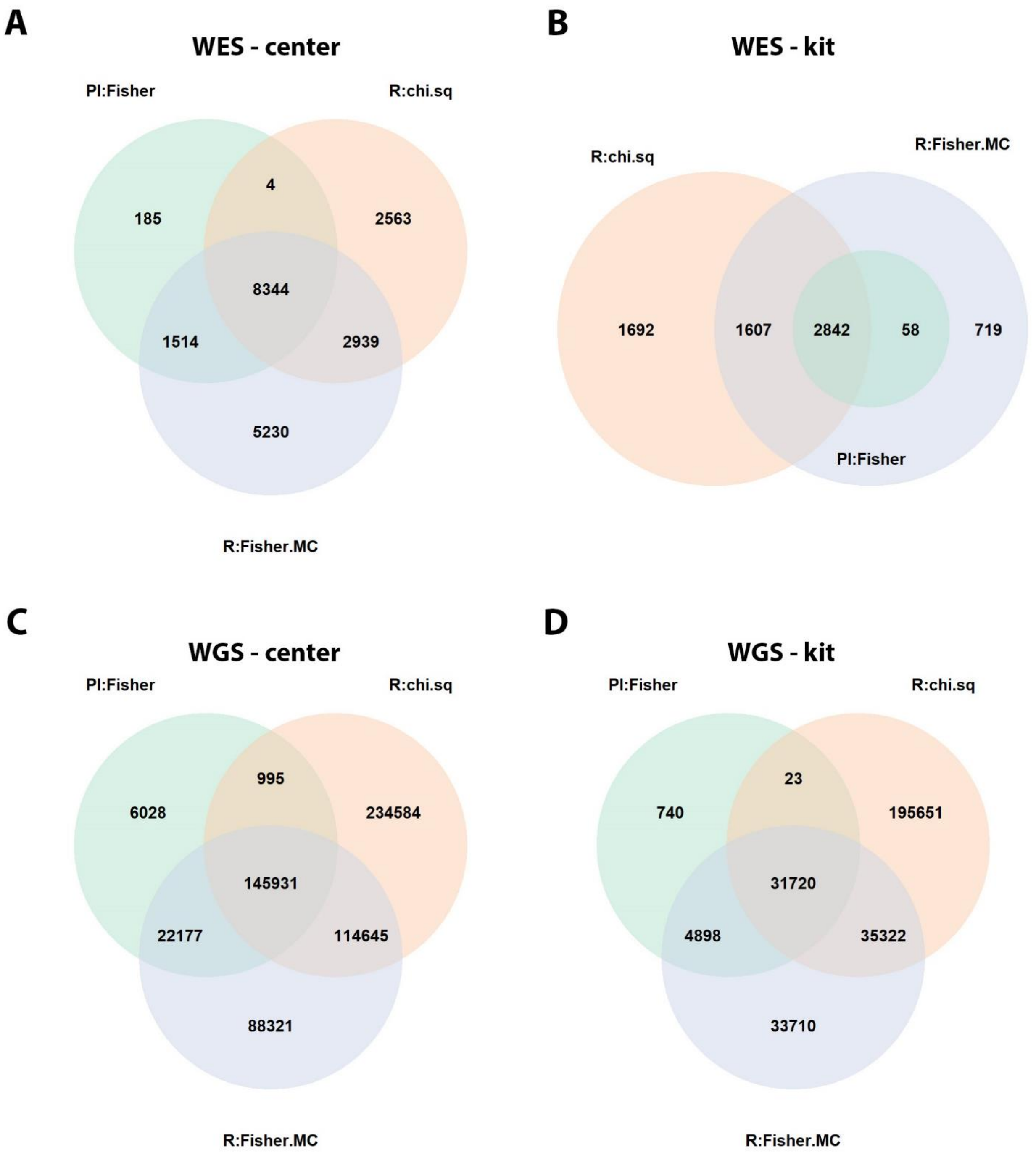

**Figure S7. Variant overlap between three types of considered sequencing center/kit-based filters. A)**. ADSP WES – sequencing center. **B)** ADSP WES – sequencing kit. **C)** ADSP WGS – sequencing center. **D)** ADSP WGS – sequencing kit.

*Abbrevations: Pl:Fisher, Plink-based Fisher test; R:chi.sq, R-based chi-square test; R:Fisher.MC, R-based fisher exact test with Monte Carlo simulation.*

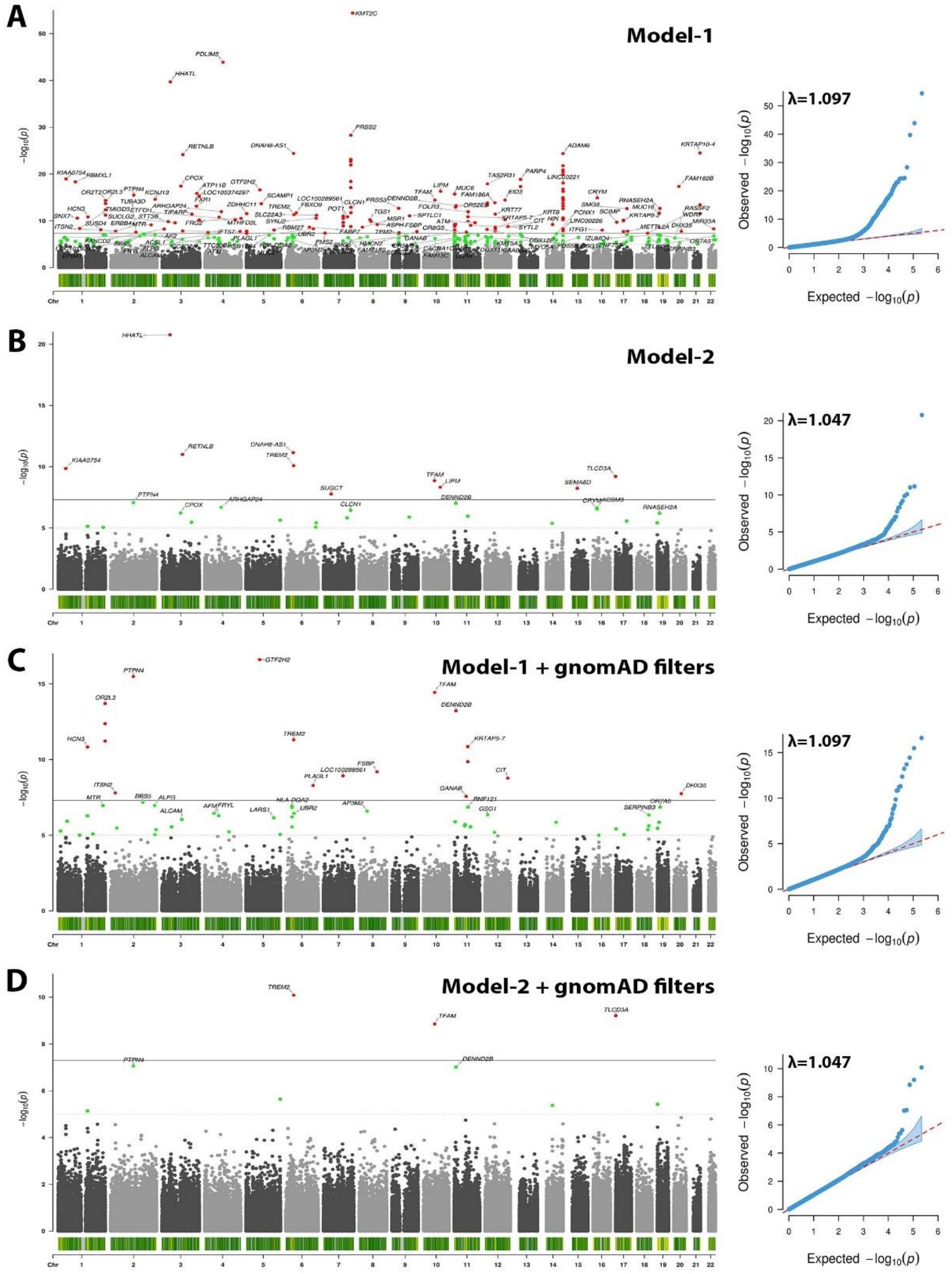

**Figure S8. The proposed gnomAD-based filters partially remove spurious associations in ADSP WES.** Figure shows Manhattan (left) and quantile-quantile (right) plots. **A)** Model-1 indicates many spurious hits. **B)** Model-2 shows that adjustment for center/kit can reduce many, but not all, spurious hits. **C)** Filters remove many spurious hits but several remain and inflation remains at the same level as in (A). **D)** Further adjustment for center/kit removes additional spurious hits but not all.

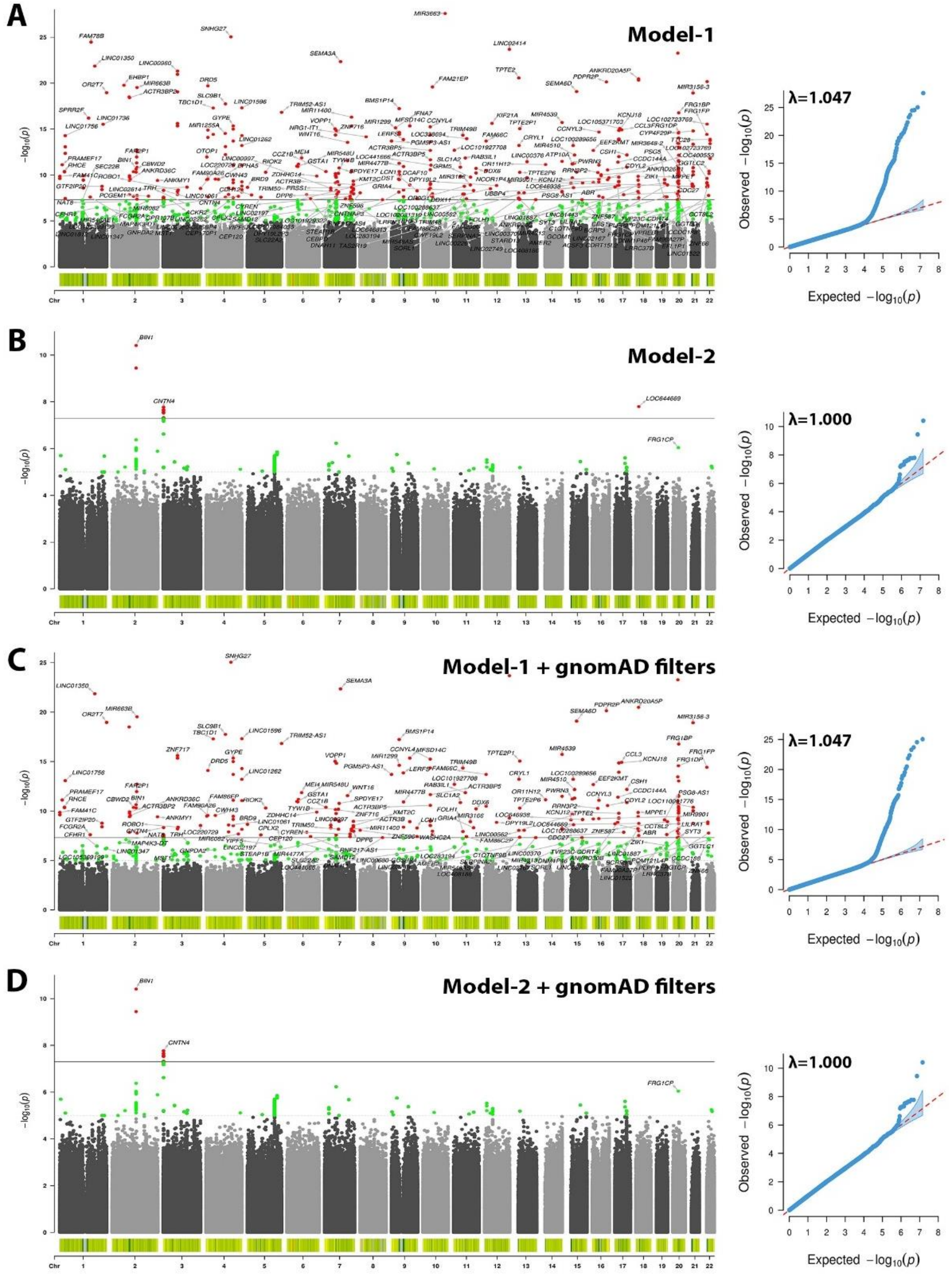

**Figure S9. The proposed gnomAD-based filters partially remove spurious associations in ADSP WGS.** Figure shows Manhattan (left) and quantile-quantile (right) plots. **A)** Model-1 indicates many spurious hits. **B)** Model-2 shows that adjustment for center/kit can reduce many, but not all, spurious hits. **C)** Filters remove many spurious hits but many remain. **D)** Further adjustment for center/kit removes most remaing spurious hits but not all as shown in Figure 3D.

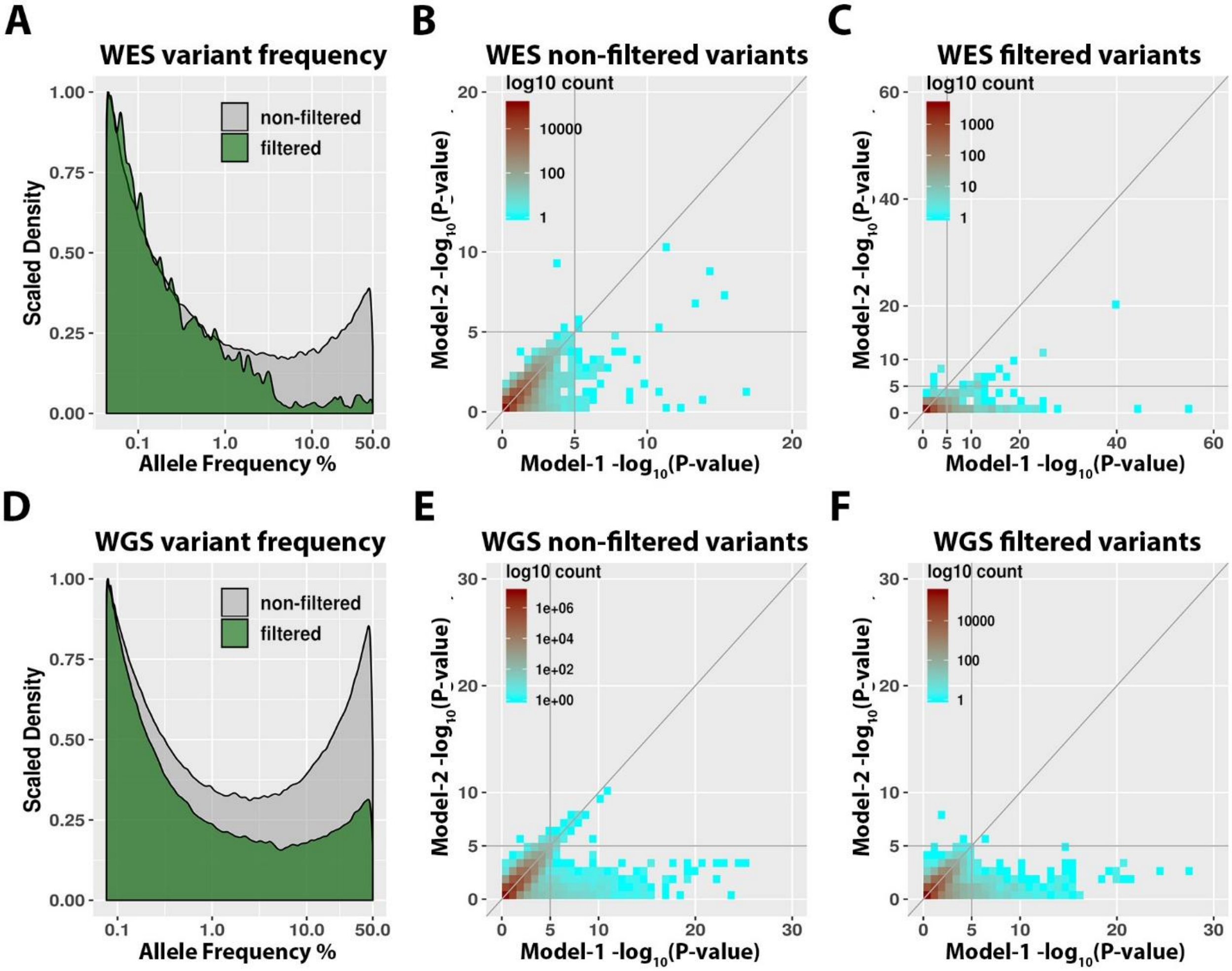

**Figure S10. Metrics of variants removed by the proposed gnomAD-based variant filters. A-C)** ADSP WES. **D-F)** ADSP WGS. **A & D)** Frequency density plots, comparing variants that were filtered/removed to those that were not filtered. Note that variants were not consistently filtered across the full frequency range, with decreased density at frequencies >1% in both ADSP WES and WGS. **B & E)** Variants that passed filters showed many inconsistent P-values across model-1 and model-2. **C & F)** Variants that were removed by filters showed even more inconsistent P-values across model-1 and model-2 as compared to (B & E).

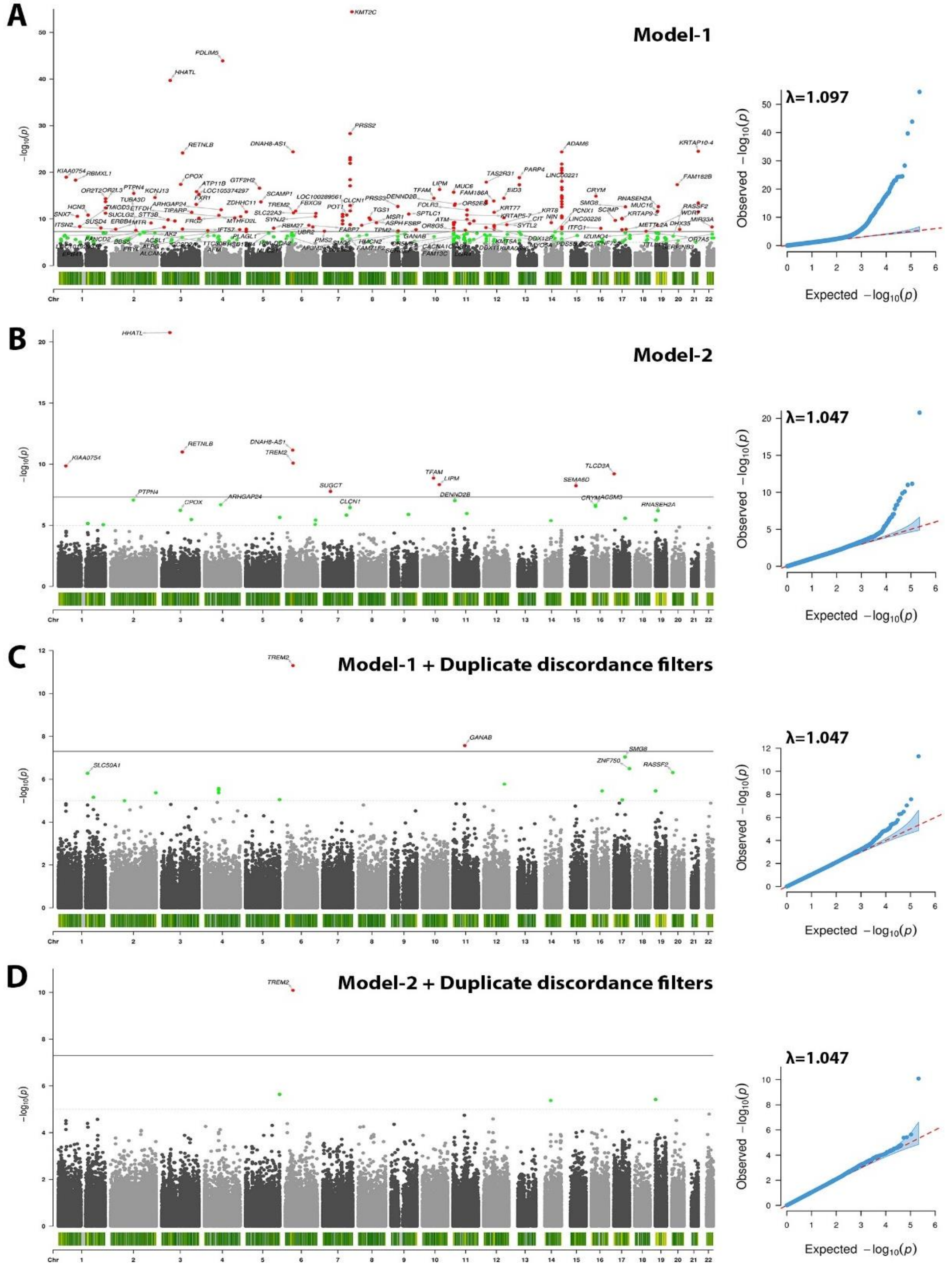

**Figure S11. The proposed duplicate discordant variant filters remove spurious associations in ADSP WES.** Figure shows Manhattan (left) and quantile-quantile (right) plots. **A)** Model-1 indicates many spurious hits. **B)** Model-2 shows that adjustment for center/kit can reduce many, but not all, spurious hits. **C)** Filter removes most spurious hits. **D)** Further adjustment for center/kit removes additional spurious hits.

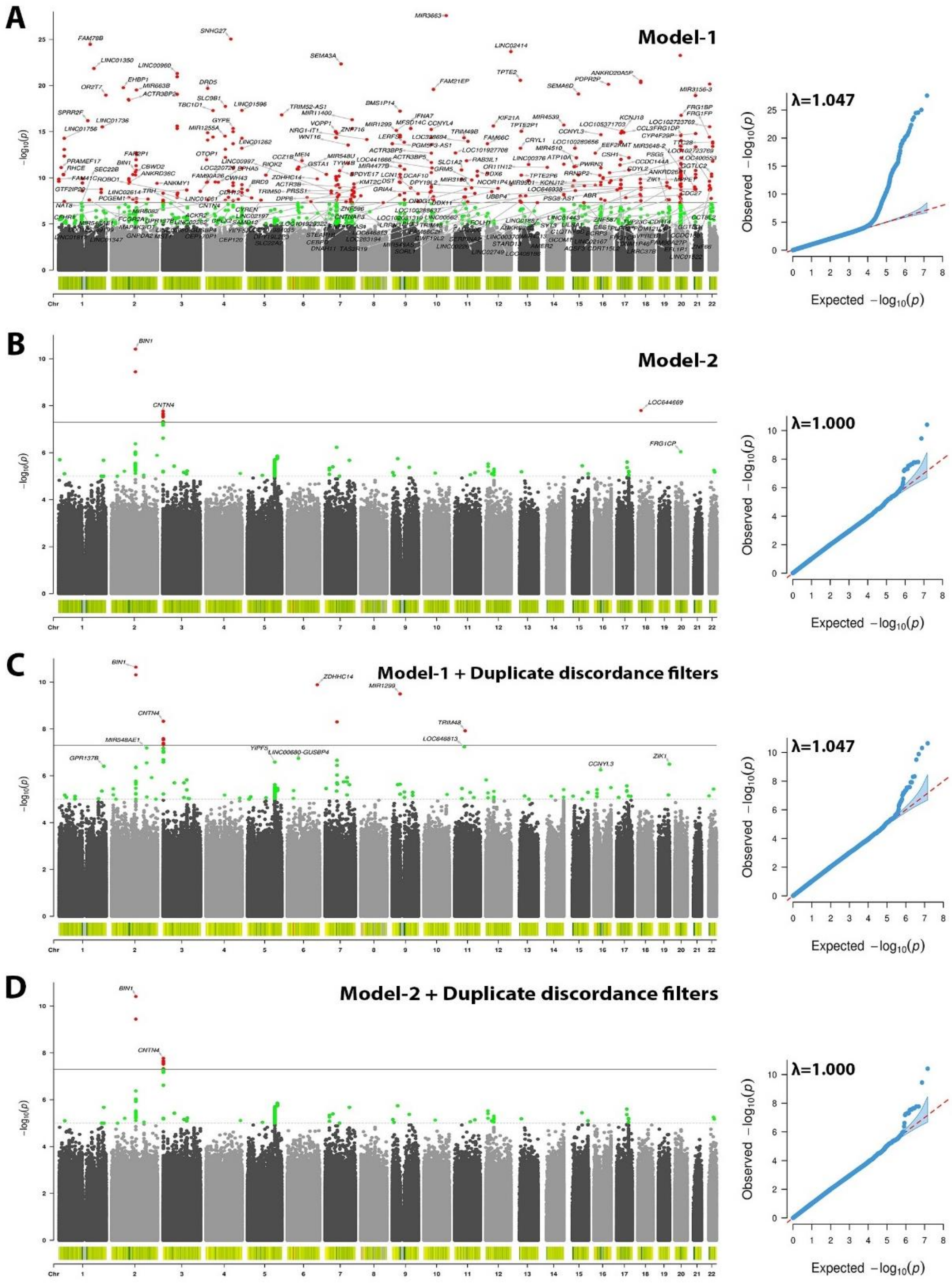

**Figure S12. The proposed duplicate discordant variant filters remove spurious associations in ADSP WGS.** Figure shows Manhattan (left) and quantile-quantile (right) plots. **A)** Model-1 indicates many spurious hits. **B)** Model-2 shows that adjustment for center/kit can reduce many, but not all, spurious hits. **C)** Filter removes many spurious hits, but not all. **D)** Further adjustment for center/kit removes additional spurious hits.

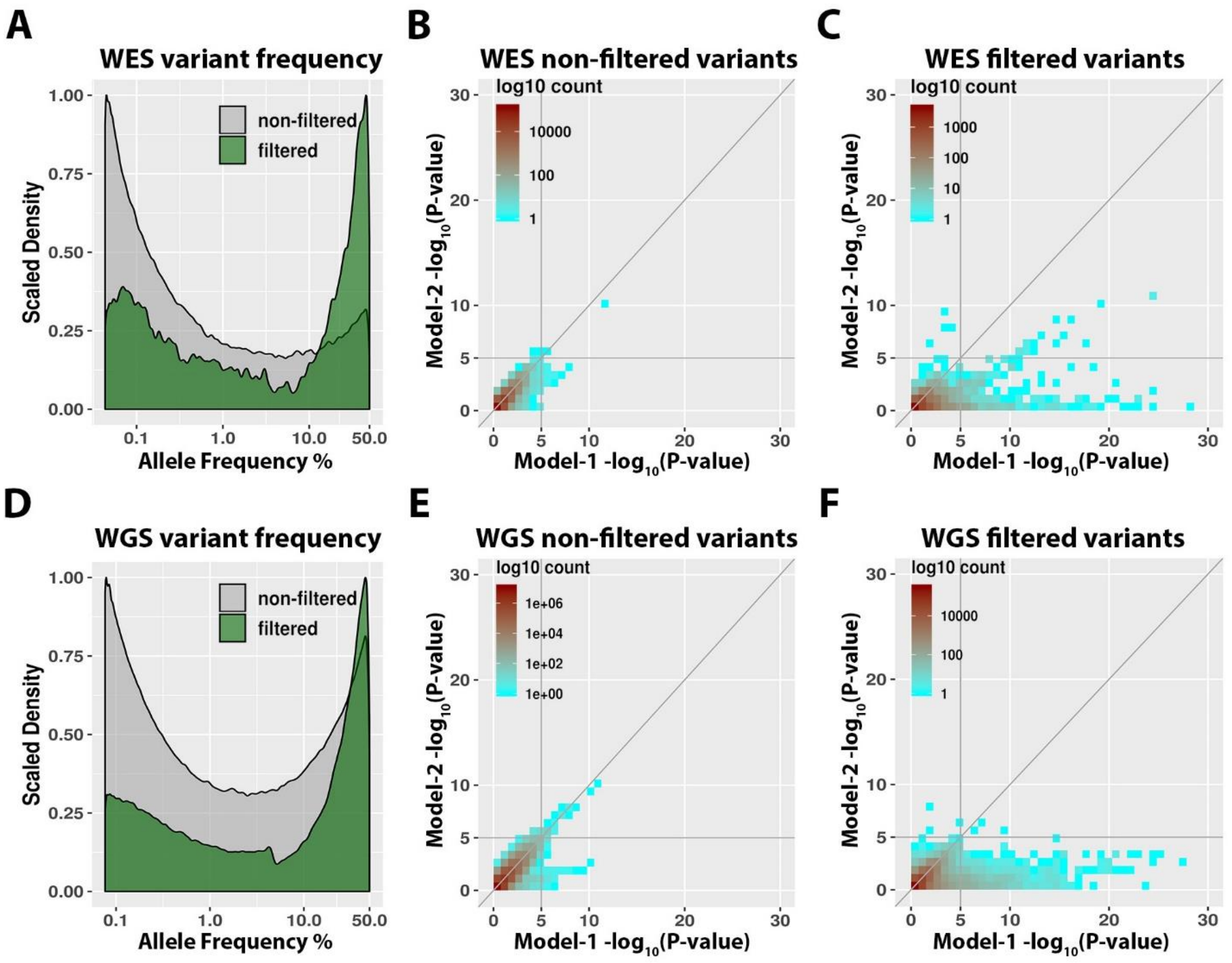

**Figure S13. Metrics of variants removed by the proposed duplicate discordance variant filters. A-C)** ADSP WES. **D-F)** ADSP WGS. **A & D)** Frequency density plots, comparing variants that were filtered/removed to those that were not filtered. Note that variants were not consistently filtered across the full frequency range, with decreased density at frequencies <10% in both ADSP WES and WGS. **B & E)** Variants that passed filters showed largely consistent P-values across model-1 and model-2 case-control association analyses, but there was still a set of variants remaining that reach suggestive significance in model-1 but lose suggestive significance upon center/kit adjustment in model-2 (lower right quadrant). **C & F)** Variants that were removed by filters showed many inconsistent P-values across model-1 and model-2, indicating that the duplicate discordance filters removed many center/kit-related variant artifacts that could not fully be accounted for by model-2.

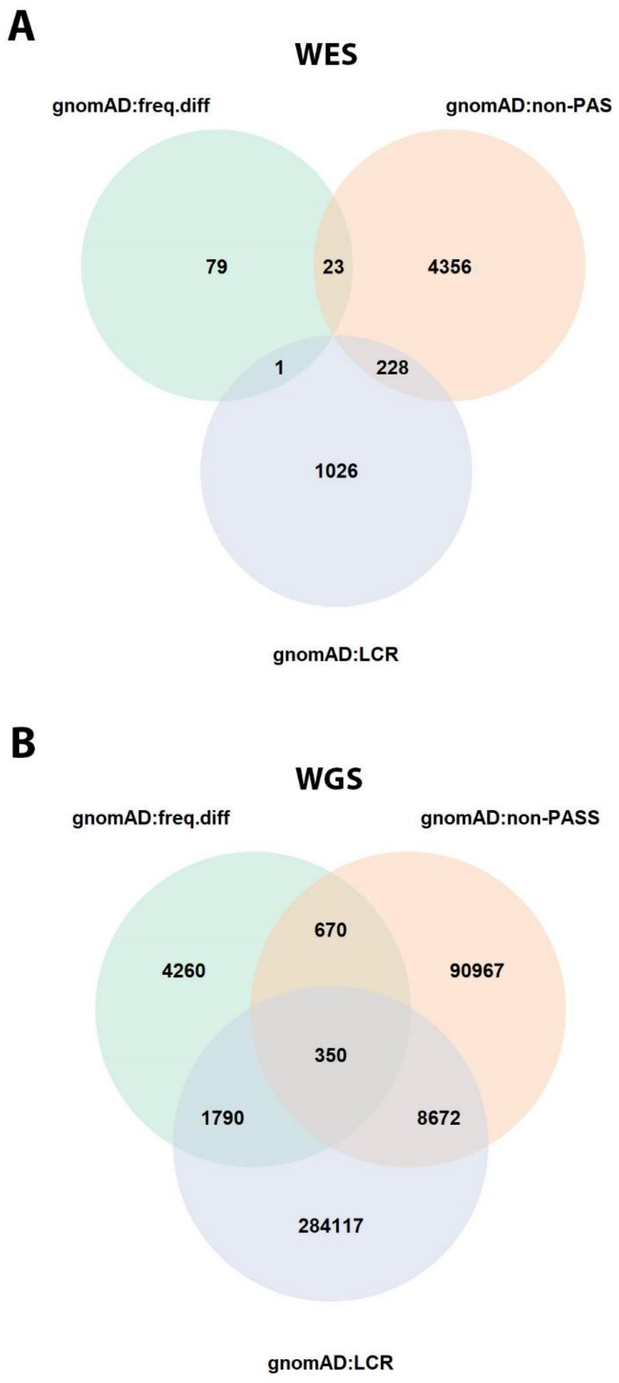

**Figure S14. Variant overlap between three types of considered gnomAD-based filters. A)** ADSP WES. **B)** ADSP WGS.

*Abbrevations: gnomAD:freq.diff, 10% frequency difference between gnomAD non-Finish Euopeans and ADSP Europeans; gnomAD:non-PASS, not having a PASS flag in gnomAD; gnomAD:LCR; gnomAD tagged low complexity region.*

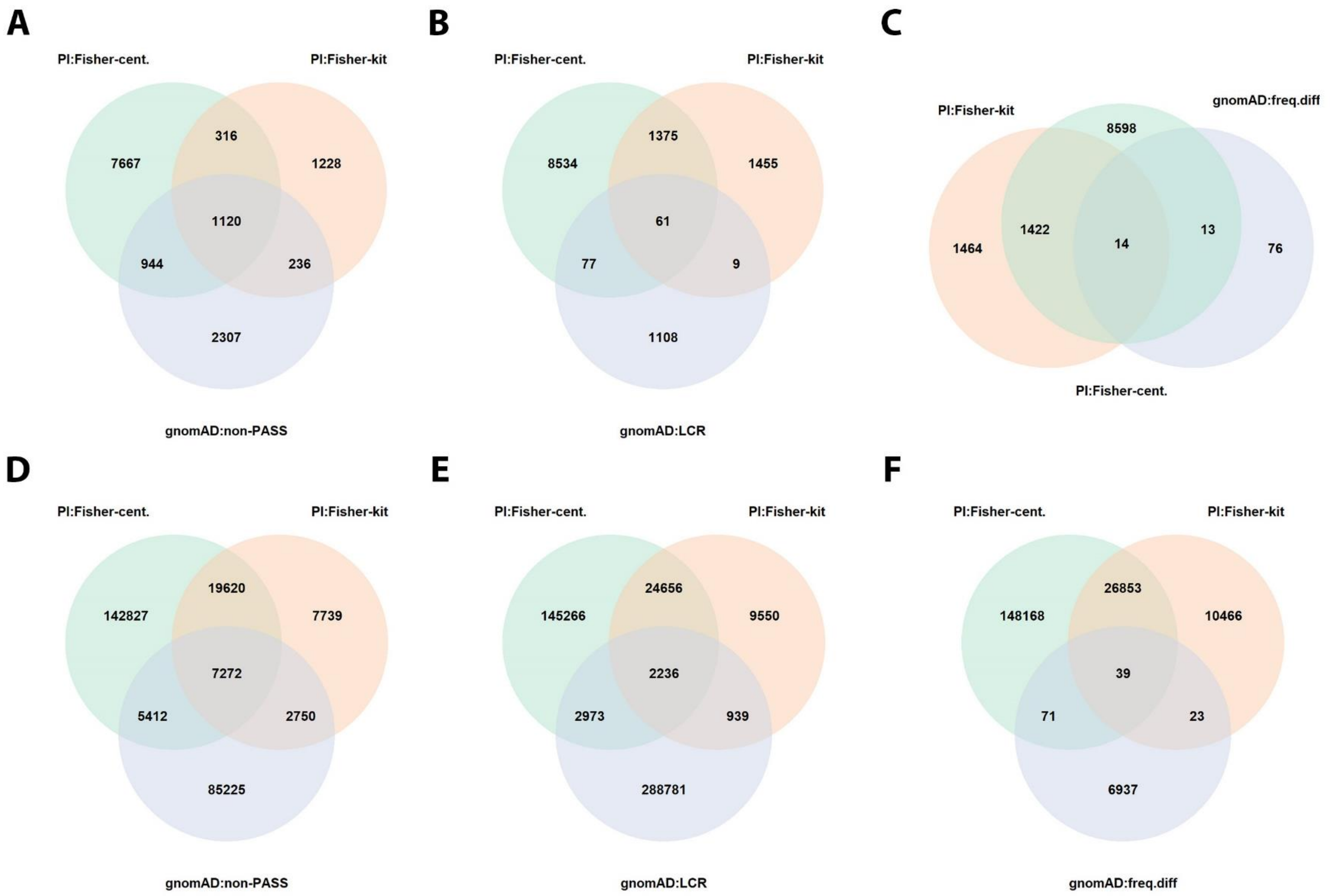

**Figure S15. Variant overlap between Plink Fisher-exact center/kit-based filters and gnomAD-based filters. A-C)** ADSP WES. **D-F)** ADSP WGS.

*Abbrevations: gnomAD:freq.diff, 10% frequency difference between gnomAD non-Finish Euopeans and ADSP Europeans; gnomAD:non-PASS, not having a PASS flag in gnomAD; gnomAD:LCR; gnomAD tagged low complexity region; Pl:Fisher, Plink-based Fisher test.*

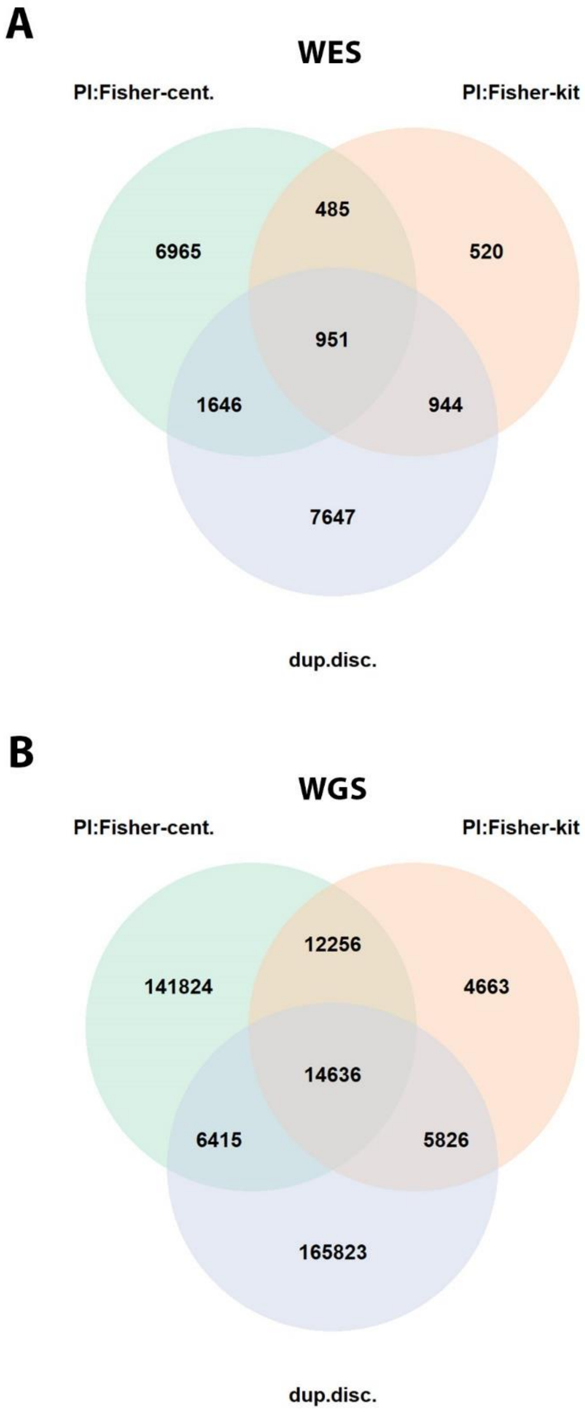

**Figure S16. Variant overlap between Plink Fisher-exact center/kit-based filters and duplicate discordant variant filters. A)** ADSP WES. **B)** ADSP WGS.

Abbrevations: dup.disc, duplicate discordant variants; Pl:Fisher, Plink-based Fisher test.

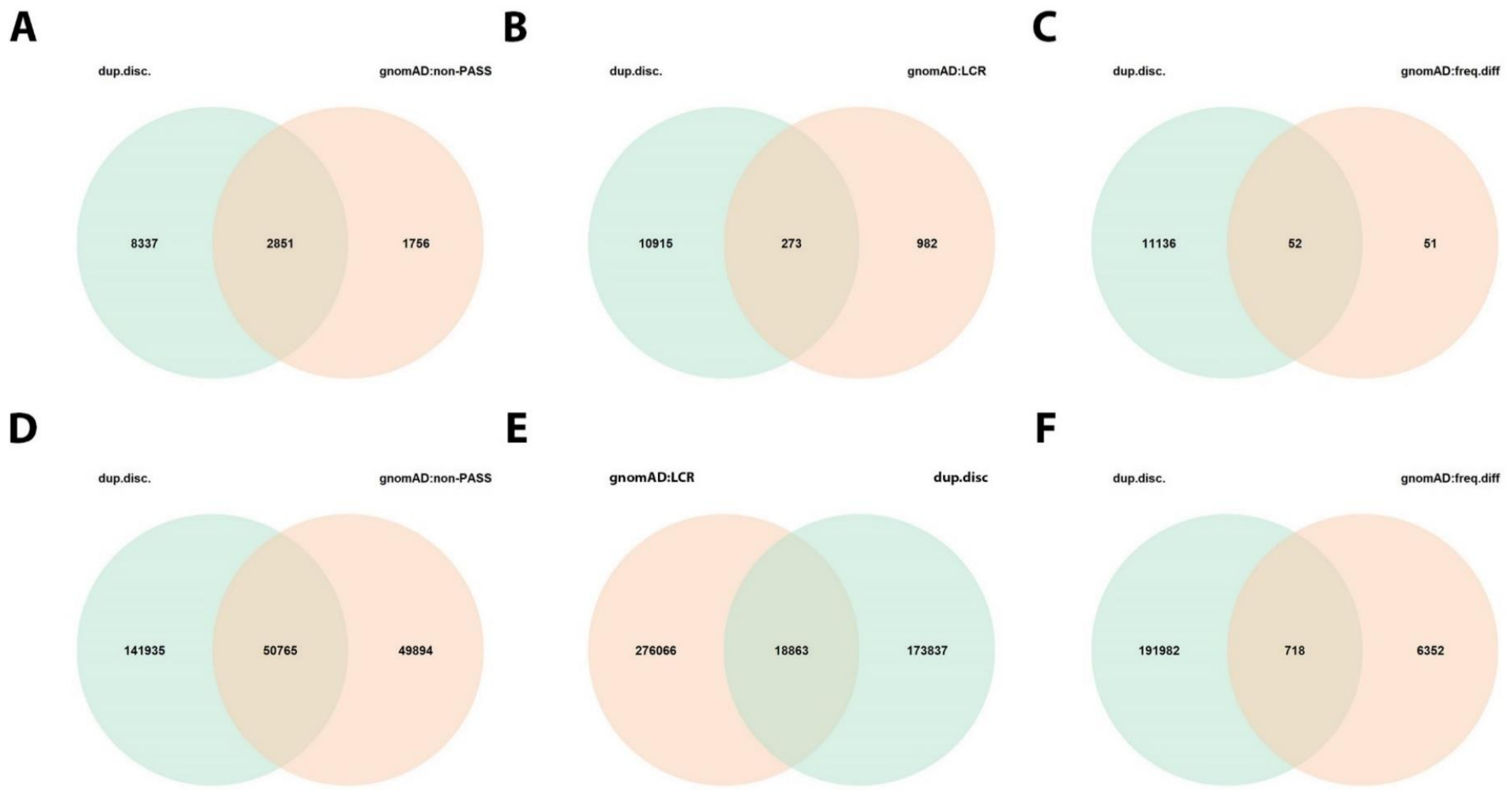

**Figure S17. Variant overlap between gnomAD-based filters and duplicate discordant variant filters. A-C)** ADSP WES**. D-F)** ADSP WGS**.**

Abbrevations: dup.disc, duplicate discordant variants; gnomAD:freq.diff, 10% frequency difference between gnomAD non-Finish Euopeans and ADSP Europeans; gnomAD:non-PASS, not having a PASS flag in gnomAD; gnomAD:LCR; gnomAD tagged low complexity region.

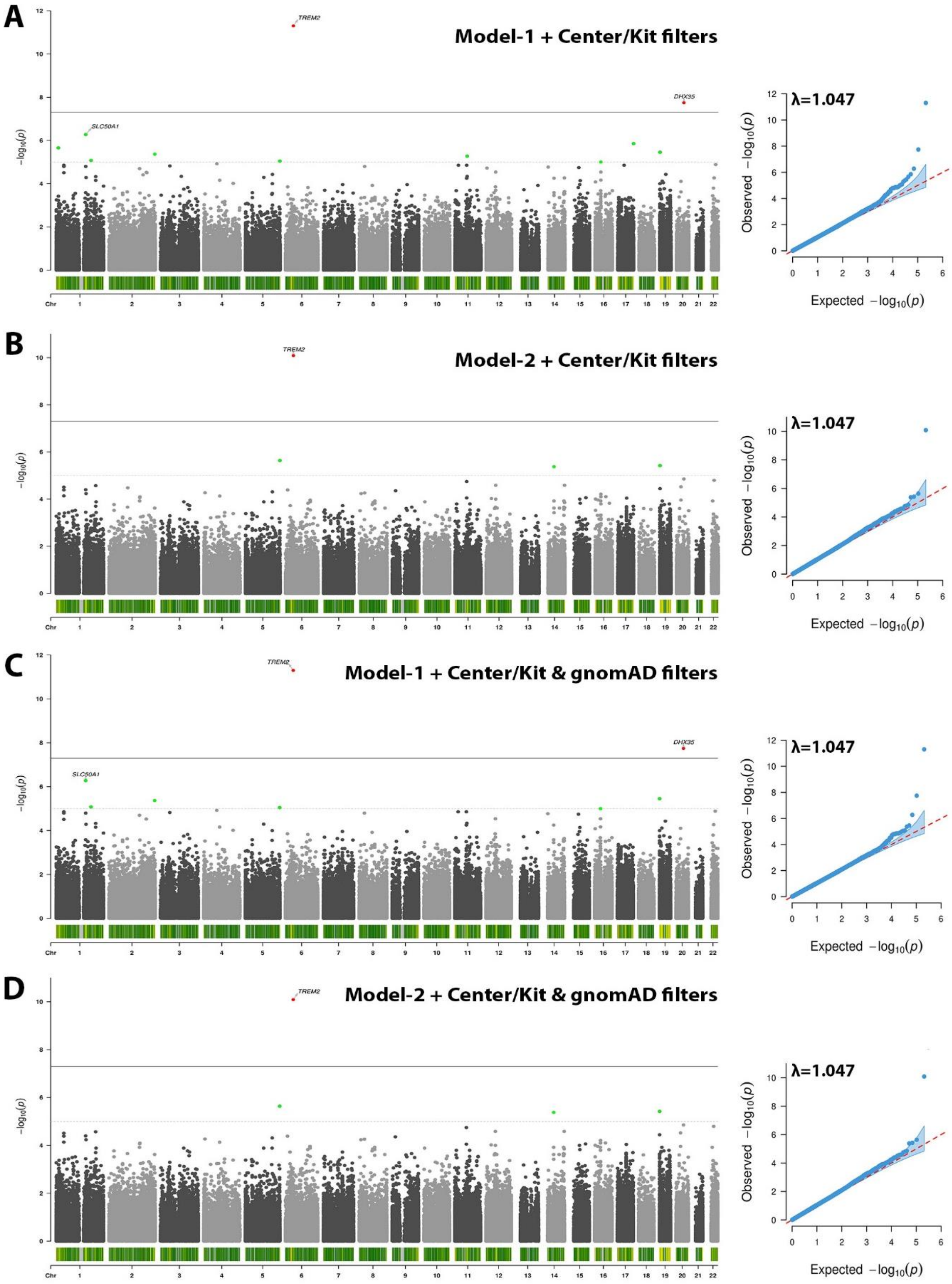

**Figure S18. GnomAD-based filters remove few additional spurious associations after applying center/kit-based variant filters in ADSP WES.**

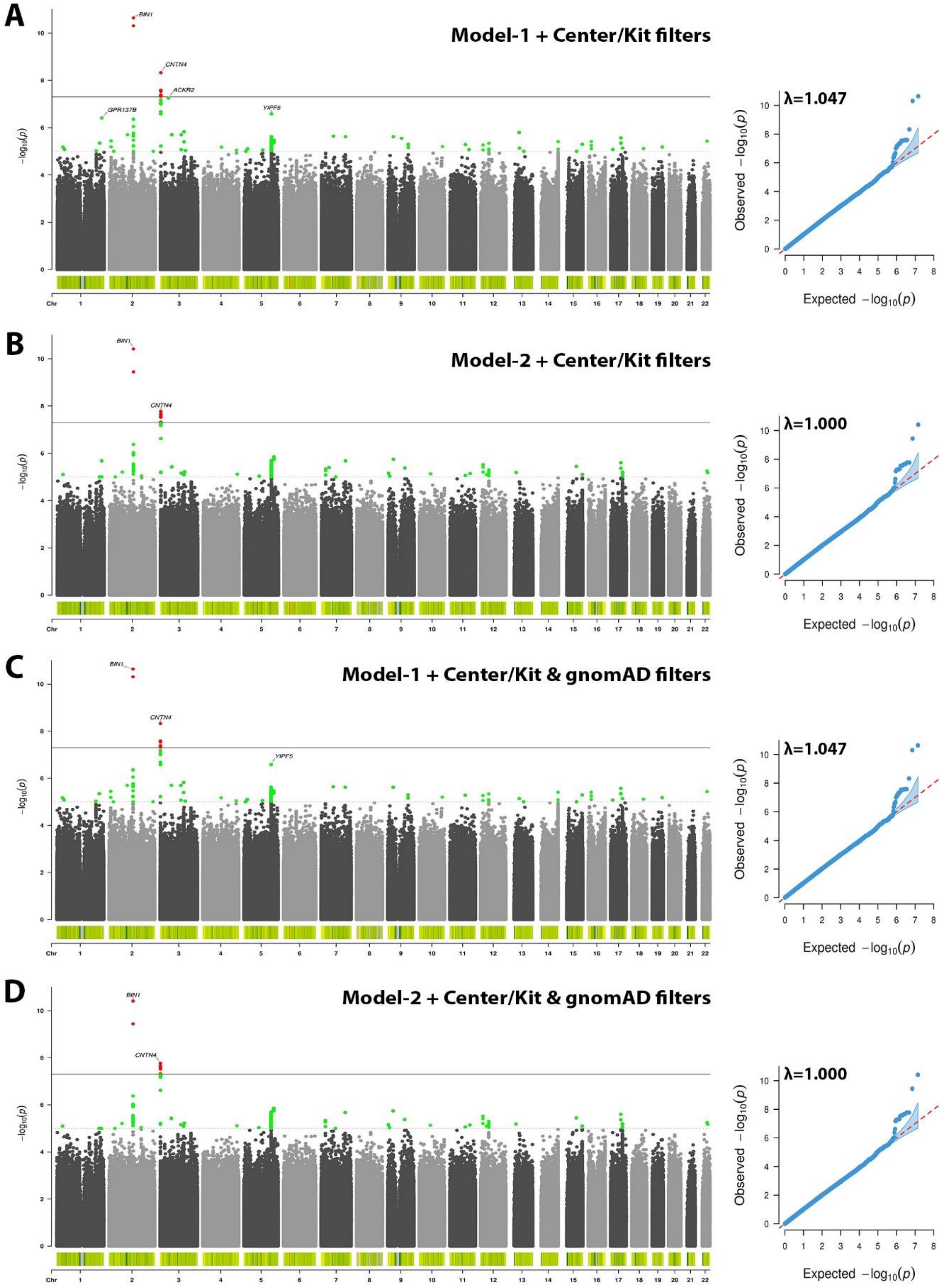

**Figure S19. GnomAD-based filters remove few additional spurious associations after applying center/kit-based variant filters in ADSP WGS.**

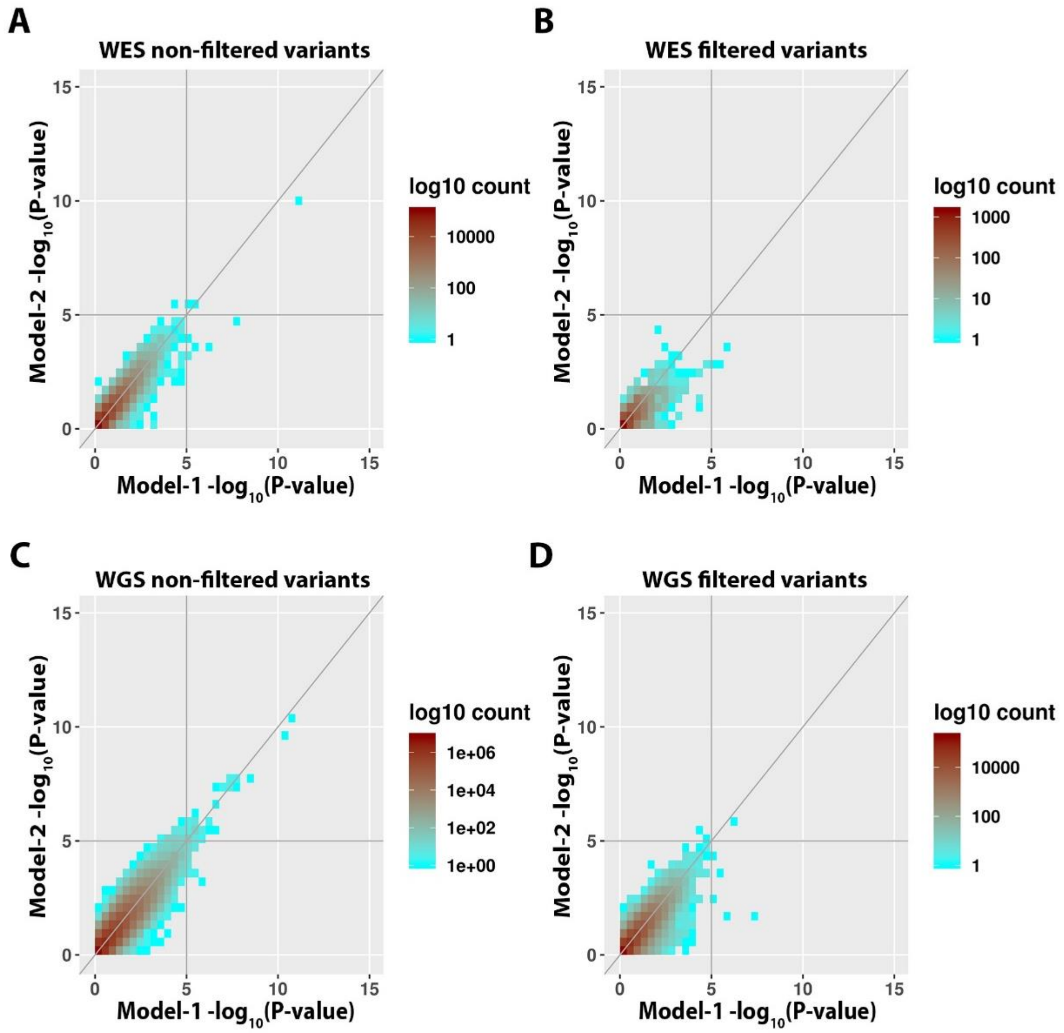

**Figure S20. Metrics of variants removed by the gnomAD-based variant filters after first applying center/kit-based variant filters. A-B)** ADSP WES. **C-D)** ADSP WGS. The density distributions appear largely consistent between non-filtered (A & C) and filtered (B & D) variants. There were few additional variants that reach suggestive significance in model-1 but lose suggestive significance upon center/kit adjustment in model-2 that were filtered (lower right quadrant in (B & D)), but some of this type variants still remained (lower right quadrant in (A & C)).

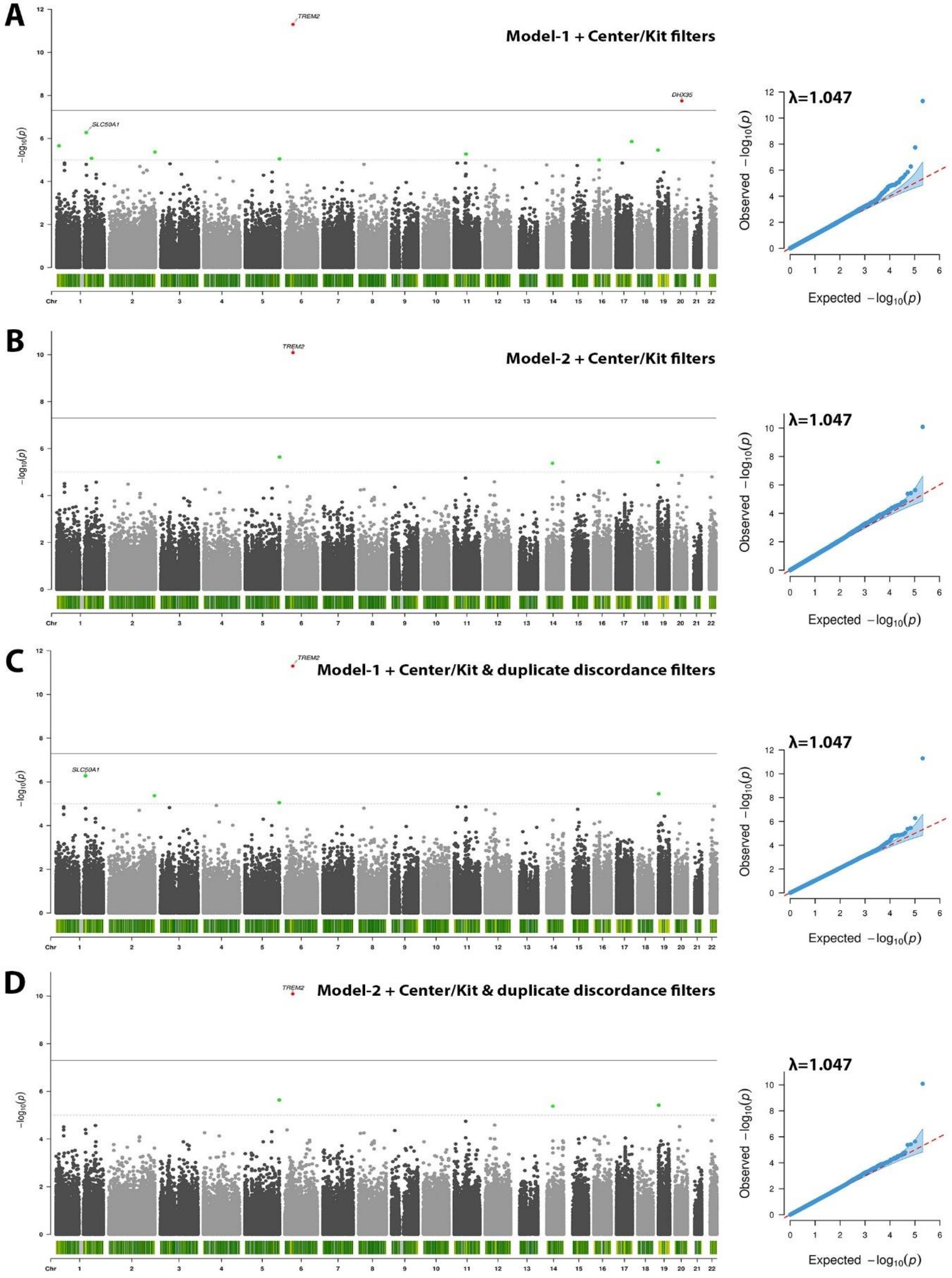

**Figure S21. Duplicate discordant variant filters remove few additional spurious associations after applying center/kit-based variant filters in ADSP WES.**

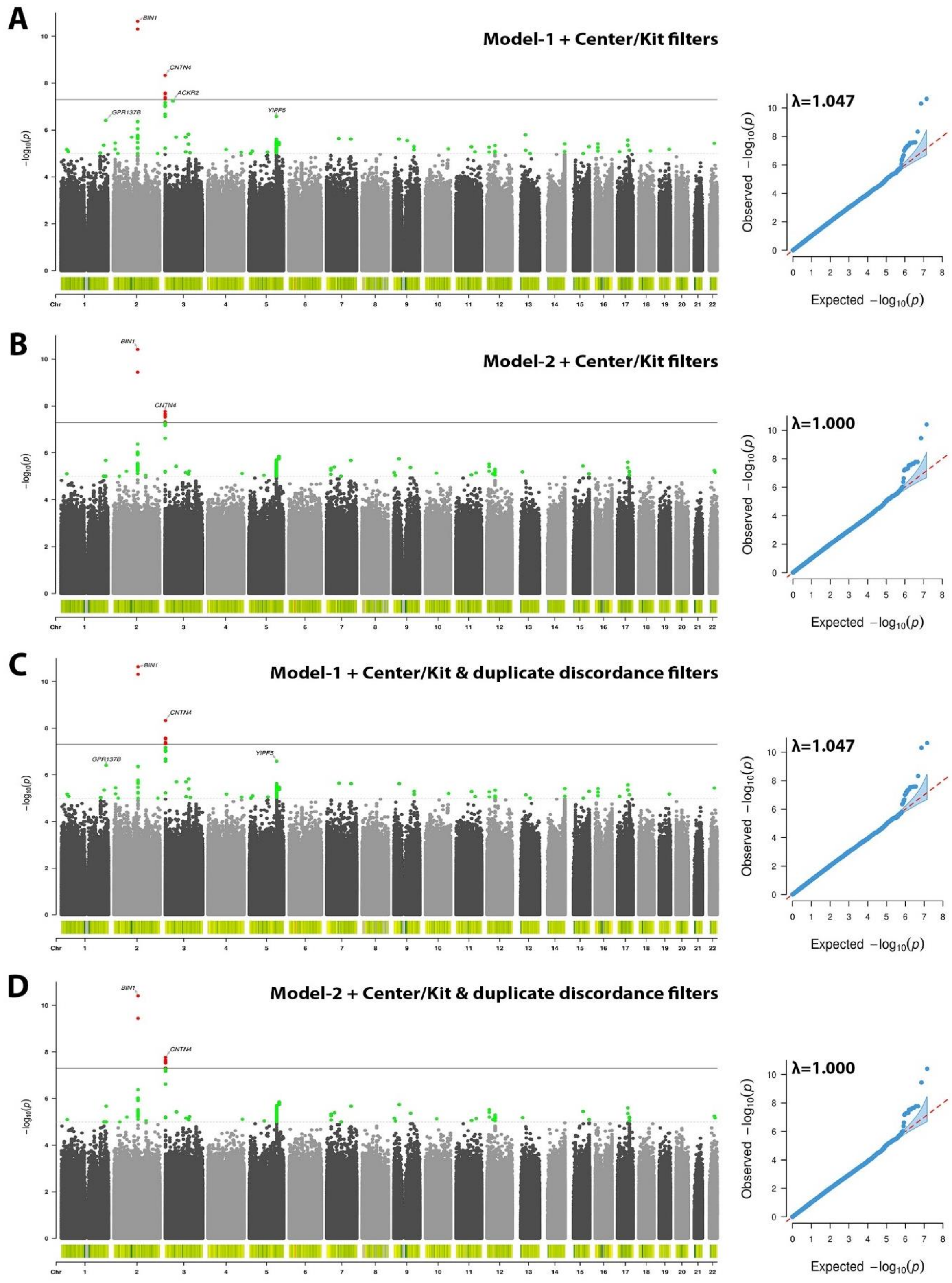

**Figure S22. Duplicate discordant variant filters remove few additional spurious associations after applying center/kit-based variant filters in ADSP WGS.**

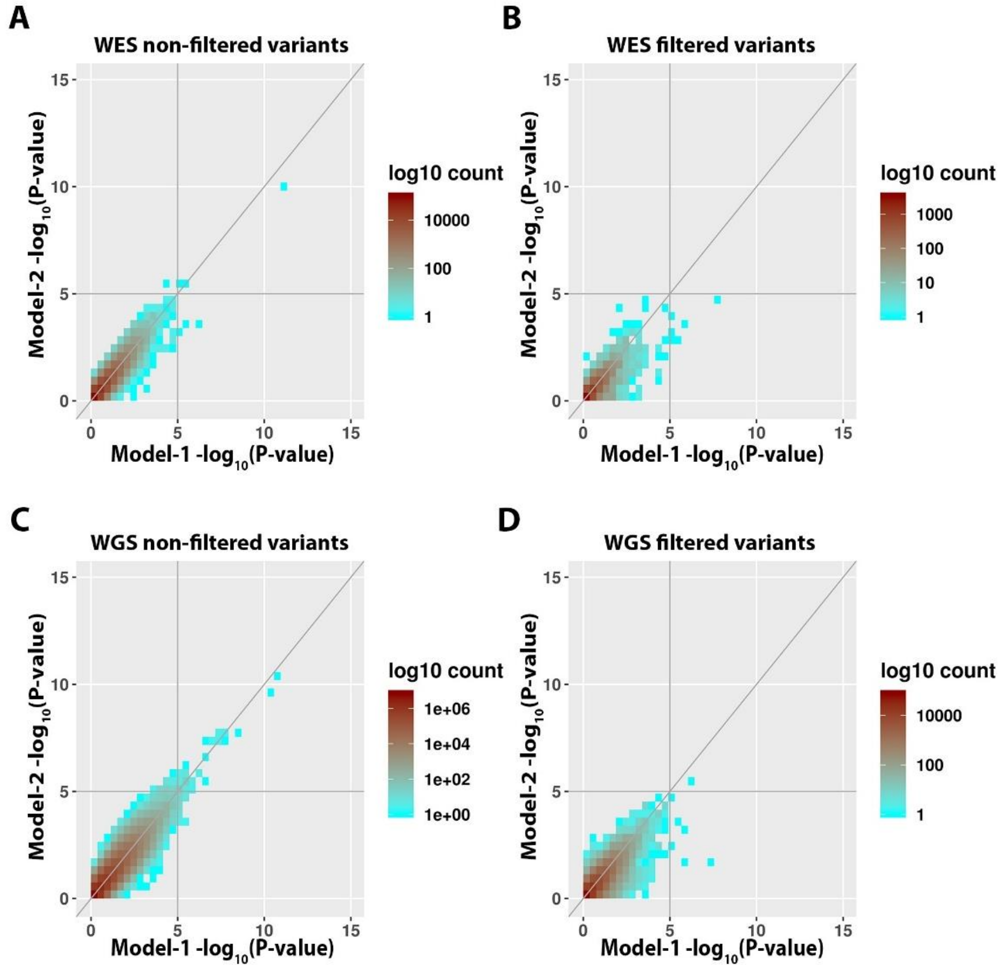

**Figure S23. Metrics of variants removed by the duplicate discordant variant filters after first applying center/kit-based variant filters. A-B)** ADSP WES. **C-D)** ADSP WGS. The density distributions appear largely consistent between non-filtered (A & C) and filtered (B & D) variants. There were few additional variants that reach suggestive significance in model-1 but lose suggestive significance upon center/kit adjustment in model-2 that were filtered (lower right quadrant in (B & D)); very few of this type variants still remained (lower right quadrant in (A & C)).

**Table S1. Overview of genotyping platforms across all available AD-related genetic data.**

| Cohort/Project | Genotyping Platform | Cohort-Platform ID | Sample count | Data Repository |
|---|---|---|---|---|
| A4 | Illumina Global Screening Array (GSA) | A4 | 3465 | LONI A4 |
| ACT | Illumina Human 660W-Quad | ACT | 2790 | NIAGADS (NG00034) / dbGaP (phs000234) |
| ADC1 | Illumina Human 660W-Quad | ADC1 | 2731 | NIAGADS (NG00022) / NACC |
| ADC2 | Illumina Human 660W-Quad | ADC2 | 928 | NIAGADS (NG00023) / NACC |
| ADC3 | Illumina Human OmniExpress | ADC3 | 1526 | NIAGADS (NG00024) / NACC |
| ADC4 | Illumina Human OmniExpress | ADC4 | 1054 | NIAGADS (NG00068) / NACC |
| ADC5 | Illumina Human OmniExpress | ADC5 | 1224 | NIAGADS (NG00069) / NACC |
| ADC6 | Illumina Human OmniExpress | ADC6 | 1333 | NIAGADS (NG00070) / NACC |
| ADC7 | Illumina Infinium Human OmniExpressExome | ADC7 | 1462 | NIAGADS (NG00071) / NACC |
| ADDNEUROMED | Illumina Human 610-Quad | ADM_Q | 315 | Synapse AddNeuroMed (syn4907804) |
| | Illumina Human OmniExpress | ADM_O | 329 | Synapse AddNeuroMed (syn4907804) |
| ADNI | Illumina Human 610-Quad | ADNI_1 | 757 | LONI ADNI |
| | Illumina Human OmniExpress | ADNI_2 | 361 | LONI ADNI |
| | Illumina Global Screening Array (GSA) | ADNI_3 | 327 | LONI ADNI |
| | Illumina Omni 2.5 | ADNI_O25 | 812 | LONI ADNI |
| | Whole Genome Sequencing - Illumina | ADNI_WGS | 812 | LONI ADNI |
| ADNI-DOD | Illumina Human OmniExpress | ADNI_DOD | 204 | LONI ADNIDOD |
| ADGC Exome-Arrays | Illumina HumanExome BeadChip v1.0 - CHOP | CHOP | 5180 | NIAGADS (NG00081) / NACC |
| | Illumina HumanExome BeadChip v1.0 - Miami | MIA | 1923 | NIAGADS (NG00080) / NACC |
| | Illumina HumanExome BeadChip v1.0 - Northshore | NS | 5998 | NIAGADS (NG00079) / NACC |
| | Illumina HumanExome BeadChip v1.0 - WashU | WU | 868 | NIAGADS (NG00085) / NACC |
| **ADSP WES** | **Whole Exome Sequencing** | **ADSP_WES** | **20503** | **NIAGADS DSS (NG00067.v5) / NACC** |
| **ADSP WGS** | **Whole Genome Sequencing** | **ADSP_WGS** | **16906** | **NIAGADS DSS (NG00067.v5) / NACC** |
| Indianapolis African-American | Illumina Human 1M-Duo | IIDP_AA | 1175 | NIAGADS (NG00047) |
| Indianapolis Yoruba | Illumina Omni 2.5 | IIDP_YOR | 1264 | dbGaP (phs000378) |
| CIDR | Illumina Human Omni1-Quad | CIDR | 3101 | NIAGADS (NG00015) / dbGAP (phs000496) |
| GenADA | Affymetrix 500K | GSK | 1571 | dbGaP (phs000219) |

| | | | | |
|---|---|---|---|---|
| HBTRC | Illumina Human Hap650Y | HBTRC_ILL | 338 | Synapse AMP-AD (syn3159435) |
| | Illumina Human Hap650Y | HBTRC_PERL | 402 | Synapse AMP-AD (syn3159435) |
| LATC | Illumina Multi-Ethnic – BU | LATC | 63 | RADC Rush (contact:) |
| NIA-LOAD | Illumina Human 610-Quad | LOAD | 5220 | NIAGADS (NG00020) |
| MARS | Illumina Multi-Ethnic – BU | MARS | 708 | RADC Rush (contact:) |
| MAYO | Illumina Human Hap300 | MAYO_1 | 2099 | Synapse AMP-AD (syn5591675) / NIAGADS (NG00029) |
| | Whole Genome Sequencing | AMP_AD_MAYO_WGS | 349 | Synapse AMP-AD (syn22264775) |
| MAYO2 | Illumina Omni 2.5 | MAYO_2 | 314 | Synapse AMP-AD (syn5550404) |
| | Whole Genome Sequencing | AMP_AD_MAYO_WGS | 349 | Synapse AMP-AD (syn22264775) |
| MIRAGE | Illumina Human CNV370-Duo | MIRAGE_370 | 397 | NIAGADS (NG00031) |
| | Illumina Human 610-Quad | MIRAGE_610 | 1105 | NIAGADS (NG00031) |
| MSBB | Whole Genome Sequencing | AMP_AD_MSBB_WGS | 349 | Synapse AMP-AD (syn3159438, syn22264775) |
| MTC | Illumina Human OmniExpress | MTC | 542 | NIAGADS (NG00096) |
| OHSU | Illumina Human CNV370-Duo | OHSU | 647 | NIAGADS (NG00017) |
| ROSMAP | Affymetrix GeneChip 6.0 - Broad Institute | ROSMAP_1B | 1126 | RADC Rush (contact:) / Synapse AMP-AD |
| | Affymetrix GeneChip 6.0 - TGen | ROSMAP_1T | 582 | RADC Rush (contact:) / Synapse AMP-AD |
| | Illumina Human OmniExpress 12 - Chop | ROSMAP_2C | 382 | RADC Rush (contact:) / Synapse AMP-AD |
| | Illumina Multi-Ethnic - BU | ROSMAP_3BU | 494 | RADC Rush (contact:) |
| | Whole Genome Sequencing | AMP_AD_ROSMAP_WGS | 1196 | RADC Rush (contact:) / Synapse AMP-AD |
| TARCC | Affymetrix 6.0 | TARCC | 625 | NIAGADS (NG00097) |
| | Illumina Multi-Ethnic – BU | TARCC_full | 2718 | TARCC (contact:) |
| TGEN2 | Affymetrix 6.0 | TGEN | 1599 | NIAGADS (NG00028) |
| UPITT | Illumina Human Omni1-Quad | UPITT | 2440 | NIAGADS (NG00026) |
| UM/VU/MSSM | Illumina Human 1M-Duo, Illumina 1M | UVM_A | 1153 | NIAGADS (NG00042) |
| | Affymetrix 6.0 | UVM_B | 864 | NIAGADS (NG00042) |
| | Illumina Human 550K. Illumina Human 610-Quad | UVM_C | 445 | NIAGADS (NG00042) |
| WASHU | Illumina Human 610-Quad | WASHU_1 | 670 | NIAGADS (NG00030) |
| WASHU2 | Illumina Human OmniExpress | WASHU_2 | 235 | NIAGADS (NG00087) |
| WHICAP | Illumina Human OmniExpress | WHICAP | 647 | NIAGADS (NG00093) |

**Table S2. Overview of ADSP studies with WES or WGS available through NIAGADS DSS (NG00067).**

| Study | Accession Number | Related Datasets |
|---|---|---|
| Accelerating Medicines Partnership- Alzheimer's Disease (AMP-AD) | sa000011 | NG00067 – ADSP Umbrella |
| Cache County Study | sa000014 | NG00067 – ADSP Umbrella |
| University of Pittsburgh- Kamboh WGS | sa000012 | NG00067 – ADSP Umbrella |
| CurePSP and Tau Consortium PSP WGS | sa000016 | NG00067 – ADSP Umbrella |
| NIH, CurePSP and Tau Consortium PSP WGS | sa000015 | NG00067 – ADSP Umbrella |
| UCLA Progressive Supranuclear Palsy | sa000017 | NG00067 – ADSP Umbrella |
| NACC Genentech WGS | sa000013 | NG00067 – ADSP Umbrella |
| Alzheimer's Disease Sequencing Project (ADSP) | sa000001 | NG00067 – ADSP Umbrella |
| Alzheimer's Disease Neuroimaging Initiative (ADNI) | sa000002 | NG00067 – ADSP Umbrella |
| Alzheimer's Disease Genetics Consortium: African Americans (ADGC AA) | sa000003 | NG00067 – ADSP Umbrella |
| The Familial Alzheimer Sequencing (FASe) project | sa000004 | NG00067 – ADSP Umbrella |
| Brkanac – Family-based genome scan for AAO of LOAD | sa000005 | NG00067 – ADSP Umbrella |
| HIHG Miami Families with AD | sa000006 | NG00067 – ADSP Umbrella |
| Washington Heights/Inwood Columbia Aging Project (WHICAP) | sa000007 | NG00067 – ADSP Umbrella |
| Charles F. and Joanne Knight Alzheimer's Disease Research Center (Knight ADRC) | sa000008 | NG00067 – ADSP Umbrella |
| Corticobasal degeneration Study (CBD) | sa000009 | NG00067 – ADSP Umbrella |
| Progressive Supranuclear Palsy Study (PSP) | sa000010 | NG00067 – ADSP Umbrella |

**Table S3. ADSP WES Sample sizes per center after sequential quality control and filtering steps (detailed in column titles).**

| Sequencing Centers | 1. All genotyped subjects | 2. No sex problems | 3. No ancestry/race discordance | 4. No duplicate discordance | 5. Filter to CN/AD | 6. *APOE* genotype available | 7. AGE available | 8. AGE 60y and up | 9. European (EU) | 10. Retain unique non-duplicate |
|---|---|---|---|---|---|---|---|---|---|---|
| ADSP_WES_Baylor | 2368 | 2367 | 2364 | 2359 | 2330 | 2327 | 2327 | 2308 | 2221 | 2203 |
| ADSP_WES_Broad | 4584 | 4583 | 4574 | 4562 | 4259 | 4259 | 4259 | 4249 | 4228 | 4222 |
| ADSP_WES_CHOP | 346 | 345 | 343 | 341 | 1 | 1 | 1 | 1 | 1 | 1 |
| ADSP_WES_CU_IGM | 3861 | 3861 | 3823 | 3811 | 3759 | 3758 | 3731 | 3730 | 830 | 719 |
| ADSP_WES_FGC | 330 | 330 | 329 | 327 | 0 | 0 | 0 | 0 | 0 | 0 |
| ADSP_WES_IDOM | 103 | 103 | 102 | 102 | 0 | 0 | 0 | 0 | 0 | 0 |
| ADSP_WES_MGI | 1036 | 1033 | 1027 | 1020 | 892 | 892 | 891 | 806 | 776 | 747 |
| ADSP_WES_Otogenetics | 714 | 714 | 714 | 705 | 608 | 608 | 608 | 605 | 594 | 564 |
| ADSP_WES_PGFI | 117 | 117 | 117 | 117 | 0 | 0 | 0 | 0 | 0 | 0 |
| ADSP_WES_UM_HIHG | 3265 | 3265 | 3248 | 3244 | 3035 | 3035 | 3009 | 2945 | 94 | 84 |
| ADSP_WES_UW_GenomeSciences | 75 | 75 | 73 | 67 | 53 | 50 | 50 | 44 | 36 | 30 |
| ADSP_WES_WashU | 3704 | 3702 | 3687 | 3661 | 3353 | 3353 | 3352 | 3337 | 3012 | 3003 |
| Total | 20503 | 20495 | 20401 | 20316 | 18290 | 18283 | 18228 | 18025 | 11792 | 11573 |

**Table S4. ADSP WGS Sample sizes per center after sequential quality control and filtering steps (detailed in column titles).**

| **Sequencing Centers** | **1. All genotyped subjects** | **2. No sex problems** | **3. No ancestry/race discordance** | **4. No duplicate discordance** | **5. Filter to CN/AD** | **6. *APOE* genotype available** | **7. AGE available** | **8. AGE 60y and up** | **9. European (EU)** | **10. Retain unique non-duplicate** |
|---|---|---|---|---|---|---|---|---|---|---|
| ADSP_WGS_BAYLOR | 1272 | 1272 | 1268 | 1268 | 1228 | 1228 | 1228 | 1207 | 119 | 119 |
| ADSP_WGS_BROAD | 1493 | 1492 | 1489 | 1482 | 1330 | 1330 | 1329 | 1277 | 1018 | 1000 |
| ADSP_WGS_GENENTECH | 55 | 55 | 52 | 51 | 49 | 49 | 48 | 26 | 26 | 21 |
| ADSP_WGS_ILLUMINA | 1450 | 1449 | 1418 | 1417 | 820 | 820 | 820 | 760 | 730 | 682 |
| ADSP_WGS_MACROGEN | 886 | 886 | 879 | 879 | 1 | 1 | 1 | 1 | 1 | 1 |
| ADSP_WGS_NYGC | 1646 | 1638 | 1621 | 1569 | 1192 | 1192 | 1192 | 1192 | 1160 | 1148 |
| ADSP_WGS_USUHS | 8777 | 8773 | 8704 | 8676 | 7240 | 7239 | 6939 | 6422 | 3474 | 3423 |
| ADSP_WGS_WASHU | 1327 | 1326 | 1310 | 1308 | 1242 | 1241 | 1240 | 1202 | 143 | 139 |
| Total | 16906 | 16891 | 16741 | 16650 | 13102 | 13100 | 12797 | 12087 | 6671 | 6533 |

### Table S5. Sample demographics for ADSP WES analyses, stratified by Sequencing Center and Kit.

| ADSP WES samples | | Diagnosis | | Sex | Age | *APOE* status | |
|---|---|---|---|---|---|---|---|
| **Name** | **Participants after QC (N)** | **Type** | **(N)** | **Female (N (%))** | **Age (Mean (SD))** | **APOE*4-pos** | **APOE*2-pos** |
| **All samples** | 11573 | CN | 5418 | 3152 (58.2 %) | 85.4 (6.5) | 926 (17.1 %) | 1057 (19.5 %) |
| | | AD | 6155 | 3619 (58.8 %) | 75.4 (8.6) | 2938 (47.7 %) | 493 (8.0 %) |
| **Sequencing Centers** | | | | | | | |
| **Baylor** | 2203 | CN | 1100 | 597 (64.3 %) | 86.2 (3.9) | 435 (39.4 %) | 243 (22.1 %) |
| | | AD | 1103 | 689 (62.5 %) | 76.2 (5.8) | 689 (62.5 %) | 113 (10.2 %) |
| **Broad** | 4222 | CN | 1606 | 995 (62.0 %) | 88.1 (4.8) | 1277 (48.8 %) | 366 (22.8 %) |
| | | AD | 2616 | 1412 (54.0 %) | 73.9 (8.4) | 1412 (54.0 %) | 203 (7.8 %) |
| **CHOP** | 1 | CN | 0 | - | - | 1 (100.0 %) | - |
| | | AD | 1 | 0 (0.0 %) | 75.0 (-) | 0 (0.0 %) | 0 (0.0 %) |
| **CU_IGM** | 719 | CN | 667 | 395 (59.2 %) | 79.8 (6.7) | 159 (23.8 %) | 75 (11.2 %) |
| | | AD | 52 | 31 (59.6 %) | 83.9 (6.9) | 15 (28.8 %) | 9 (17.3 %) |
| **MGI** | 747 | CN | 356 | 206 (57.9 %) | 74.1 (7.5) | 125 (35.1 %) | 54 (15.2 %) |
| | | AD | 391 | 210 (53.7 %) | 72.5 (7.3) | 262 (67.0 %) | 22 (5.6 %) |
| **Otogenetics** | 564 | CN | 146 | 83 (56.8 %) | 79.8 (7.3) | 68 (46.6 %) | 12 (8.2 %) |
| | | AD | 418 | 279 (66.7 %) | 74.0 (7.0) | 320 (76.6 %) | 11 (2.6 %) |
| **UM_HIHG** | 84 | CN | 17 | 6 (35.3 %) | 76.2 (6.8) | 4 (23.5 %) | 1 (5.9 %) |
| | | AD | 67 | 46 (68.7 %) | 76.2 (5.8) | 38 (56.7 %) | 6 (9.0 %) |
| **UW_GenomeSciences** | 30 | CN | 0 | - | - | - | - |
| | | AD | 30 | 19 (63.3 %) | 72.9 (7.7) | 19 (63.3 %) | 1 (3.3 %) |
| **WashU** | 3003 | CN | 1526 | 870 (57.0 %) | 87.9 (4.1) | 204 (13.4 %) | 306 (20.5 %) |
| | | AD | 1477 | 933 (63.2 %) | 78.4 (8.6) | 571 (38.7 %) | 128 (8.7 %) |
| **Sequencing Kits** | | | | | | | |
| **HiSeq_2000** | 11021 | CN | 5125 | 2980 (58.1 %) | 86.1 (5.8) | 837 (16.3 %) | 1010 (19.7 %) |
| | | AD | 5896 | 3486 (59.1 %) | 75.5 (8.6) | 2791 (47.3 %) | 417 (8.0 %) |
| **HiSeq_2000/2500** | 84 | CN | 17 | 6 (35.3 %) | 76.2 (6.8) | 4 (23.5 %) | 1 (5.9 %) |
| | | AD | 67 | 46 (68.7 %) | 76.2 (5.8) | 38 (56.7 %) | 6 (9.0 %) |
| **HiSeq_2500** | 1 | CN | 0 | - | - | - | - |
| | | AD | 1 | 0 (0.0 %) | 75.0 (-) | 1 (100.0 %) | 0 (0.0 %) |
| **HiSeq_4000** | 467 | CN | 276 | 166 (60.1 %) | 73.1 (7.4) | 85 (30.8 %) | 46 (16.7 %) |
| | | AD | 191 | 87 (45.5 %) | 71.0 (7.4) | 108 (56.5 %) | 16 (8.4 %) |

### Table S6. Sample demographics for ADSP WGS analyses, stratified by Sequencing Center and Kit.

| ADSP WGS samples | | Diagnosis | | Sex | Age | *APOE* status | |
|---|---|---|---|---|---|---|---|
| **Name** | **Participants after QC (N)** | **Type** | **(N)** | **Female (N (%))** | **Age (Mean (SD))** | **APOE*4-pos** | **APOE*2-pos** |
| **All samples** | 6533 | CN | 2949 | 1791 (60.7 %) | 81.6 (6.6) | 1075 (36.4 %) | 204 (6.9 %) |
| | | AD | 3584 | 2051 (57.2 %) | 76.7 (8.3) | 2078 (58.0 %) | 177 (4.9 %) |
| **Sequencing Centers** | | | | | | | |
| **Baylor** | 119 | CN | 75 | 48 (64.0 %) | 81.0 (4.5) | 25 (33.3 %) | 5 (6.7 %) |
| | | AD | 44 | 37 (84.1 %) | 74.5 (6.8) | 29 (65.9 %) | 2 (4.5 %) |
| **Broad** | 1000 | CN | 437 | 265 (60.6 %) | 81.3 (6.6) | 91 (20.8 %) | 68 (15.6 %) |
| | | AD | 563 | 303 (53.8 %) | 75.0 (8.8) | 353 (62.7 %) | 29 (5.2 %) |
| **Genentech** | 21 | CN | 6 | 1 (16.7 %) | 80.8 (2.9) | 6 (100.0 %) | 0 (0.0 %) |
| | | AD | 15 | 12 (80.0 %) | 76.2 (6.7) | 6 (40.0 %) | 4 (26.7 %) |
| **Illumina** | 682 | CN | 409 | 225 (55.0 %) | 80.3 (5.9) | 253 (61.9 %) | 32 (7.8 %) |
| | | AD | 273 | 111 (40.7 %) | 71.6 (7.8) | 207 (75.8 %) | 10 (3.7 %) |
| **Macrogen** | 1 | CN | 0 | - | - | - | - |
| | | AD | 1 | 0 (0.0 %) | 64.0 (-) | 0 (0 %) | 0 (0 %) |
| **NYGC** | 1148 | CN | 300 | 187 (62.3 %) | 85.4 (7.0) | 44 (14.7 %) | 60 (15.6 %) |
| | | AD | 848 | 566 (66.7 %) | 80.4 (8.6) | 432 (50.9 %) | 69 (8.1 %) |
| **USHS** | 3423 | CN | 1682 | 1034 (61.5 %) | 81.6 (6.3) | 643 (38.2 %) | 33 (2.0 %) |
| | | AD | 1741 | 961 (55.2 %) | 76.4 (7.3) | 1006 (57.8 %) | 55 (3.2 %) |
| **WashU** | 139 | CN | 40 | 31 (77.5 %) | 70.8 (8.4) | 13 (32.5 %) | 6 (15.0 %) |
| | | AD | 99 | 61 (61.6 %) | 76.2 (8.1) | 45 (45.5 %) | 8 (8.1 %) |
| **Sequencing Kits** | | | | | | | |
| **Illumina_HiSeq_2000** | 569 | CN | 210 | 105 (50.0 %) | 79.7 (7.6) | 50 (23.8 %) | 34 (16.2 %) |
| | | AD | 359 | 193 (53.8 %) | 75.0 (7.0) | 223 (62.1 %) | 20 (5.6 %) |
| **Illumina_HiSeq_X_Ten** | 1014 | CN | 523 | 325 (62.1 %) | 80.7 (6.9) | 118 (22.6 %) | 74 (14.1 %) |
| | | AD | 491 | 259 (52.7 %) | 76.3 (8.5) | 286 (58.2 %) | 29 (5.9 %) |
| **PCRAmplified_HiSeq2000** | 304 | CN | 218 | 129 (59.1 %) | 80.8 (3.7) | 213 (97.7 %) | 0 (0.0 %) |
| | | AD | 86 | 39 (45.3 %) | 65.7 (6.5) | 77 (89.5 %) | 4 (4.7 %) |
| **PCRAmplified_HiSeq2000/2500** | 15 | CN | 12 | 7 (58.3 %) | 67.8 (3.7) | 3 (25.0 %) | 2 (16.7 %) |
| | | AD | 3 | 0 (0.0 %) | 76.7 (5.5) | 2 (66.7 %) | 0 (0.0 %) |
| **PCRAmplified_HiSeqX** | 65 | CN | 8 | 5 (62.5 %) | 86.1 (5.8) | 6 (75.0 %) | 1 (12.5 %) |
| | | AD | 57 | 33 (57.9 %) | 75.5 (8.6) | 53 (93.0 %) | 0 (0.0 %) |
| **PCRFree_HiSeqX** | 3965 | CN | 1777 | 1117 (62.9 %) | 82.7 (6.4) | 591 (33.3 %) | 68 (3.8 %) |
| | | AD | 2188 | 1343 (61.4 %) | 78.6 (7.8) | 1190 (54.4 %) | 101 (4.6 %) |
| **PCRFree_Novaseq** | 601 | CN | 201 | 103 (62.9 %) | 77.9 (6.3) | 94 (46.8 %) | 25 (12.4 %) |
| | | AD | 400 | 184 (46.0 %) | 72.9 (7.5) | 247 (61.8 %) | 23 (5.8 %) |

**Table S7. Illustration of a variant underlying an artifactual genome-wide significant signal in model-1 after applying duplicate discordance filters.** The displayed variant (rs895262150) corresponds to the hit annotated with gene "*ZDHHC14*" in Figure S16. Note the enrichment in control carriers on the Illumina center and PCR amplified HiSeq2000 kit. Closer inspection revealed that this variant shows a considerably higher variant missingness rate in subjects sequenced using the PCR amplified HiSeq2000 kit compared to other kits (40%). Control individuals on this kit were almost solely contributed by a single cohort (G-CCS) with a variant missingness rate of 50%, whereas cases were mainly contributed by two other cohorts (A-ADC, A-LOAD), with a variant missingness rate rate of 25%. All control variant carriers were contributed by a single cohort (G-CCS). These observations confirm the variant appears artifactual.

| Sequencing Center | | ADSP_WGS_BAYLOR | ADSP_WGS_BROAD | ADSP_WGS_GENENTECH | ADSP_WGS_ILLUMINA | ADSP_WGS_MACROGEN | ADSP_WGS_NYGC | ADSP_WGS_USUHS | ADSP_WGS_WASHU |
|---|---|---|---|---|---|---|---|---|---|
| **AD** | **WT** | 40 | 561 | 4 | 239 | 1 | 846 | 1735 | 84 |
| | **HET** | 0 | 0 | 0 | 3 | 0 | 2 | 5 | 0 |
| | **HOM** | 0 | 0 | 0 | 0 | 0 | 0 | 0 | 0 |
| **CN** | **WT** | 75 | 435 | 0 | 257 | 0 | 300 | 1672 | 37 |
| | **HET** | 0 | 2 | 0 | 35 | 0 | 0 | 10 | 0 |
| | **HOM** | 0 | 0 | 0 | 0 | 0 | 0 | 0 | 0 |

| Sequencing Kit | | Illumina_HiSeq_2000 | Illumina_HiSeq_X_Ten | PCRAmplified_HiSeq2000 | PCRAmplified_HiSeq2000/2500 | PCRAmplified_HiSeqX | PCRFree_HiSeqX | PCRFree_Novaseq |
|---|---|---|---|---|---|---|---|---|
| **AD** | **WT** | 315 | 491 | 64 | 3 | 57 | 2180 | 354 |
| | **HET** | 2 | 0 | 1 | 0 | 0 | 7 | 0 |
| | **HOM** | 0 | 0 | 0 | 0 | 0 | 0 | 0 |
| **CN** | **WT** | 183 | 521 | 84 | 12 | 8 | 1769 | 46 |
| | **HET** | 2 | 2 | 33 | 0 | 0 | 8 | 1 |
| | **HOM** | 0 | 0 | 0 | 0 | 0 | 0 | 0 |

**Table S8 (part 1). ADSP WGS variants passing suggestive significance after applying centers/kit-based filters.** Note that many variants that lose suggestive significance after center/kit adjustment in model-2 have fairly small P-values (but above threshold) in the center/kit Fisher tests and/or are present in the gnomAD-based or duplicate discordance variant filters.

| Variant info | | | | | | | Model 1 | | | | Model 2 | | | | Filters | | | |
|---|---|---|---|---|---|---|---|---|---|---|---|---|---|---|---|---|---|---|
| **GENE** | **CHR** | **BP** | **dbSNP153 ID** | **effect allele** | **other allele** | **effect allele frq.** | **OR** | **95% CI (lb)** | **95% CI (ub)** | **P** | **OR** | **95% CI (lb)** | **95% CI (ub)** | **P** | **Center Fisher P** | **Kit Fisher P** | **gnomAD filter** | **Duplicate check** |
| LINC01648 | 1 | 30307706 | rs147201606 | T | C | 1.11% | 0.48 | 0.35 | 0.66 | **6.7E-06** | 0.50 | 0.37 | 0.68 | **7.9E-06** | 0.98 | 0.98 | PASS | ok |
| LINC01343 | 1 | 38312264 | rs140123944 | A | G | 1.22% | 0.51 | 0.38 | 0.68 | **8.3E-06** | 0.54 | 0.40 | 0.72 | 2.3E-05 | 0.93 | 0.97 | PASS | ok |
| CR1 | 1 | 207510847 | rs12037841 | T | G | 19.96% | 1.21 | 1.11 | 1.32 | **9.5E-06** | 1.19 | 1.10 | 1.29 | 1.8E-05 | 0.03 | 0.94 | PASS | ok |
| CNIH3 | 1 | 224768842 | rs193214846 | T | C | 0.68% | 0.40 | 0.27 | 0.61 | 1.4E-05 | 0.42 | 0.28 | 0.61 | **1.0E-05** | 0.97 | 0.96 | PASS | ok |
| DNAH14 | 1 | 225331510 | rs41303992 | A | G | 0.34% | 0.26 | 0.15 | 0.46 | **4.5E-06** | 0.29 | 0.17 | 0.51 | **1.0E-05** | 0.98 | 0.96 | PASS | ok |
| GPR137B | 1 | 236155980 | rs187878039 | T | C | 0.19% | 0.14 | 0.06 | 0.30 | **3.9E-07** | 0.17 | 0.08 | 0.35 | **2.1E-06** | 0.99 | 0.89 | LCR | ok |
| LINC01814 | 2 | 8511225 | - | G | GAA | 70.11% | 1.19 | 1.10 | 1.28 | **3.6E-06** | 1.14 | 1.07 | 1.22 | 1.7E-04 | 0.08 | 1.2E-04 | PASS | ok |
| LINC01884 | 2 | 22439246 | rs115335046 | G | A | 0.25% | 0.22 | 0.11 | 0.43 | **1.0E-05** | 0.24 | 0.13 | 0.46 | 1.8E-05 | 0.98 | 0.98 | PASS | ok |
| GALNT14 | 2 | 31129922 | rs11676188 | G | C | 20.69% | 0.84 | 0.77 | 0.91 | 3.2E-05 | 0.84 | 0.77 | 0.91 | **1.0E-05** | 0.66 | 0.19 | PASS | ok |
| PLEK | 2 | 68419120 | rs149490106 | T | C | 0.76% | 0.44 | 0.30 | 0.64 | 2.1E-05 | 0.43 | 0.30 | 0.62 | **6.2E-06** | 0.98 | 0.93 | PASS | ok |
| FAHD2CP | 2 | 96008264 | rs138643748 | C | G | 1.06% | 0.45 | 0.33 | 0.63 | **2.0E-06** | 0.58 | 0.42 | 0.79 | 6.4E-04 | 1.4E-03 | 5.3E-04 | PASS | discordant |
| BIN1 | 2 | 127135234 | rs6733839 | T | C | 40.39% | 1.26 | 1.18 | 1.35 | **2.3E-11** | 1.25 | 1.17 | 1.33 | **3.9E-11** | 0.62 | 0.02 | PASS | ok |
| MYO3B-AS1 | 2 | 170490308 | rs111867349 | A | G | 1.87% | 0.59 | 0.47 | 0.76 | 3.1E-05 | 0.59 | 0.47 | 0.74 | **9.0E-06** | 0.41 | 0.46 | PASS | ok |
| INPP5D | 2 | 233073186 | rs72982255 | A | G | 13.02% | 0.80 | 0.72 | 0.88 | **1.0E-05** | 0.81 | 0.74 | 0.89 | 1.3E-05 | 0.90 | 4.8E-04 | PASS | ok |
| CNTN4 | 3 | 1780132 | rs113207766 | TTAAA | T | 49.63% | 0.82 | 0.77 | 0.87 | **4.7E-09** | 0.83 | 0.78 | 0.89 | **1.7E-08** | 0.08 | 0.15 | PASS | ok |
| ACKR2 | 3 | 42837221 | rs879898582 | GT | G | 0.49% | 3.83 | 2.36 | 6.21 | **5.7E-08** | 1.81 | 1.09 | 3.03 | 0.02 | 0.97 | 0.88 | non-PASS | discordant |
| FHIT | 3 | 59786111 | rs374541147 | C | A | 1.63% | 0.52 | 0.40 | 0.68 | **2.0E-06** | 0.55 | 0.42 | 0.71 | **3.7E-06** | 0.96 | 0.96 | PASS | ok |
| - | 3 | 110419318 | rs115395207 | T | G | 1.38% | 0.50 | 0.37 | 0.66 | **2.0E-06** | 0.53 | 0.40 | 0.70 | **6.9E-06** | 0.96 | 0.96 | PASS | ok |
| ITGB5 | 3 | 124876108 | rs1948696 | T | C | 37.20% | 1.19 | 1.11 | 1.27 | **1.5E-06** | 1.16 | 1.09 | 1.24 | **8.1E-06** | 0.65 | 0.50 | PASS | ok |
| LINC01565 | 3 | 128575196 | rs532515415 | A | G | 0.25% | 0.26 | 0.14 | 0.51 | 8.1E-05 | 0.23 | 0.12 | 0.44 | **6.0E-06** | 0.98 | 0.95 | PASS | ok |
| BFSP2 | 3 | 133401263 | rs138196830 | A | T | 0.14% | 0.13 | 0.05 | 0.32 | **9.3E-06** | 0.16 | 0.07 | 0.37 | 2.6E-05 | 0.98 | 0.96 | PASS | ok |
| STPG2-AS1 | 4 | 97428849 | rs866286162 | C | G | 0.47% | 0.32 | 0.19 | 0.52 | **6.7E-06** | 0.39 | 0.25 | 0.63 | 1.1E-04 | 0.98 | 0.98 | PASS | ok |
| NEIL3 | 4 | 177342041 | rs34539659 | C | A | 0.30% | 0.25 | 0.13 | 0.46 | **9.0E-06** | 0.32 | 0.18 | 0.58 | 1.6E-04 | 0.98 | 0.89 | PASS | ok |
| LINC02500 | 4 | 181486820 | rs17071607 | A | G | 2.00% | 1.67 | 1.31 | 2.12 | 2.6E-05 | 1.69 | 1.34 | 2.12 | **7.7E-06** | 0.98 | 0.96 | PASS | ok |
| MTRR | 5 | 7931168 | rs6883636 | A | T | 7.05% | 1.34 | 1.18 | 1.53 | **1.0E-05** | 1.28 | 1.13 | 1.45 | 9.0E-05 | 0.50 | 0.98 | PASS | ok |
| MYO10 | 5 | 16924467 | rs112418255 | T | C | 0.35% | 0.28 | 0.16 | 0.49 | **7.9E-06** | 0.35 | 0.20 | 0.59 | 1.1E-04 | 0.97 | 0.02 | PASS | ok |
| ARSB | 5 | 78811930 | rs75497745 | C | T | 2.50% | 0.63 | 0.51 | 0.79 | 3.2E-05 | 0.63 | 0.51 | 0.77 | **9.2E-06** | 0.98 | 0.96 | PASS | ok |
| LINC01340 | 5 | 97649168 | rs145076322 | T | A | 0.26% | 0.22 | 0.11 | 0.43 | **8.8E-06** | 0.26 | 0.14 | 0.49 | 3.1E-05 | 0.95 | 0.98 | PASS | ok |
| YIPF5 | 5 | 144039218 | rs113589858 | AT | A | 9.88% | 1.35 | 1.20 | 1.51 | **2.6E-07** | 1.30 | 1.16 | 1.45 | **2.2E-06** | 0.03 | 0.02 | PASS | ok |
| YIPF5 | 5 | 144078830 | rs7708467 | C | T | 25.16% | 1.18 | 1.09 | 1.28 | 2.1E-05 | 1.20 | 1.11 | 1.29 | **2.0E-06** | 0.91 | 0.96 | PASS | ok |
| CYFIP2 | 5 | 157393949 | rs6555992 | G | A | 18.42% | 0.81 | 0.75 | 0.89 | **3.4E-06** | 0.82 | 0.75 | 0.89 | **1.4E-06** | 0.96 | 0.93 | PASS | ok |
| LOC401312 | 7 | 22662017 | rs10265117 | A | G | 12.66% | 0.80 | 0.72 | 0.88 | 1.3E-05 | 0.80 | 0.73 | 0.88 | **4.6E-06** | 0.65 | 0.18 | PASS | ok |
| SUGCT | 7 | 40867131 | rs146711196 | T | TATA | 32.26% | 0.86 | 0.80 | 0.93 | 5.0E-05 | 0.85 | 0.79 | 0.91 | **4.0E-06** | 0.19 | 0.95 | LCR | ok |
| YWHAEP1 | 7 | 64472092 | rs143068421 | A | C | 0.73% | 0.39 | 0.26 | 0.57 | **2.3E-06** | 0.43 | 0.30 | 0.63 | 1.3E-05 | 1.7E-03 | 0.96 | PASS | ok |
| STYXL1 | 7 | 76006286 | rs182846177 | G | C | 0.32% | 0.30 | 0.16 | 0.53 | 5.0E-05 | 0.28 | 0.16 | 0.49 | **1.0E-05** | 0.98 | 0.93 | PASS | ok |

**Table S8 (part 2). ADSP WGS variants passing suggestive significance after applying centers/kit-based filters.** Note that many variants that lose suggestive significance after center/kit adjustment in model-2 have fairly small P-values (but above threshold) in the center/kit Fisher tests and/or are present in the gnomAD-based or duplicate discordance variant filters.

| Variant info | | | | | | | Model 1 | | | | Model 2 | | | | Filters | | | |
|---|---|---|---|---|---|---|---|---|---|---|---|---|---|---|---|---|---|---|
| GENE | CHR | BP | dbSNP153 ID | effect allele | other allele | effect allele frq. | OR | 95% CI (lb) | 95% CI (ub) | P | OR | 95% CI (lb) | 95% CI (ub) | P | Center Fisher P | Kit Fisher P | gnomAD filter | Duplicate check |
| LRRC4 | 7 | 128041888 | rs76593352 | T | G | 0.77% | 0.40 | 0.27 | 0.58 | **2.4E-06** | 0.41 | 0.29 | 0.59 | **2.1E-06** | 0.90 | 0.30 | PASS | ok |
| KCNV2 | 9 | 2751431 | rs543363829 | T | C | 0.31% | 0.29 | 0.16 | 0.53 | 6.6E-05 | 0.27 | 0.15 | 0.47 | **7.0E-06** | 0.98 | 0.96 | PASS | ok |
| PTPRD-AS1 | 9 | 8880476 | rs138987427 | A | G | 0.27% | 3.49 | 1.83 | 6.68 | 1.6E-04 | 4.06 | 2.19 | 7.55 | **9.3E-06** | 0.99 | 0.99 | PASS | ok |
| LINGO2 | 9 | 28300621 | rs4587420 | G | C | 38.87% | 0.85 | 0.79 | 0.91 | **2.4E-06** | 0.85 | 0.80 | 0.91 | **1.8E-06** | 1.6E-03 | 0.16 | PASS | ok |
| C9orf85 | 9 | 71914326 | rs113367159 | T | TG | 0.22% | 5.66 | 2.75 | 11.67 | **2.8E-06** | 3.60 | 1.77 | 7.33 | 4.0E-04 | 0.99 | 0.99 | LCR | discordant |
| SHC3 | 9 | 89033784 | rs1537144 | A | C | 19.51% | 0.84 | 0.77 | 0.91 | 3.5E-05 | 0.83 | 0.76 | 0.90 | **4.2E-06** | 0.92 | 0.43 | PASS | ok |
| RAD23B | 9 | 107355656 | rs10739241 | T | A | 41.20% | 0.86 | 0.80 | 0.91 | **5.1E-06** | 0.87 | 0.81 | 0.93 | 1.5E-05 | 0.21 | 0.39 | PASS | ok |
| LINC00844 | 10 | 58995166 | rs140004050 | T | C | 0.77% | 2.06 | 1.41 | 3.01 | 1.8E-04 | 2.29 | 1.59 | 3.29 | **7.4E-06** | 0.98 | 0.98 | PASS | ok |
| ATE1 | 10 | 121774625 | rs775805330 | C | T | 0.18% | 0.16 | 0.07 | 0.35 | **6.3E-06** | 0.25 | 0.11 | 0.53 | 3.0E-04 | 0.97 | 5.2E-03 | PASS | ok |
| MIR4300HG | 11 | 81712477 | rs200761787 | A | AG | 1.52% | 0.53 | 0.41 | 0.70 | **5.2E-06** | 0.56 | 0.43 | 0.72 | **8.7E-06** | 0.97 | 0.95 | PASS | ok |
| ARHGAP42-AS1 | 11 | 100680925 | rs758006599 | A | T | 0.38% | 0.29 | 0.17 | 0.50 | **8.5E-06** | 0.33 | 0.20 | 0.56 | 3.4E-05 | 0.98 | 0.96 | LCR | ok |
| LOC101928535 | 11 | 106426120 | rs117682555 | T | C | 2.21% | 1.58 | 1.26 | 1.99 | 9.0E-05 | 1.65 | 1.33 | 2.06 | **7.2E-06** | 0.96 | 0.11 | PASS | ok |
| ETV6 | 12 | 11698764 | rs2724652 | G | T | 45.30% | 0.85 | 0.80 | 0.91 | **5.4E-06** | 0.86 | 0.80 | 0.91 | **3.0E-06** | 6.65E-05 | 0.38 | PASS | ok |
| CAPRIN2 | 12 | 30718899 | rs188591971 | T | G | 0.64% | 2.30 | 1.52 | 3.48 | 8.7E-05 | 2.46 | 1.66 | 3.66 | **7.9E-06** | 0.97 | 0.02 | PASS | ok |
| LINC02451 | 12 | 42653330 | rs184022552 | A | G | 7.45% | 0.74 | 0.65 | 0.84 | **4.6E-06** | 0.76 | 0.67 | 0.86 | 1.5E-05 | 2.7E-04 | 0.15 | PASS | ok |
| LINC02451 | 12 | 42657809 | rs141146804 | C | T | 7.36% | 0.74 | 0.65 | 0.85 | **8.1E-06** | 0.75 | 0.66 | 0.85 | **5.0E-06** | 2.7E-04 | 0.09 | PASS | ok |
| ATP8A2 | 13 | 25623164 | rs547117207 | A | G | 0.25% | 3.87 | 1.97 | 7.61 | 8.1E-05 | 4.41 | 2.31 | 8.41 | **6.5E-06** | 0.99 | 0.99 | PASS | ok |
| VWA8 | 13 | 41939480 | rs796820552 | TA | T | 0.40% | 3.69 | 2.17 | 6.28 | **1.6E-06** | 1.95 | 1.12 | 3.40 | 0.02 | 0.98 | 0.98 | non-PASS | discordant |
| DNAJC15 | 13 | 43061521 | rs143663531 | G | A | 0.60% | 2.70 | 1.75 | 4.17 | **7.3E-06** | 2.50 | 1.65 | 3.78 | 1.6E-05 | 0.98 | 0.99 | PASS | ok |
| MIR4704 | 13 | 66170147 | rs9540673 | T | G | 16.61% | 0.82 | 0.75 | 0.89 | **1.0E-05** | 0.83 | 0.76 | 0.91 | 3.2E-05 | 0.60 | 0.97 | PASS | ok |
| MIR4539 | 14 | 105778867 | rs188538741 | T | A | 0.61% | 2.74 | 1.79 | 4.22 | **3.9E-06** | 2.51 | 1.66 | 3.78 | 1.1E-05 | 0.98 | 0.08 | PASS | ok |
| LOC102723493 | 15 | 67048252 | rs78650348 | C | A | 1.82% | 0.59 | 0.46 | 0.76 | 3.4E-05 | 0.57 | 0.45 | 0.72 | **3.6E-06** | 0.90 | 0.09 | PASS | ok |
| CALML4 | 15 | 68164398 | rs148101423 | G | A | 0.21% | 0.19 | 0.09 | 0.39 | **1.0E-05** | 0.21 | 0.10 | 0.42 | 1.2E-05 | 0.97 | 0.99 | PASS | ok |
| FAM169B | 15 | 98506192 | rs4465592 | G | C | 35.69% | 1.17 | 1.10 | 1.26 | **5.1E-06** | 1.16 | 1.08 | 1.24 | 1.2E-05 | 0.87 | 0.76 | PASS | ok |
| FAM169B | 15 | 98528459 | rs72766230 | G | A | 13.74% | 1.22 | 1.11 | 1.35 | 5.0E-05 | 1.23 | 1.13 | 1.35 | **7.9E-06** | 0.60 | 0.27 | PASS | ok |
| BMERB1 | 16 | 15459834 | rs56189737 | A | G | 2.43% | 1.66 | 1.34 | 2.06 | **3.9E-06** | 1.57 | 1.28 | 1.93 | 1.8E-05 | 0.79 | 0.06 | PASS | ok |
| PIRT | 17 | 10902147 | rs34731238 | T | C | 7.06% | 0.74 | 0.65 | 0.85 | **8.4E-06** | 0.77 | 0.68 | 0.87 | 5.0E-05 | 0.02 | 0.38 | PASS | ok |
| KIF2B | 17 | 54042628 | rs137879811 | C | T | 0.66% | 2.70 | 1.79 | 4.09 | **2.7E-06** | 2.59 | 1.74 | 3.84 | **2.5E-06** | 0.98 | 0.98 | PASS | ok |
| LOC105371855 | 17 | 62716778 | rs12602916 | G | A | 39.26% | 1.16 | 1.08 | 1.24 | 2.5E-05 | 1.16 | 1.09 | 1.24 | **6.3E-06** | 0.90 | 0.96 | PASS | ok |
| LOC105371855 | 17 | 62722776 | rs34113842 | A | AT | 6.54% | 0.74 | 0.64 | 0.84 | **7.3E-06** | 0.76 | 0.66 | 0.86 | 2.1E-05 | 0.81 | 0.98 | PASS | ok |
| TNFRSF11A | 18 | 62385675 | - | CCG | C | 0.52% | 0.34 | 0.21 | 0.55 | **7.7E-06** | 0.38 | 0.24 | 0.59 | 2.6E-05 | 0.01 | 0.93 | PASS | discordant |
| ZNF600 | 19 | 52762647 | rs564984950 | G | A | 1.86% | 0.57 | 0.44 | 0.73 | **6.6E-06** | 0.61 | 0.48 | 0.77 | 3.0E-05 | 0.96 | 0.32 | PASS | ok |
| MCM5 | 22 | 35469225 | rs28620909 | A | G | 31.56% | 0.84 | 0.79 | 0.91 | **3.7E-06** | 0.85 | 0.80 | 0.91 | **5.6E-06** | 0.42 | 0.10 | PASS | ok |
| APOBEC3A | 22 | 38960570 | rs6001341 | A | G | 0.25% | 0.24 | 0.12 | 0.47 | 3.2E-05 | 0.22 | 0.12 | 0.43 | **6.7E-06** | 0.98 | 0.98 | PASS | ok |

**Table S9. Comparing current ADSP WES association statistics for AD risk genes/variants identified in a prior study, using a similar model and largely overlapping data.** Table shows variants identified in Le Guen et al. 2021, considering the case-control analyses not adjusting for age. We additionally highlight the rs3764645 variant on *ABCA7*, which was not present in the prior study, but was identified at suggestive signicance level here (the prior study indicated a different significant variant on *ABCA7*). Not all variants were shared across the current and prior study, which is mainly due to the differences in data releases (the prior study used the original ADSP WES and WGS discovery samples, covering fewer participants than considered here). Note that associations findings were highly consistent across both studies.

| Gene | rsID | current P-value | Le Guen et al. 2021 - P-value |
|---|---|---|---|
| ***KIF21B*** | rs2297911 | 1.60E-03 | 2.00E-04 |
| ***USH2A*** | rs111033333 | 2.40E-03 | 4.00E-03 |
| ***RAB10*** | rs149622307 | 0.31 | 0.06 |
| ***TREM2*** | rs75932628 | 8.20E-11 | 3.00E-10 |
| ***PILRA*** | rs2405442 | - | 2.10E-05 |
| ***MS4A6A*** | rs12453 | 8.90E-05 | 9.00E-06 |
| ***RIN3*** | rs150221413 | 1.80E-03 | 7.00E-03 |
| ***TAOK2*** | rs4077410 | - | 6.10E-05 |
| ***NSF/MAPT/KANSL1*** | rs199533 | 9.10E-05 | 5.10E-06 |
| ***ABCA7*** | rs547447016 | - | 1.10E-04 |
| ***ABCA7*** | rs3764645 | 3.80E-06 | - |

9. American Psychiatric Association. Diagnostic and Statistical Manual of Mental Disorders 5th edn. *Am Psychiatr Assoc*. Published online 2013.

10. American Psychiatric Association. Diagnostic and Statistical Manual of Mental Disorders 4th edn. *Am Psychiatr Assoc*. Published online 1994.

11. Hughes CP, Berg L, Danziger WL, Coben LA, Martin RL. A new clinical scale for the staging of dementia. *Br J Psychiatry*. 1982;140(6):566-572. doi:10.1192/bjp.140.6.566

12. Braak H, Braak E. Neuropathological stageing of Alzheimer-related changes. *Acta Neuropathol*. 1991;82:239-259. doi:10.1109/ICINIS.2015.10

13. Mirra SS, Heyman A, McKeel D, et al. The Consortium to Establish a Registry for Alzheimer's Disease (CERAD). Part II. Standardization of the neuropathologic assessement of Alzheimer's disease. *Neurology*. 1991;41:479-486. doi:10.1017/CBO9781107415324.004

14. Hyman BT, Trojanowski JQ. Editorial on Consensus Recommendations for the Postmortem Diagnosis of Alzheimer Disease from the National Institute on Aging and the Reagan Institute

Working Group on Diagnositic Criteria for the Neuropathological Assessment of Alzheimr Disease. *J Neuropathol Exp Neurol*. 1997;56(10):1095-1097.

26. Bennett DA, Wilson RS, Schneider JA, et al. Natural history of mild cognitive impairment in older

persons. *Neurology*. 2002;59(2):198-205. doi:10.1212/WNL.59.2.198

36. Zhu C, Farrell J, Kuzma A, Schellenberg G, Farrer L. Precision genotyping of APOE from whole genome sequencing. In: *American Society of Human Genetics Congress*. ; 2019.
